# Sexual dimorphism in human skeletal muscle ageing

**DOI:** 10.1101/2025.01.06.25319958

**Authors:** Thomas W. Fieldsend, Callum R. O’Neill, Akshita Shrivastava, Helen E. Ogden, Nick Dand, Simon M. Hughes

## Abstract

Primary sarcopenia is a progressive, age-related decline of skeletal muscle strength, size, and quality, the socio-economic and health impacts of which are set to increase due to global ageing. Despite differences in the physiology of female and male skeletal muscle being well characterised, their alteration with age is less clear. Here we report a striking sexual dimorphism in arm muscle ageing in 478,438 UK Biobank participants aged 40–82 yr. Although the sex difference in age-related arm muscle strength decline is modest, muscle mass loss is considerably more pronounced in males, both in absolute and percentage terms. We also present two alternative measures of muscle quality, each of which exhibits substantially greater age-related decline in females. These trends hold across independent analyses of separate cross-sectional and longitudinal participant groups, persist after accounting for systematic size differences between the sexes, and are apparent irrespective of female menopause status and hormone replacement therapy usage history, despite an sharp reduction in female strength during the perimenopause. Our findings confirm the importance of sex to effective diagnosis and mitigation of sarcopenia, and prompt consideration of the physiological basis of this pronounced sex difference in skeletal muscle ageing.

## Introduction

Sarcopenia is a syndrome characterised by a progressive and generalised reduction of skeletal muscle strength, size, and quality^1,2^, which is associated with a host of negative outcomes including functional impairment^3^, loss of independence^4^, lowered quality of life^5^, and increased all-cause mortality^6,7^. Although reported sarcopenia incidence is heavily dependant on the diagnostic criteria used^8^, true global prevalence in those aged 60 and above is probably ∼10%^8,9^, which would make sarcopenia more common in this age group than other ageing-related conditions such as rheumatoid arthritis (∼1–2%^10,11^) and cognitive impairment (∼5.4– 7.3%^12,13^). The combined socio-economic and health impacts of sarcopenia therefore represent a considerable ‘social burden’, the weight of which will likely increase as the global population continues to age^14^.

Although sarcopenia can occur secondary to other factors (e.g. systemic disease), it is predominantly a geriatric syndrome (‘primary sarcopenia’) ^2^. At the neuromuscular level, strength, mass, and quality of skeletal muscle can be diminished by changes in the size, number, relative abundance, force-generating capacity, and pennation angle of muscle fibres^15–18^; infiltration of intramuscular non-contractile tissue^19^; and reduced neural activation^20^. Primary sarcopenia also evidently has a large genetic component^21^, with muscle strength and size heritability generally estimated to exceed 50%^22,23^. Finally, the development and progression of primary sarcopenia may be influenced by other factors, including lifestyle^24–26^, socio-economic status^27^, ethnicity^28^, and the menopause^29^.

Humans are sexually dimorphic, with males being generally taller and heavier than females, and possessing more skeletal muscle and less adipose tissue for their size^30–36^. Body size and composition change substantially with age, with both sexes typically exhibiting an age-related decline in height^37,38^, fat-free mass, and bone mass^39,40^, whereas fat mass generally increases with age^39^. The timing and magnitude of some of these changes differ by sex^38,41^, as does the prevalence of some age-related diseases^42^. It is therefore essential from a public health perspective to identify and accurately characterise any sex differences in the age-related reductions of skeletal muscle strength, size, and quality that characterise primary sarcopenia, independent of changes in other body size parameters.

Muscle strength is typically defined as the maximum force or torque generated at maximal voluntary effort^43–45^, and can be measured during concentric, isometric, or eccentric contraction^46^. There is incontrovertible evidence that average muscle strength is considerably greater in males than females at all ages, and that muscle strength declines with age in both sexes^16,19,30,47–49^. However, it remains an open question whether sexual dimorphism exists in the age-related decline of muscle strength: although a limited number of small studies have reported greater age-related strength declines in males^50–52^, a comparable amount of evidence exists that rates of decline are in fact similar between the sexes^19,35,47,48^.

Muscle size can be quantified as muscle mass, volume, or cross-sectional area^30,53,54^. Any sex differences in the size-age relationship would be highly relevant to the definition and diagnosis of primary sarcopenia. For example, diagnoses based on low relative muscle mass (as compared to reference populations of young adults) assume no sex difference in the general trajectory of age-related muscle size decline^3,55–58^, despite there being only one study^25^ to our knowledge that actually reports such a trend. The evidence scattered across the literature is suggestive of greater age-related muscle size decline in males^30,36,52–54,57,59–66^.

Muscle quality is defined as a measure of function (force exerted) per unit of muscle. Given the various ways in which both muscle function and size can be quantified, the literature on muscle quality is highly heterogenous. It is well established that age-related strength decline typically exceeds that attributable to size reduction alone^67^, indicating that reduced quality is a general feature of aged muscle. However, evidence of sexual dimorphism in age-related muscle quality decline is inconsistent, with studies variously reporting a greater decline in females^47,68^ or males^46,50^, or no sex difference^46,50,65,68^. Indeed, several studies provide evidence for multiple trends: for instance, Lynch et al. (1999) ^50^ detected no sex difference in age-related leg muscle quality decline, whereas a greater decline in arm muscle quality was evident in males. Similarly, Lindle et al. (1997) ^46^ reported similar rates of age-related muscle quality decline when strength was quantified using concentric peak torque, but greater male decline when eccentric peak torque measurements were used. It is therefore apparent that no consensus exists regarding the general relationship between age, sex, and muscle quality^48,69^.

Here we employ both cross-sectional and longitudinal study designs to characterise sex-specific ageing trajectories of muscle strength, size, and quality within the UK Biobank (UKB), a prospective cohort study of >500,000 UK participants aged 40–70 yr at recruitment in 2006–2010^70^, constituting >3% of the entire UK population within the age-range. We report a modest sex difference in age-related arm muscle strength decline, and demonstate that age-related arm muscle mass loss is considerably more pronounced in males, both in absolute and percentage terms. We also introduce two derived measures of muscle quality, measures of specific force that both exhibit substantially greater age-related decline in females. These trends hold across independent analyses of separate cross-sectional and longitudinal participant groups, and persist after accounting for systematic size differences between the sexes. Furthermore, sexual dimorphism is apparent irrespective of female menopause status or hormone replacement therapy (HRT) usage history, and is not explained by age-related changes in circulating levels of the sex hormones testosterone and oestradiol.

## Results

### Modest sexual dimorphism in age-related strength decline

We first performed a cross-sectional analysis of mean hand grip strength (HGS) readings taken at the baseline assessments of 368,336 UKB participants (202,028 females and 166,308 males) aged ≥40 and <70 years (‘Cross-sectional Participants’; Table 1; Supplementary Data 1). As expected, mean HGS of males (M) was greater than that of females (F) (by ∼68%; F 23.7 ± 0.01 kgf vs M 39.8 ± 0.02 kgf [mean ± standard error of the mean (SEM)]; *P* <0.001; False Positive Risk [FPR] <0.001), despite males being slightly older on average (Table 1). A multiple linear regression (MLR) model regressing HGS on age—with sex and an age-sex interaction term included as covariates (Methods)—indicated an annual linear decline of −0.26 kgf·yr^−1^ (−1.05%·yr^−1^) in females between 40–69 yr; the equivalent male decline was −0.30 kgf·yr^−1^ (−0.74%·yr^−1^), and this sex difference in magnitude of age-related decline (difference in slopes [β]: −0.043 ± 0.006 kgf·yr^−1^) proved to be statistically significant (*P* < 0.001; Supplementary Data 2) (note that when describing cross-sectional results, we use ‘decline’ and ‘trajectory’ to refer to age-related differences across the participant group as a whole, rather than change with age in individual participants). Although the fitted values from the MLR model accorded reasonably well with the observed data (Fig. 1a; Table 2), age-related male HGS decline clearly exhibited a gradually accelerating trajectory, rather than the linear trajectory assumed by the MLR model; in contrast, the female trajectory did not display a comparable acceleration beyond 60 yr (Fig. 1a), suggesting sexual dimorphism in age-related muscle strength decline.

**Figure 1.**
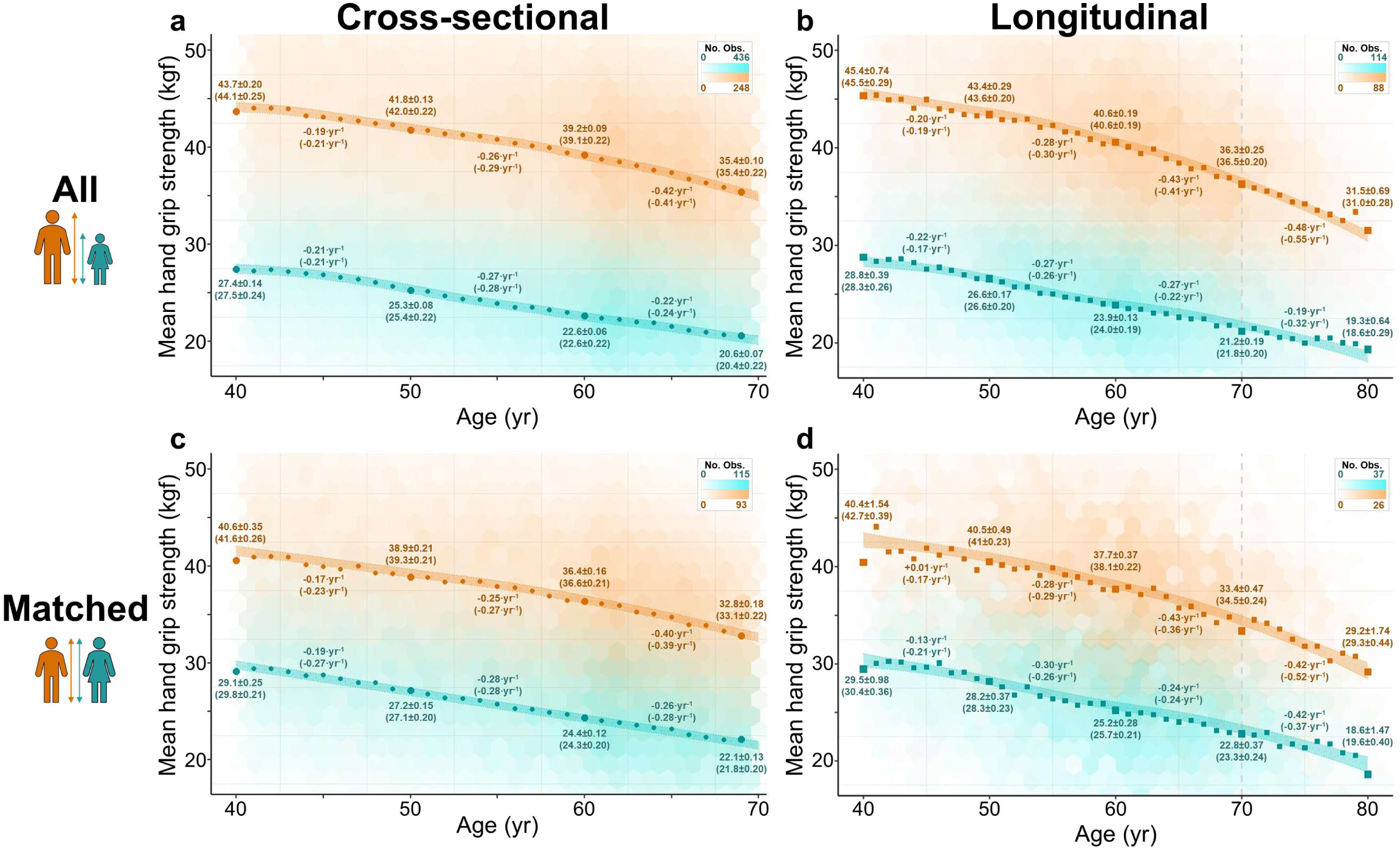
Modest sexual dimorphism in age-related strength decline. Sex-specific relationships between mean hand grip strength (HGS) and age for UK Biobank participants (females, teal; males, orange). **a** Cross-sectional participants (N = 368,336). **b** Longitudinal participants (N = 33,362). **c** Matched Cross-sectional Participants (N = 104,820). **d** Matched Longitudinal Participants (N = 7,958). Hexagonal bins: observed data, with greater opacity indicating higher density of participant measures (see scale bars in top right-hand corner of each panel). Coloured bands show 95% confidence interval for the HGS-age relationship fitted by a generalised additive mixed model (GAMM) (plotted with covariates fixed at ‘White’ for ethnicity and ‘Spring’ for season of assessment—Methods). Complete specifications for all models are provided in Supplementary Data 2, with fitted relationships in a and c based on HGS models 1 and 5, respectively. Fitted relationships in b and d are based on HGS models 2 and 6, respectively, that account for repeated measures (and therefore estimate true longitudinal change in HGS). Points represent observed mean values for each year age-group. Large points and values without parentheses are observed mean values (± SEM) at 40, 50, 60, and 69 yr (a,c) or 40, 50, 60, 70 and 80 yr (b,d), and inferred annual change (kgf·yr^−1^) over the intervening period. The equivalent fitted values are given within parentheses.

**Table 1.** Characteristics of five UK Biobank participant groups included in this study. Note that the Matched Cross-sectional Participants and Matched Longitudinal Participants are subsets of the Cross-sectional Participants and Longitudinal Participants, respectively, and that some participants are therefore included in both an unmatched and a matched group. In contrast, cross-sectional and longitudinal participant groups are completely independent, with no shared participants. Values for the longitudinal participant groups are from baseline assessment, with the exception of ‘Interval between assessments’. Menopause Participants were drawn from the Cross-sectional Participants group. ‘Pre-’ and ‘Post-’ refer to self-declared pre- and postmenopausal status, respectively. Data are either simple counts (‘N’ and ‘Ethnicity’), or mean ± the standard deviation. Equivalent values for the 23,930 participants excluded from this study (Methods) are available in Supplementary Data 1.\

|  | Cross-sectional Participants |  |  | Matched Cross-sectional Participants |  |  | Longitudinal Participants |  |  | Matched Longitudinal Participants |  |  | Menopause Participants |  |  |  |
| --- | --- | --- | --- | --- | --- | --- | --- | --- | --- | --- | --- | --- | --- | --- | --- | --- |
|  | All | Female | Male | All | Female | Male | All | Female | Male | All | Female | Male | All | Female (Pre-) | Female (Post-) | Male |
| N | 432,809 | 237,015 | 195,794 | 123,366 | 61,683 | 61,683 | 45,629 | 23,448 | 22,181 | 10,870 | 5,435 | 5,435 | 365,895 | 53,840 | 116,261 | 195,794 |
| Age (yr) | 57.04 ± 8.09 | 56.93 ± 7.99 | 57.19 ± 8.22 | 56.94 ± 8.15 | 56.94 ± 8.15 | 56.94 ± 8.15 | 55.67 ± 7.54 | 54.98 ± 7.38 | 56.39 ± 7.64 | 55.54 ± 7.46 | 55.54 ± 7.46 | 55.55 ± 7.46 | 56.74 ± 8.21 | 46.47 ± 3.8 | 60.73 ± 5.17 | 57.19 ± 8.22 |
| Interval between assessments (yr) | - | - | - | - | - | - | 8.92 ± 1.76 | 8.92 ± 1.75 | 8.91 ± 1.76 | 8.93 ± 1.75 | 8.92 ± 1.76 | 8.94 ± 1.75 | - | - | - | - |
| Standing height (cm) | 168.3 ± 9.3 | 162.4 ± 6.3 | 175.6 ± 6.9 | 168.9 ± 4.8 | 168.8 ± 4.7 | 168.9 ± 4.8 | 169.7 ± 9.2 | 163.3 ± 6.2 | 176.5 ± 6.6 | 169.7 ± 4.7 | 169.6 ± 4.6 | 169.7 ± 4.7 | 169.5 ± 9.3 | 164 ± 6.4 | 161.9 ± 6.2 | 175.6 ± 6.9 |
| Body mass (kg) | 78.2 ± 16.0 | 71.6 ± 14.1 | 86.1 ± 14.4 | 78.0 ± 12.6 | 78.0 ± 12.6 | 78.1 ± 12.6 | 77.0 ± 15.0 | 69.7 ± 12.8 | 84.8 ± 13.1 | 75.8 ± 10.0 | 75.7 ± 10.0 | 75.8 ± 10.0 | 79.1 ± 16.1 | 71.3 ± 14.8 | 71.0 ± 13.5 | 86.1 ± 14.4 |
| Ethnicity |  |  |  |  |  |  |  |  |  |  |  |  |  |  |  |  |
| Asian (% of group) | 10,095 (2.3) | 4,993 (2.1) | 5,102 (2.6) | 3,630 (3) | 323 (0.5) | 3,307 (5.3) | 625 (1.4) | 262 (1.1) | 363 (1.6) | 238 (2.2) | 19 (0.4) | 219 (4) | 8,851 (2.4) | 1,650 (3.1) | 2,099 (1.8) | 5,102 (2.6) |
| Black (% of group) | 7,215 (1.7) | 4,159 (1.8) | 3,056 (1.6) | 2,361 (1.9) | 1,085 (1.8) | 1,276 (2.1) | 296 (0.6) | 161 (0.7) | 135 (0.6) | 82 (0.8) | 39 (0.7) | 43 (0.8) | 5,936 (1.7) | 1,557 (2.9) | 1,323 (1.2) | 3,056 (1.6) |
| Other inc. Mixed and Unknown (% of group) | 8,262 (1.9) | 4,628 (1.9) | 3,634 (1.8) | 2,483 (2) | 833 (1.3) | 1,650 (2.7) | 548 (1.2) | 311 (1.3) | 237 (1.1) | 133 (1.2) | 50 (0.9) | 83 (1.5) | 7,022 (1.9) | 1,613 (3) | 1,775 (1.5) | 3,634 (1.8) |
| White (% of group) | 407,237 (94.1) | 223,235 (94.2) | 184,002 (94) | 114,892 (93.1) | 59,442 (96.4) | 55,450 (89.9) | 44,160 (96.8) | 22,714 (96.9) | 21,446 (96.7) | 10,417 (95.8) | 5,327 (98) | 5,090 (93.7) | 344,046 (94) | 49,020 (91) | 111,064 (95.5) | 184,002 (94) |

**Table 2.** Modest sexual dimorphism in age-related strength decline. Mean hand grip strength (HGS) summary data for four UK Biobank participant groups included in this study, showing mean for the entire cohort, at specific ages, and the differences and rates of decline in both absolute force and percentage terms. Observed (‘Obs.’; mean ± SEM [N]) and generalised additive mixed model (GAMM)-fitted (‘Fitt.’; fitted value ± standard error of the fitted value) values are given. ‘Inferred declines’ are derived from the difference in values at 40 vs 69 yr (cross-sectional participant groups) or 40 vs 80 yr (longitudinal participant groups), and are different from the multiple linear regression (MLR) model-derived annual declines detailed elsewhere (Results; Methods; Supplementary Data 2). Note that fitted values assume a White participant assessed during Spring (Methods), and that N varies in the matched cohort single year groups due to the matching criterion for age at baseline assessment being ± 6 months.

|  | Cross-sectional Participants |  |  |  | Matched Cross-sectional Participants |  |  |  |
| --- | --- | --- | --- | --- | --- | --- | --- | --- |
|  | Females |  | Males |  | Females |  | Males |  |
| HGS | Obs. | Fitt. | Obs. | Fitt. | Obs. | Fitt. | Obs. | Fitt. |
| All participants [kgf]<br>(N) | 23.7 ± 0.01<br>(202,028) | - | 39.8 ± 0.02<br>(166,308) | - | 25.4 ± 0.03<br>(52,723) | - | 37.1 ± 0.04<br>(52,097) | - |
| 40 yr [kgf]<br>(N) | 27.4 ± 0.14<br>(1,969) | 27.5 ± 0.24 | 43.7 ± 0.20<br>(1,934) | 44.1 ± 0.25 | 29.1 ± 0.25<br>(571) | 29.8 ± 0.21 | 40.6 ± 0.35<br>(594) | 41.6 ± 0.26 |
| 69 yr [kgf]<br>(N) | 20.6 ± 0.07<br>(6,251) | 20.4 ± 0.22 | 35.4 ± 0.10<br>(6,080) | 35.4 ± 0.22 | 22.1 ± 0.13<br>(1,775) | 21.8 ± 0.20 | 32.8 ± 0.18<br>(1,755) | 33.1 ± 0.22 |
| Difference 40 vs 69 yr [kgf] | -6.82 | -7.10 | -8.24 | -8.70 | -7.03 | -8.00 | -7.76 | -8.50 |
| Inferred decline 40–69 yr [kgf·yr<sup>-1</sup>] | -0.24 | -0.24 | -0.28 | -0.30 | -0.24 | -0.28 | -0.27 | -0.29 |
| Difference 40 vs 69 yr [%] | -24.9 | -25.8 | -18.9 | -19.7 | -24.1 | -26.80 | -19.10 | -20.40 |
| Inferred decline 40–69 yr [%·yr<sup>-1</sup>] | -0.98 | -1.02 | -0.72 | -0.75 | -0.95 | -1.07 | -0.73 | -0.79 |
|  | Longitudinal Participants |  |  |  | Matched Longitudinal Participants |  |  |  |
|  | Females |  | Males |  | Females |  | Males |  |
| HGS | Obs. | Fitt. | Obs. | Fitt. | Obs. | Fitt. | Obs. | Fitt. |
| 40 yr [kgf]<br>(N) | 28.8 ± 0.39<br>(211) | 28.3 ± 0.26 | 45.4 ± 0.74<br>(156) | 45.5 ± 0.29 | 29.5 ± 0.98<br>(42) | 30.4 ± 0.36 | 40.4 ± 1.54<br>(38) | 42.7 ± 0.39 |
| 80 yr [kgf]<br>(N) | 19.3 ± 0.64<br>(70) | 18.6 ± 0.29 | 31.6 ± 0.69<br>(119) | 31.0 ± 0.3 | 18.6 ± 1.47<br>(16) | 19.6 ± 0.4 | 29.2 ± 1.74<br>(18) | 29.3 ± 0.44 |
| Difference 40 vs 80 yr [kgf] | -9.45 | -9.7 | -13.8 | -14.5 | -10.9 | -10.8 | -11.3 | -13.4 |
| Inferred decline 40–80 yr [kgf·yr<sup>-1</sup>] | -0.24 | -0.24 | -0.35 | -0.36 | -0.27 | -0.27 | -0.28 | -0.34 |
| Difference 40 vs 80 yr [%] | -32.8 | -34.3 | -30.5 | -31.9 | -36.9 | -35.5 | -27.9 | -31.4 |
| Inferred decline 40–80 yr [%·yr<sup>-1</sup>] | -0.99 | -1.04 | -0.9 | -0.95 | -1.14 | -1.09 | -0.81 | -0.94 |

To quantify the apparent sexual dimorphism more rigorously, we next fitted a generalised additive mixed model (GAMM) on all Cross-sectional Participants, this time regressing HGS on a global age spline and sex, as well as including ethnicity and potential technical confounders as additional covariates (Methods). Including sex-specific age splines as an additional model term substantially improved model fit compared with the global age spline alone (Akaike weight-derived percentage probability that including sex-specific splines improved model fit [‘prob_AW_’] >99.9%—HGS model 1 in Supplementary Data 2; Methods), thereby confirming sexual dimorphism in the HGS-age trajectory in the Cross-sectional Participants. The general trends identified in the observed data and MLR model analysis were supported by the sex-specific age splines GAMM, with fitted values at 40 vs 69 yr equivalent to annual HGS decline of −0.24 kgf·yr^−1^ (−1.02 %·yr^−1^) in females, and −0.30 kgf·yr^−1^ (−0.75%·yr^−1^) in males (Fig. 1a; Table 2). Therefore, coss-sectional analysis indicates that males lose more absolute strength—but less proportional strength—than females between 40–70 yr.

Cross-sectional and longitudinal studies can produce markedly different results when used to assess age-related anthropometric change, due to phenomena such as birth cohort effects^37,38,40^. We therefore investigated the relationship between HGS and age in an entirely separate group of 45,629 UKB participants (‘Longitudinal Participants’; Table 1; Supplementary Data 1) who attended an additional (‘repeat’) assessment 3.83–12.83 years after their baseline assessment. 33,362 Longitudinal Participants had suitable HGS readings taken at both assessments (17,146 females and 16,216 males). MLR model analysis of baseline and repeat assessment measures in each of the Longitudinal Participants returned fitted declines between 40–80 yr of −0.25 kgf·yr^−1^ in females (−1.04 %·yr^−1^) and −0.34 kgf·yr^−1^ in males (−0.85 %·yr^−1^) (β: −0.090 ± 0.011 kgf·yr^−1^; *P* < 0.001). The GAMM-fitted mean HGS value for females at 80 yr (18.6 ± 0.29 kgf) was 9.7 kgf (−34.3%) lower than at 40 yr (28.3 ± 0.26 kgf), corresponding to a linear age-related decline of −0.24 kgf·yr^−1^ (−1.04%·yr^−1^), whereas the fitted value for males at 80 yr (31.0 ± 0.30 kgf) was 14.5 kgf (−31.9%) lower than at 40 yr (45.5 ± 0.29 kgf), equivalent to a linear age-related change of −0.36 kgf·yr^−1^ (−0.95%·yr^−1^). Because these analyses did not explicitly link the repeated measures on each individual, they were effectively a second cross-sectional analysis on a non-overlapping group of UKB participants. As with the Cross-sectional Participants, the HGS-age trajectories of female and male Longitudinal Participants appeared similar until ∼60 yr, after which an accelerating rate of HGS decline was apparent in males only, although females showed a potential acceleration in decline beyond 70 yr (Fig. 1b).

We next implemented a GAMM on the Longitudinal Participants that explicitly accounted for repeated measures. This formal longitudinal analysis confirmed the sexual dimorphism (prob_AW_ >99.9%—HGS model 2 in Supplementary Data 2), with the trajectories of the fitted declines between 40–80 yr (F −0.24 kgf·yr^−1^ vs M −0.36 kgf·yr^−1^; Fig. 1b) similar to those presented in both the observed Longitudinal Participant data and the analysis of Cross-sectional Participants. Thus, the results of longitudinal analyses broadly replicated the cross-sectional findings, thereby confirming sexual dimorphism in the HGS-age relationship within the overall UKB cohort.

HGS is positively and strongly correlated with body size^71^, but none of the above results account for body-size differences, including systematic sex differences in body size. We therefore refitted the cross-sectional and longitudinal GAMMs with both global and sex-specific splines for the covariates standing height and body mass (Methods). The same general sexual dimorphism in trajectories was observed in both models (each prob_AW_ >99.9%—HGS models 3–4 in Supplementary Data 2; Supplementary Fig. 1). As a further control for body size, we also analysed two age- and size-matched subsets: 61,683 female:male pairs drawn from the Cross-sectional Participants (‘Matched Cross-sectional Participants’), and 5,435 female:male pairs drawn from the Longitudinal Participants (‘Matched Longitudinal Participants’) (Table 1; Methods). Unsurprisingly, males remained substantially stronger than females after matching (e.g. ∼46% in the Matched Cross-sectional Participants; F 25.35 ± 0.03 kgf vs M 37.10 ± 0.04 kgf; *P* <0.001; FPR <0.001). GAMM analysis confirmed sexual dimorphism in age-related HGS decline in both matched subsets (each prob_AW_ >99.9%—HGS models 5–6 in Supplementary Data 2), and the trajectories were broadly comparable to those of the unmatched participant groups, with an acceleration between 60–70 yr in males that was not observed in females, although, as with the Longitudinal Participants, female Matched Longitudinal Participants showed some evidence of accelerating decline beyond 70 yr (Figs 1a–d). Interestingly, MLR model analysis of Matched Cross-sectional Participants did not support a sex difference in the HGS-age relationship (β: −0.006 ± 0.010 kgf·yr^−1^; *P* = 0.29), thereby indicating a sex difference in trajectory shape rather than absolute rate of decline within this participant group. In all other analyses, age-related HGS decline across the assessed age range unequivocally differed by sex, being greater in males in terms of kilogram-force, and greater in proportional terms in females. This held true across both the observed data and the model-fitted relationships (Figs 1a–d; Table 2; Supplementary Data 2).

### Greater age-related muscle mass decline in males

Novel estimates of mean arm muscle mass (MM) (Fig. 2; Table 3) were generated using a modified version of an equation from Powell et al. (2020) ^72^, which combines impedance values (ohms—Ω) from Bioelectrical Impedance Analysis (BIA) with age, sex, and anthropometric data (e.g. standing height and body mass). These novel estimates were validated against dual x-ray absorptiometry (DXA) scan data, and were shown to be significantly more accurate than the standard arm muscle mass estimates available in the UKB dataset (Methods).

**Figure 2.**
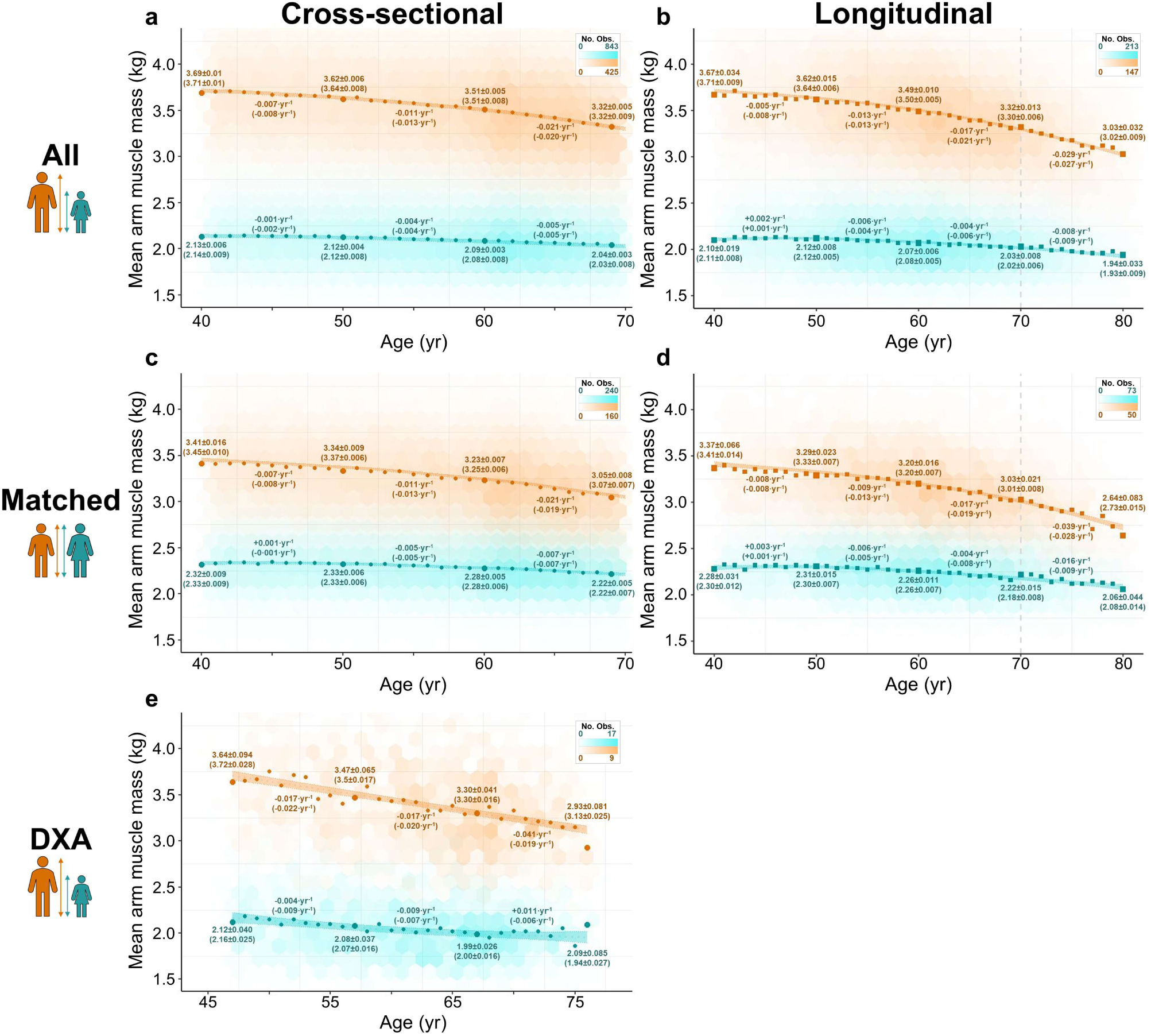
Age-related muscle mass decline is much greater in males than females. Sex-specific relationships between mean arm muscle mass (MM) and age for UK Biobank participants (females, teal; males, orange). **a** Cross-sectional participants (N = 432,809). **b** Longitudinal participants (N = 44,609). **c** Matched Cross-sectional Participants (N = 123,366). **d** Matched Longitudinal Participants (N = 10,601). **e** Dual x-ray absorptiometry (DXA)-derived MM estimates for a subset of Longitudinal Participants ≥44.5 and ≤78 yr (one observation per participant) (N = 4,253). Hexagonal bins: observed data, with greater opacity indicating higher density of participant measures (see scale bars in top right-hand corner of each panel). Coloured bands show 95% confidence interval for the MM-age relationship fitted by a generalised additive mixed model (GAMM) (plotted with covariates fixed at ‘White’ for ethnicity and ‘Spring’ for season of assessment—Methods). Complete specifications for all models are provided in Supplementary Data 2, with fitted relationships in a, c, and e based on MM models 1, 5, and 7 respectively. Fitted relationships in b and d are based on MM models 2 and 6, respectively, that account for repeated measures (and therefore estimate true longitudinal change in MM). Points represent observed mean values for each year age-group. Large points and values without parentheses are observed mean values (± SEM) at 40, 50, 60, and 69 yr (a,c), 40, 50, 60, 70 and 80 yr (b,d), or 47, 57, 67, and 76 yr (e), and inferred annual change (kg·yr^−1^) over the intervening period. The equivalent fitted values are given within parentheses.

**Table 3.** Age-related muscle mass decline is much greater in males than females. Mean arm muscle mass (MM) summary data for four UK Biobank participant groups included in this study, showing mean for the entire cohort, at specific ages, and the differences and rates of decline in both absolute force and percentage terms. Observed (‘Obs.’; mean ± SEM [N]) and generalised additive mixed model (GAMM)-fitted (‘Fitt.’; fitted value ± standard error of the fitted value) values are given. ‘Inferred declines’ are derived from the difference in values at 40 vs 69 yr (cross-sectional participant groups) or 40 vs 80 yr (longitudinal participant groups), and are different from the multiple linear regression (MLR) model-derived annual declines detailed elsewhere (Results; Methods; Supplementary Data 2). Note that fitted values assume a White participant assessed during Spring (Methods), and that N varies in the matched cohort single year groups due to the matching criterion for age at baseline assessment being ± 6 months.

|  | Cross-sectional Participants |  |  |  | Matched Cross-sectional Participants |  |  |  |
| --- | --- | --- | --- | --- | --- | --- | --- | --- |
|  | Females |  | Males |  | Females |  | Males |  |
| MM | Obs. | Fitt. | Obs. | Fitt. | Obs. | Fitt. | Obs. | Fitt. |
| All participants [kg] | 2.10 ± 0.001 | - | 3.53 ± 0.001 | - | 2.29 ± 0.001 | - | 3.26 ± 0.002 | - |
| (N) | (237,015) |  | (195,794) |  | (61,683) |  | (61,683) |  |
| 40 yr [kg] | 2.13 ± 0.01 | 2.14 ± 0.01 | 3.69 ± 0.01 | 3.71 ± 0.01 | 2.32 ± 0.01 | 2.33 ± 0.01 | 3.41 ± 0.02 | 3.45 ± 0.01 |
| (N) | (2,433) |  | (2,346) |  | (687) |  | (723) |  |
| 69 yr [kg] | 2.04 ± 0.003 | 2.03 ± 0.01 | 3.32 ± 0.01 | 3.32 ± 0.01 | 2.22 ± 0.01 | 2.22 ± 0.01 | 3.05 ± 0.01 | 3.07 ± 0.01 |
| (N) | (7,310) |  | (7,090) |  | (2,067) |  | (2,065) |  |
| Difference 40 vs 69 yr [kg] | -0.09 | -0.11 | -0.37 | -0.39 | -0.10 | -0.11 | -0.37 | -0.38 |
| Inferred decline 40–69 yr [kg·yr<sup>-1</sup>] | -0.003 | -0.004 | -0.013 | -0.013 | -0.003 | -0.004 | -0.013 | -0.013 |
| Difference 40 vs 69 yr [%] | -4.27 | -5.00 | -10.00 | -10.60 | -4.36 | -4.84 | -10.75 | -11.10 |
| Inferred decline 40–69 yr [%·yr<sup>-1</sup>] | -0.15 | -0.18 | -0.36 | -0.38 | -0.15 | -0.17 | -0.39 | -0.40 |
|  | Longitudinal Participants |  |  |  | Matched Longitudinal Participants |  |  |  |
|  | Females |  | Males |  | Females |  | Males |  |
| MM | Obs. | Fitt. | Obs. | Fitt. | Obs. | Fitt. | Obs. | Fitt. |
| 40 yr [kg] | 2.10 ± 0.02 | 2.11 ± 0.01 | 3.67 ± 0.03 | 3.71 ± 0.01 | 2.28 ± 0.03 | 2.30 ± 0.01 | 3.37 ± 0.07 | 3.41 ± 0.01 |
| (N) | (230) |  | (170) |  | (47) |  | (39) |  |
| 80 yr [kg] | 1.94 ± 0.03 | 1.93 ± 0.01 | 3.03 ± 0.03 | 3.02 ± 0.01 | 2.06 ± 0.04 | 2.08 ± 0.01 | 2.64 ± 0.08 | 2.73 ± 0.02 |
| (N) | (84) |  | (147) |  | (20) |  | (19) |  |
| Difference 40 vs 80 yr [kg] | -0.15 | -0.18 | -0.64 | -0.69 | -0.22 | -0.21 | -0.73 | -0.68 |
| Inferred decline 40–80 yr [kg·yr<sup>-1</sup>] | -0.004 | -0.004 | -0.016 | -0.017 | -0.005 | -0.005 | -0.018 | -0.017 |
| Difference 40 vs 80 yr [%] | -7.31 | -8.50 | -17.50 | -18.60 | -9.48 | -9.23 | -21.80 | -20.00 |
| Inferred decline 40–80 yr [%·yr<sup>-1</sup>] | -0.19 | -0.22 | -0.48 | -0.51 | -0.25 | -0.24 | -0.61 | -0.56 |

MM of male Cross-sectional Participants was ∼68% greater than that of females (F 2.10 ± 0.001 kg vs M 3.53 ± 0.001 kg; *P* <0.001; FPR <0.001), and remained ∼42% greater in the Matched Cross-sectional Participants (F 2.29 ± 0.001 kg vs M 3.26 ± 0.002 kg; *P* <0.001; FPR <0.001), confirming the expected substantial sex difference in muscularity not attributable to differences in body size (Figs 2a,c). Linear rates of age-related MM decline determined by MLR models were over three times faster in males than females, at −0.004 kg·yr^−1^ (F) vs −0.013 kg·yr^−1^ (M) in Cross-sectional Participants between 40–69 yr, and −0.005 kg·yr^−1^ (F) vs −0.016 kg·yr^−1^ (M) in Longitudinal Participants between 40–80 yr. These declines were also over twice as great in males in percentage terms, at −0.18%·yr^−1^ (F) vs −0.38%·yr^−1^ (M) in the Cross-sectional Participants (40–69 yr), and −0.22%·yr^−1^ (F) vs −0.46%·yr^−1^ (M) in the Longitudinal Participants (40–80 yr) (Supplementary Data 2). Consequently, the magnitude of the observed sex difference in muscularity was found to decrease with age: for instance, observed MM of male Longitudinal Participants was ∼75% greater than that of their female counterparts at 40 yr, whereas the difference was ∼56% at 80 yr (Fig. 2b; Table 3).

Sexual dimorphism in the MM-age relationship was strongly supported by GAMMs fitted to all matched and unmatched participants groups, irrespective of whether standing height and body mass were included as covariates (each prob_AW_ >99.9%—MM models 1–6 in Supplementary Data 2). GAMM-fitted values were in close accordance with the observed data, with fitted male declines again over three times as great as those of females in absolute terms, and over twice as great in percentage terms (Fig. 2; Table 3; Supplementary Fig. 2). Importantly, the longitudinal results closely replicated those observed in the cross-sectional data (Fig. 2b,d), and analysis of body-size matched participant groups produced similar results to analyses of unmatched participants (Fig. 2c,d). Thus, the key finding of substantially greater age-related muscle mass decline in males was consistent across the Cross-sectional Participants, Matched Cross-sectional Participants, Longitudinal Participants, and Matched Longitudinal Participants.

Substantially greater male muscle mass decline with age was also evident in DXA scan data available for 4,253 Longitudinal Participants (Fig. 2e; prob_AW_ >99.9%—MM model 7 in Supplementary Data 2), and across three alternative sets of BIA-derived muscle mass estimates (Supplementary Fig. 3). Intriguingly, models fitted to both BIA- and DXA-derived muscle mass estimates imply that female age-related muscle mass loss essentially reflects general age-related body-size decline, whereas male age-related muscle mass decline far exceeds that attributable to body-size decline alone (Supplementary Fig. 4), further emphasising the existence of a striking sexual dimorphism in the mass-age relationship.

### Greater age-related muscle quality decline in females

Regression of measured HGS (a functional output) on MM (an intrinsic property of muscle) revealed that much of the variance in HGS was not explained by variance in MM, thereby suggesting differences in muscle ‘quality’ between individuals. A sex difference was also evident, with a stronger positive correlation between HGS and MM apparent in males than females (Supplementary Fig. 5). These initial findings prompted us to develop muscle quality measures with which to investigate potential sexual dimorphism in age-related muscle quality change. Because isometric force production—as measured in HGS—is biomechanically proportional to the cross-sectional area (CSA) of active muscle rather than its mass^44,73^, we first derived muscle quality values by combining the HGS and MM observations with individual estimates of forearm (ulna) length to estimate forearm muscle specific force (i.e. force·CSA^−1^) (MQ—Methods).

Age-related MQ decline was substantially greater in females than males across all participant groups and analysis methods (Fig. 3; Table 4; Supplementary Data 2). The MLR model-derived linearised rate of female MQ decline between 40–69 yr was around twice that of males when assessed cross-sectionally (Cross-sectional Participants: F −1.03%·yr^−1^ vs M −0.45%·yr^−1^) and after body-size matching (Matched Cross-sectional Participants: F −0.98%·yr^−1^ vs M −0.47%·yr^−1^). This sex difference was similarly pronounced between 40–80 yr in both the Longitudinal Participants (F −0.91%·yr^−1^ vs M −0.44%·yr^−1^) and Matched Longitudinal Participants (F −0.88%·yr^−1^ vs M −0.43%·yr^−1^) (Supplementary Data 2). Sexual dimorphism in ageing trajectories was strongly supported by GAMMs fitted to all participant groups, including those in which standing height and body mass were included as covariates (each prob_AW_ >99.9%—MQ models 1–6 in Supplementary Data 2; Supplementary Fig. 6), with fitted relationships closely resembling the observed MQ-age trajectories (Fig. 3; Table 4; Supplementary Data 2). Analysis of the DXA-scanned Longitudinal Participants (N = 3,972— 2,061 females and 1,911 males) provided additional support for sex-specific muscle quality trajectories (Fig. 3e; prob_AW_ >99.9%—MQ model 7 in Supplementary Data 2), as the decline in muscle quality between 47–76 yr was over twice as great in females as in males in this participant group (observed: F −1.44%·yr^−1^ vs M −0.66%·yr^−1^; GAMM-fitted: F −1.10%·yr^−1^ vs M −0.50%·yr^−1^; Fig. 3e).

**Figure 3.**
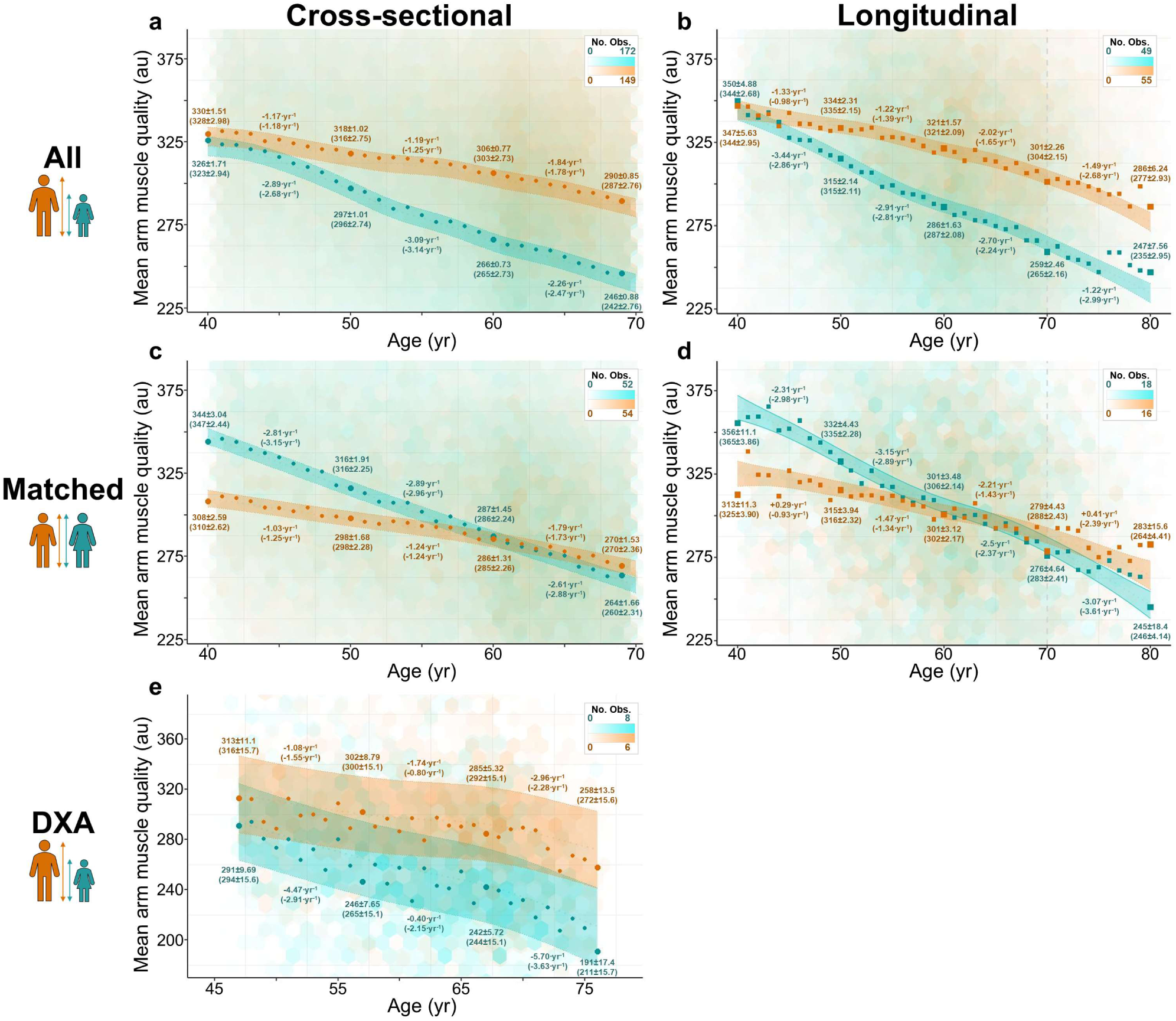
Age-related muscle quality decline is substantially greater in females than males. Sex-specific relationships between mean arm muscle quality (MQ) and age for UK Biobank participants (females, teal; males, orange). **a** Cross-sectional participants (N = 368,336). **b** Longitudinal participants (N = 32,663). **c** Matched Cross-sectional Participants (N = 104,820). **d** Matched Longitudinal Participants (N = 7,778). **e** Dual x-ray absorptiometry (DXA)-derived MQ estimates for a subset of Longitudinal Participants ≥44.5 and ≤78 yr (one observation per participant) (N = 3,972). Hexagonal bins: observed data, with greater opacity indicating higher density of participant measures (see scale bars in top right-hand corner of each panel). Coloured bands show 95% confidence interval for the MQ-age relationship fitted by a generalised additive mixed model (GAMM) (plotted with covariates fixed at ‘White’ for ethnicity and ‘Spring’ for season of assessment—Methods). Complete specifications for all models are provided in Supplementary Data 2, with fitted relationships in a, c, and e based on MQ models 1, 5, and 7 respectively. Fitted relationships in b and d are based on MQ models 2 and 6, respectively, that account for repeated measures (and therefore estimate true longitudinal change in MQ). Points represent observed mean values for each year age-group. Large points and values without parentheses are observed mean values (± SEM) at 40, 50, 60, and 69 yr (a,c), 40, 50, 60, 70 and 80 yr (b,d), or 47, 57, 67, and 76 yr (e), and inferred annual change (in arbitrary units [au]) (au·yr^−1^) over the intervening period. The equivalent fitted values are given within parentheses.

**Table 4.** Age-related muscle quality decline is substantially greater in females than males. Mean arm muscle quality (MQ) summary data for four UK Biobank participant groups included in this study, showing mean for the entire cohort, at specific ages, and the differences and rates of decline in both absolute force and percentage terms. Observed (‘Obs.’; mean ± SEM [N]) and generalised additive mixed model (GAMM)-fitted (‘Fitt.’; fitted value ± standard error of the fitted value) values are given. ‘Inferred declines’ are derived from the difference in values at 40 vs 69 yr (cross-sectional participant groups) or 40 vs 80 yr (longitudinal participant groups), and are different from the multiple linear regression (MLR) model-derived annual declines detailed elsewhere (Results; Methods; Supplementary Data 2). Note that fitted values assume a White participant assessed during Spring (Methods), and that N varies in the matched cohort single year groups due to the matching criterion for age at baseline assessment being ± 6 months. The discrepancy between observed and GAMM-fitted values in the male Matched Longitudinal Participants apparently reflects the small sample sizes amongst the very oldest participants in this participant group (e.g. 18 males at 80 yr).

|  | Cross-sectional Participants |  |  |  | Matched Cross-sectional Participants |  |  |  |
| --- | --- | --- | --- | --- | --- | --- | --- | --- |
|  | Females |  | Males |  | Females |  | Males |  |
| MQ | Obs. | Fitt. | Obs. | Fitt. | Obs. | Fitt. | Obs. | Fitt. |
| All participants [au]<br>(N) | 279 ± 0.2<br>(202,028) | - | 310 ± 0.2<br>(166,308) | - | 298 ± 0.3<br>(52,723) | - | 290 ± 0.3<br>(52,097) | - |
| 40 yr [au]<br>(N) | 326 ± 1.7<br>(1,969) | 323 ± 2.9 | 330 ± 1.5<br>(1,934) | 328 ± 3.0 | 344 ± 3.0<br>(571) | 347 ± 2.4 | 308 ± 2.6<br>(594) | 310 ± 2.6 |
| 69 yr [au]<br>(N) | 246 ± 0.9<br>(6,251) | 242 ± 2.8 | 290 ± 0.9<br>(6,080) | 287 ± 2.8 | 264 ± 1.7<br>(1,775) | 260 ± 2.3 | 270 ± 1.5<br>(1,755) | 270 ± 2.4 |
| Difference 40 vs 69 yr (au) | -80.1 | -80.4 | -40.2 | -40.3 | -80.5 | -87.0 | -38.8 | -40.5 |
| Inferred decline 40–69 yr [au·yr<sup>-1</sup>] | -2.76 | -2.77 | -1.39 | -1.39 | -2.78 | -3.00 | -1.34 | -1.40 |
| Difference 40 vs 69 yr [%] | -24.6 | -24.9 | -12.2 | -12.3 | -23.4 | -25.1 | -12.6 | -13.1 |
| Inferred decline 40–69 [%·yr<sup>-1</sup>] | -0.97 | -0.98 | -0.45 | -0.45 | -0.91 | -0.99 | -0.46 | -0.48 |
|  | Longitudinal Participants |  |  |  | Matched Longitudinal Participants |  |  |  |
|  | Females |  | Males |  | Females |  | Males |  |
| MQ | Obs. | Fitt. | Obs. | Fitt. | Obs. | Fitt. | Obs. | Fitt. |
| 40 yr [au]<br>(N) | 350 ± 4.9<br>(211) | 344 ± 2.78 | 347 ± 5.6<br>(156) | 344 ± 3.0 | 355 ± 11.1<br>(42) | 365 ± 3.7 | 313 ± 11.3<br>(38) | 325 ± 3.9 |
| 80 yr [au]<br>(N) | 247 ± 7.6<br>(68) | 235 ± 3.0 | 286 ± 6.2<br>(116) | 277 ± 2.9 | 245 ± 18.4<br>(16) | 246 ± 4.1 | 283 ± 15.6<br>(18) | 264 ± 4.4 |
| Difference 40 vs 80 yr [au] | -103.0 | -109.0 | -60.6 | -67.0 | -110.0 | -119.0 | -29.8 | -60.9 |
| Inferred decline 40–80 yr [au·yr<sup>-1</sup>] | -2.57 | -2.73 | -1.51 | -1.68 | -2.76 | -2.96 | -0.74 | -1.52 |
| Difference 40 vs 80 yr [%] | -29.40 | -31.70 | -17.50 | -9.50 | -31.00 | -32.5 | -9.52 | -18.70 |
| Inferred decline 40–80 yr [%·yr<sup>-1</sup>] | -0.87 | -0.95 | -0.48 | -0.54 | -0.92 | -0.98 | -0.25 | -0.52 |

Observed MQ of females and males was almost identical at 40 yr in both the Cross-sectional Participants and the Longitudinal Participants (Fig. 3a,b). In contrast, average female MQ exceeded that of males until ∼60 yr in the matched participant groups (Fig. 3c,d), and also when systematic sex differences in body size were accounted for within GAMMs (Supplementary Fig. 6). This distinction was explained by a positive correlation between MQ and standing height in both sexes. The greater mean standing height of males compensated for their lower MQ in the unmatched participant groups, whereas the elimination of this sex difference in the matched participant groups caused female MQ to appear relatively greater (Supplementary Fig. 7). These complex allometric relationships between standing height, CSA, MM, and MQ suggest that changes in muscle architecture with age contribute to the observed sexual dimorphism.

An important aspect of muscle architecture is pennation angle, which is rarely exactly 0° (as implictly assumed in our MQ calculation), and is typically between 2–17° in forearm muscle^74,75^. We therefore assessed age-related differences in a second measure of muscle quality (MQ’), defined as *HGS/MM*, which can be thought of as an alternative extreme muscle quality measure assuming highly pennate muscles. The similar trends observed in MQ’ (each prob_AW_ >99.9%—MQ’ models 1–5 in Supplementary Data 2; Supplementary Fig. 8) reinforce our conclusion that age-related muscle quality decline is greater in females than males.

### Menopause, HRT, and sex hormones do not explain sex differences

To investigate the relationship between menopause status and muscle ageing, we used GAMMs regressing HGS, MM, and MQ on age to perform pairwise comparisons on groups of 53,840 premenopausal females, 116,261 postmenopausal females, and 195,794 males drawn from the Cross-sectional Participants (‘Menopause Participants’, N = 365,895; Methods; Table 1; Supplementary Data 1; Supplementary Fig. 9). GAMMs also included self-reported ethnicity, technical covariates, standing height, and body mass. Models were fitted between 45–60 yr, the window within which menopause typically occurs^76^. Perimenopause was not included in these analyses because perimenopausal individuals cannot reliably be identified in the UKB.

GAMM comparison supported distinct HGS-age trajectories for pre- and postmenopausal females (prob_AW_ 98.7%; HGS menopause model 1 in Supp. Info 2), and strongly supported distinct trajectories between males and both female groups (each prob_AW_ >99.9%; HGS menopause models 2 and 3 in Supplementary Data 2) (Fig. 4a). Age-related decline in female MM was minimal (Fig. 2), and there was no evidence of a difference in MM-age trajectory between pre- and postmenopausal females (prob_AW_ 54.4%; MM menopause model 1 in Supplementary Data 2). However, postmenopausal status was associated with significantly lower MM than premenopausal status (∼0.037 kg lower MM, as estimated without group-specific age splines; *P* <0.001; FPR <0.001; MM menopause model 1_GLOBAL_ in Supplementary Data 2). The MM-age trajectories of both female groups were distinct from that of males (each prob_AW_ >99.9%; MM menopause models 2 and 3 in Supplementary Data 2) (Fig. 4b). The MQ-age trajectory of males was also distinct from that of both female groups (each prob_AW_ >99.9%; MQ menopause models 2 and 3 in Supplementary Data 2) (Fig. 4c). There was weak support for distinct MQ-age trajectories between pre- and postmenopausal females (prob_AW_ 92.1%; MQ menopause model 1 in Supplementary Data 2), indicating that any difference in the HGS-age trajectory between pre- and postmenopausal females is attributable to MQ differences between these groups.

**Figure 4.**
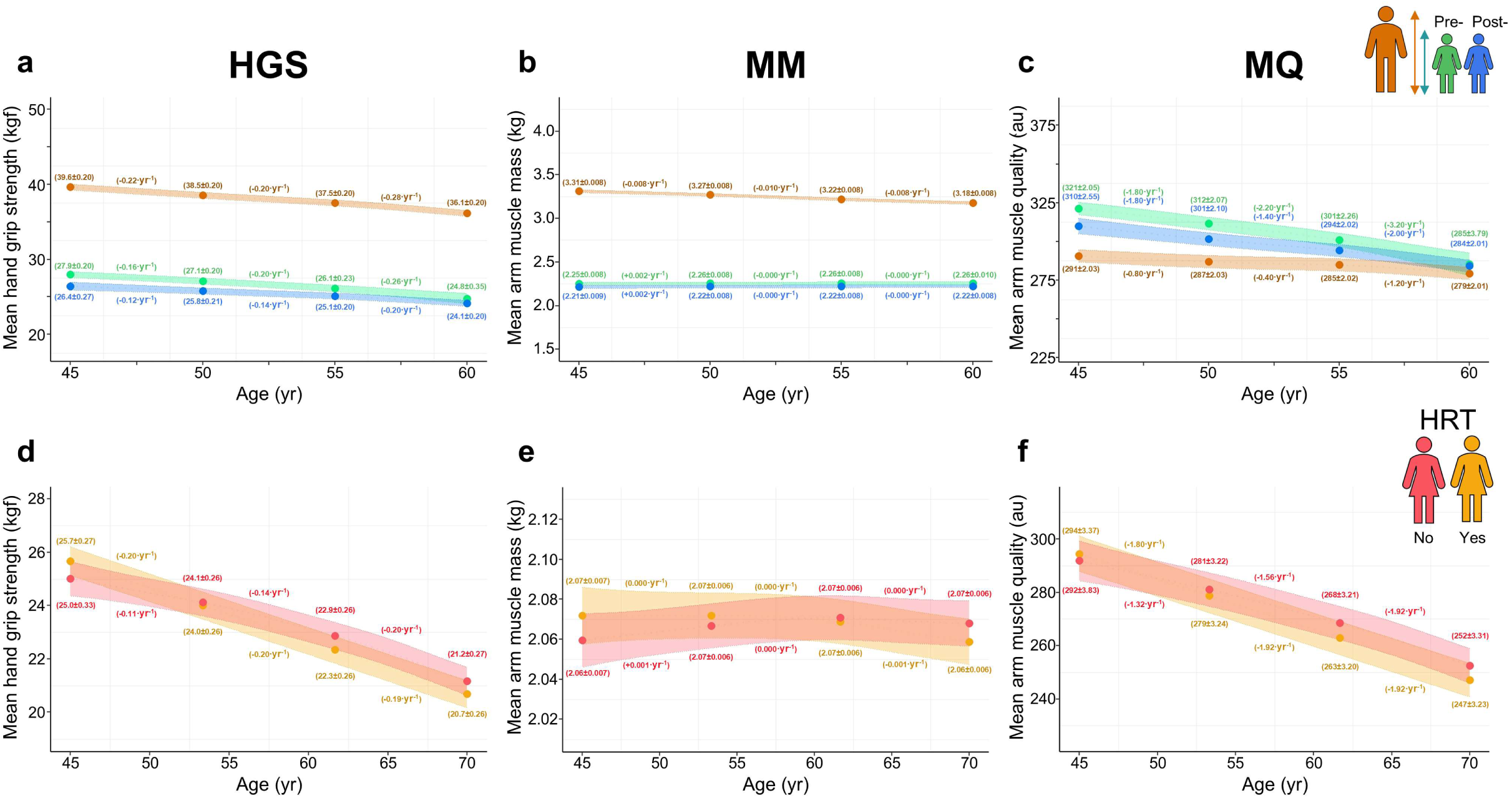
Females exhibit distinct muscle ageing trajectories from males irrespective of menopause status and history of HRT use. Generalised additive mixed model (GAMM)-fitted relationships between age and mean hand grip strength (HGS) (a,d), mean arm muscle mass (MM) (b,e), and mean arm muscle quality (MQ) (c,f) in the Menopause Participants, a subset of Cross-sectional Participants (Supplementary Data 1). Groups were not matched on body size, but standing height and body mass were included as covariates. Coloured bands show 95% confidence interval (CI) for the fitted phenotype-age relationship (plotted with covariates fixed at ‘White’ for ethnicity and ‘Spring’ for season of assessment—Methods). Complete specifications for all models are provided in Supplementary Data 2. Large points and values in parentheses are fitted values (± SEM) at 45, 50, 55, and 60 yr (a–c), or 45, 53.33, 61.66, and 70 yr (d–f), and inferred annual change (unit·yr^−1^) over the intervening period. **a–c** Females are stratified by menopause status (premenopausal females, green; postmenopausal females, blue; males, orange). Standing height and body mass are fixed for plotting at 167.15 cm and 76.13 kg, respectively—calculated as *(mean premenopausal female value + mean postmenopausal female value + mean male value)/3*. Note that the models plotted here were fitted to all three groups, and thus differ slightly from the pairwise models presented in the Results. **d–f** 115,990 postmenopausal Menopause Participants, stratified by response to the question “Have you ever used hormone replacement therapy (HRT)?”. Pink and gold represent participants who answered “No” (N = 66,291) and “Yes” (N = 49,699), respectively. Standing height and body mass are fixed for plotting at 161.90 cm and 70.99 kg, respectively—calculated as *(mean “Yes” value + mean “No” value)/2*. Despite there being some statistical support for distinct muscle ageing trajectories for these two groups, the widely overlapping CIs indicate that these differences are minor, at least in the case of a White female of average body size (as plotted here).

Further stratifying postmenopausal females by history of HRT use (Methods) revealed distinct HGS-age, MM-age, and MQ-age trajectories for females with versus without a history of HRT (each prob_AW_ >99.9%; HGS/MM/MQ menopause model 4 in Supplementary Data 2; Fig. 4d–f), although it was not possible to determine whether these differences were due to the effects of HRT *per se*, or to systematic pre-existing differences between females that affected their HRT usage history. Importantly, differences between males and postmenopausal females were still apparent after stratifying the latter by HRT usage history (each prob_AW_ >99.9%; HGS/MM/MQ menopause models 5–6 in Supplementary Data 2), demonstrating that sexual dimorphism in muscle ageing persists irrespective of both female menopause status and history of HRT use.

Circulating levels and balance of the sex hormones testosterone and oestradiol did not reflect the patterns of male skeletal muscle ageing observed in this study, and only partially reflected those observed in females (Supplementary Fig. 10). In males between 40–70 yr, circulating levels of testosterone (an androgen) exhibited a decelerating decline, while circulating levels of oestradiol (an oestrogen) remained constant, meaning that neither hormone mirrored the accelerating declines in HGS, MM, and MQ observed over the same age range. In females, the relatively constant rates of age-related HGS and MQ decline observed during middle age were somewhat mirrored by a relatively steady decline in circulating testosterone levels, but not by circulating levels of oestradiol, which dropped sharply after the menopause, and presumably during perimenopause. Furthermore, this sharp drop in circulating oestradiol levels after menopause was at odds with the parallel MM- age trajectories of pre- and postmenopausal females. Finally, testosterone levels showed little difference between pre- and postmenopausal females, despite postmenopausal status being associated with significantly lower MM. Thus, circulating sex hormone levels do not evince explanations for muscle ageing trajectories in either sex.

## Discussion

Our analysis of 478,438 UKB participants aged 40–82 yr identified striking sexual dimorphism in arm muscle ageing. Whereas only a modest sex difference was detected in age-related hand grip strength (HGS) decline, age-related muscle mass (MM) decline was considerably more pronounced in males. Consequently, age-related decline of muscle quality (MQ)—an estimate of specific force—was substantially greater in females. These general trends held across cross-sectional and longitudinal analyses, persisted after accounting for systematic sex differences in body size, were apparent irrespective of female menopause status and HRT usage history, and did not reflect age-related changes in circulating sex hormone levels.

Our study provides strong evidence that age-related MM decline is substantially greater in males than females beyond 40 yr. Although the magnitude of this dimorphism is surprising, the general finding is in broad agreement with the available literature^30,36,52–54,57,59–66^. Perhaps more striking is the evidence that age-related MM decline is minimal in females after accounting for decline in overall body size, whereas male MM loss with age clearly exceeds that attributable to overall body size decrease. While this is a remarkable finding, it is nevertheless consistent with four low-powered studies, which also contain hints that age-related decline of female arm muscle size is minimal, including in the forearm^30,53,57,66^. We could not investigate the possibility that greater age-related MM decline in males beyond 40 yr simply represents a reversal of a comparatively larger gain between pubescence and 40 yr (e.g. due to higher levels of engagement in resistance exercise^35^ and manual labour^77^). However, the finding of Janssen et al. (2000) ^36^ that muscle mass remains largely stable in both sexes until ∼45 yr (N = 468) argues against this being the primary causal mechanism. Furthermore, it seems counterintuitive that greater male participation in physically demanding activities prior to 40 yr would result in an accelerated loss of MM—but not HGS— thereafter, as observed in this study, in which HGS-age trajectories of females and males were relatively parallel between 40–60 yr, whereas MM-age trajectories were not.

The pronounced sex difference in age-related MM decline is especially compelling in light of the comparatively modest sex difference in age-related HGS decline. Sexual dimorphism in age-related decline of HGS and MM ultimately presented as a greater age-related muscle quality (MQ and MQ’) decline in females across all ages and participants groups. This finding is compatible with Hakkinen et al. (1996) ^47^, who reported significantly lower quadriceps muscle quality in older (∼70 yr) than younger (∼50 yr) females, but no difference in analogous groups of males. Similarly, a longitudinal study of 511 participants >50 yr detected significantly faster age-related quadriceps muscle quality decline in females, although—unlike in our study—this difference was not statistically significant after adjusting for body size (standing height) ^68^. Muscle quality appears to be the sarcopenia-associated muscle phenotype for which age-sex differences are least characterised, and no consistent relationship is evident in the literature^46–48,50,65,68,69^. Nevertheless, a decline in function of remaining muscle mass is acknowledged as a key diagnostic criterion for sarcopenia in the most-recent European Working Group on Sarcopenia in Older People (EWGSOP2) guidelines^2^. Our finding of greater age-related muscle quality loss in females is plausible, given that the existing evidence supporting greater age-related muscle size decline in males is relatively strong^30,36,52–54,59–65^, whereas evidence of a correspondingly greater loss of male muscle strength is equivocal^19,35,47,48,50–52^.

The distinct muscle ageing trajectories of females and males may derive from intrinsic sex differences in the composition, architecture, and/or neural activation of human skeletal muscle. Although sex differences in these aspects of muscle physiology have been reported^16,20,31,65,78^, their alteration with age is less clear.

Females and males exhibit little difference in musculature until puberty, at which point a rapid and marked divergence occurs^31,35,79^. Most notably, the average muscle mass of adult males is considerably greater than that of adult females, even after accounting for systematic sex differences in body size^30–35^. In our study, male MM exhibited an accelerating decline between 40–70 yr, whereas the modest decline in female MM over this age range occurred at a relatively constant rate. Consequently, the magnitude of the sex difference in MM decreased with age. Because sex hormones—particularly male testosterone—are integral to muscle growth and maintenance^80–82^, convergence of the sex hormone milieus of females and males is an attractive *prima facie* explanation for the observed convergence of MM. However, our data do not support this explanation, or at least not a simple dose-response model between sex hormones and skeletal muscle mass. Firstly, the sex difference in average testosterone levels showed no convergence between 40–70 yr, instead remaining relatively constant. Secondly, circulating levels of testosterone exhibited a decelerating decline with age in males, whereas male MM decline clearly accelerated. Finally, testosterone levels were similar between pre- and postmenopausal females (as also reported elsewhere^83,84^), despite postmenopausal females having significantly lower MM than their premenopausal counterparts. Although the sex difference in circulating oestrogen (oestradiol) levels was markedly reduced between 40–70 yr, this was entirely due to lower circulating oestradiol levels in older females, as male circulating oestradiol levels remained constant with age. As our results are not consistent with oestradiol being a key driver of MM decline in either sex, the modest convergence of the sex hormone milieu of females and males that occurs during middle age does not appear to explain the coincident convergence of musculature.

In the search for a single physiological, cytological, or biochemical difference that is consistent with all aspects of the observed sexual dimorphism in muscle ageing trajectories, one attractive possibility is that females experience an age-associated replacement of force-generating contractile material with material that does not enhance isometric force. Age-related increase in intra- and intermuscular adipose tissue (IMAT) is well established^85,86^, but neither BIA nor DXA can detect IMAT^45,87,88^, meaning that greater IMAT accumulation with age in females could theoretically explain why age-related MM decline is substantially greater in males, whereas MQ decline is more pronounced in females. However, there is little evidence for more intramuscular adipocytes in females than males^89^, and a previous longitudinal study using CT scans found no significant relationship between thigh IMAT and muscle quality^68^. Moreover, MRI scans of the thighs of UKB participants also show that thigh IMAT volume correlates only weakly with fat-free thigh muscle volume^54^. Since both CT scans and MRI scans can reliably detect IMAT^45,90^, a sex difference in replacement of arm muscle tissue by IMAT with age seems unlikely to explain our findings, barring a radical difference in IMAT accretion between the upper and lower limbs. The potential for a sex difference in accumulation of alternative non-contractile material including 1) non-IMAT fat in the form of intramyofibre lipid droplets, 2) insoluble protein aggregates^91^, 3) non-contractile tissues (e.g. fibrous connective tissue), or 4) other cytoplasmic components (such as mitochondria or non-functional myosin) remains to be investigated.

Sexual dimorphism in muscle ageing also likely reflects age-sex differences in gene expression. At least two studies^66,92^ have reported that age-related changes in transcriptional profile are more pronounced in females. Interestingly, one of these studies^92^ was performed on upper arm (biceps brachii) muscle, and found that older females exhibited elevated transcription levels of genes associated with muscle inflammation and lipid infiltration. This finding led the authors to conclude that “*alteration of muscle “quality” during the aging process might be unfavorable to females*”, which supports our own finding of greater age-related muscle quality decline in females. Future genome-wide association studies (GWASs) on our muscle phenotypes will characterise the genetic architecture underpinning the age-sex differences we describe, and should also help to explain the high degree of variation observed in these phenotypes, much of which likely has a genetic basis^22,23,93,94^.

Other potential explanatory factors include age-sex differences in muscle architecture (Supplementary Discussion), or in health and lifestyle factors such as physical activity levels^25^, socio-economic circumstances^27^, diet^26^, and acquired disease^95^. Although systematic age-sex differences in health and lifestyle appear unlikely to explain fully the observed sexual dimorphism, it is possible that each factor plays a contributory role. Although the primary aim of our study was to characterise—rather than explain—sex differences in skeletal muscle ageing, it is nevertheless clear that future studies should examine how both intrinsic and extrinsic factors contribute to the sexual dimorphism that we report, as well as the potential for intrinsic factors to influence lifestyle in ‘virtuous’ or ‘vicious’ cycles.

An intriguing finding of our study was that postmenopausal females were expected to have ∼37 g less MM than their premenopausal counterparts after controlling for age and body size, despite there being no significant difference in shape of the MM-age trajectory between these two groups. This finding suggests substantial MM reduction during the brief (mean duration ∼1.3 yr^76^) perimenopausal phase, and is strikingly consistent with the findings of a recent literature review^83^, which concluded that 1) postmenopausal females have lower muscle mass than premenopausal females, and 2) oestradiol decline influences muscle mass loss during—but not after—menopause. Interestingly, our data offer no clear evidence of the protective effect of HRT on female muscle reported elsewhere^96,97^. However, we reiterate that it is not possible to determine whether the minor differences that we detected between postmenopausal females with and without a history of HRT use are directly attributable to HRT, or whether traits associated with muscle ageing are simply also associated with likelihood of undergoing HRT. The effects of perimenopause and HRT use on female muscle ageing warrant further investigation.

The wealth of phenotypic and genotypic data available in the UKB dataset has facilitated several large-scale studies on sarcopenia and relevant muscle phenotypes^24,34,54,64,93,98–104^. Nevertheless, the UKB cohort is not wholly representative of the general UK population, being healthier and less socio-economically deprived ^105^, and having an overrepresentation of white Europeans (94.6%^105^ vs 81.7% in England and Wales in 2021^106^). We found the health and socio-economic biases to be particularly pronounced in our Longitudinal Participants, who were typically even less deprived than the Cross-sectional Participants, and were demonstrably healthier, e.g. having lower mean body-mass index scores and fewer self-reported cancers than their cross-sectional counterparts (Methods). It is therefore apparent that the known volunteer bias in the UKB^105^ is compounded by a ‘retention bias’ in returning participants, a phenomenon that has been reported in other biobanks^107^. This bias towards healthy, affluent, white participants in the UKB means that caution is warranted when extrapolating the current findings to other populations—both in the UK and elsewhere—and highlights the need for more diverse biobank cohorts in which to study sex differences in muscle ageing.

HGS and MM measurement error represents another potential concern. HGS readings were taken on dynamometers, many of which showed evidence of miscalibration/measurement drift. We took steps to account for this—both by adjusting or excluding HGS readings, and by controlling for dynamometer-level effects within our models. Reassuringly, however, we detected no compelling evidence of a systematic effect of dynamometer measurement error, and therefore deem it unlikely that dynamometer effects introduced artefactual sexual dimorphism. Additionally, no evidence exists of a sex difference in voluntary activation during isometric contractions^35^, further indicating that our HGS readings reflect the true magnitude of sex difference in muscle strength. Most MM values in this study are not direct measurements, but BIA-derived estimates generated using novel predictive equations. We optimised these equations, and are confident that the MM estimates that they produce are superior to the default values provided in the UKB dataset (themselves estimates generated by predictive equations^108,109^). However, BIA estimates will inevitably be incorrect for some participants, and the possibility of systematic bias due to prediction error (e.g. persistent under- or overestimation of MM in one sex) cannot be discounted, although DXA measures showed close accordance with BIA estimates.

Our results are directly relevant to the detection and diagnosis of primary sarcopenia, a complex syndrome with no universally accepted diagnostic criteria^6,8,110^. Despite a 2018 literature review finding that ∼70% of research studies diagnosed sarcopenia using skeletal muscle mass alone^6^, the pronounced sex difference in age-related MM decline calls into question the suitability of muscle mass as an appropriate diagnostic criterion for sarcopenia. For example, muscle mass more than two standard deviations below the mean for a sex-specific reference population of healthy young adults is sometimes used to diagnose sarcopenia^3,55–58^, but sex differences in the age-mass relationship suggest that, for the median individual of each sex, these thresholds are likely to be passed at different ages^57^.

Furthermore, while sex-specific strength thresholds are often used in sarcopenia diagnosis^8^, older females and males in modern societies may not require substantially different levels of absolute strength to complete normal daily activities. Amara et al. (2003) ^30^ suggested that muscle-group-specific minimum strength thresholds might instead exist, below which day-to-day living may be hindered. Given the substantial sex difference in average strength, the decision to use muscle-group-specific or sex-specific strength thresholds must drastically impact the relative incidence of diagnosed sarcopenia in females versus males (Supplementary Discussion). Urgent consideration should therefore be given to whether sex-independent strength thresholds represent more useful diagnostic markers of sarcopenia than sex-specific ones. Our findings also highlight the necessity of focussing on sex in muscle ageing research, which frequently receives only superficial consideration, and is often disregarded entirely: for example, Haynes et al. (2020) ^111^ found that only ∼1% of studies of age-related muscle strength decline stratified results by sex. The dramatic sexual dimorphism in physiological and cell biological ageing-related changes of human muscle that we report demonstrate the importance of determining the mechanisms driving primary sarcopenia in each sex, and thus whether sex-specific approaches to mitigation of primary sarcopenia are required^65^.

## Methods

### Data preparation

Data (Supplementary Data 3) for 502,461 participants were retrieved from the UKB^70^. All analyses were conducted in R version 4.3.0^112^. Participant sex data (Data-Field 31) were acquired from NHS central registry at recruitment, but were in some cases updated if the participant reported that the central registry-obtained sex was incorrect^113^. 372 participants with a mismatch between recorded sex and genetic sex (Data-Field 22011) were excluded from analyses (Supplementary Data 1). No data on gender identity were used in this study. Our study and reporting adhere to Sex and Gender Equity in Research (SAGER) guidelines^114^. Age at assessment was calculated to the nearest month with the ‘lubridate’ R package^115^ using date of assessment and year and month of birth; as day of birth is not available in UKB, day of birth was nominally designated as the 15th for all participants. Season of attendance was defined as meteorological season (i.e. Spring: March–May; Summer: June–August; Autumn: September–November; Winter: December–February). Self-reported ethnicity was used to assign participants to one of three primary ethnic group categories used in the 2021 UK Census: Asian or Asian British (‘Asian’); Black, Black British, Caribbean or African (‘Black’); and White (‘White’) ^116^. For the purpose of analysis, all other participants (reporting Mixed or multiple ethnic groups, Other ethnic group, Do not know, Prefer not to answer, or NA) were grouped into a category termed ‘Other’. We chose not to exclude participants based on a history of medical conditions that might potentially affect muscle size or function, as preliminary analyses indicated that doing so would substantially reduce sample size without materially affecting overall trends (e.g. removal of >36,000 participants with one-or-more self-reported cancer diagnoses; Supplementary Fig. 11).

23,930 participants were excluded from this study for reasons including missing data and implausible anthropometric values. Participant exclusion flowcharts and the characteristics of the excluded participants are provided in Supplementary Data 1. The remaining 478,438 participants were split into those who attended an additional (‘repeat’) assessment in Cheadle, Newcastle, or Reading between April 2014 and March 2020 (‘Longitudinal Participants’, N = 45,629) and those who did not (‘Cross-sectional Participants’, N = 432,809) (Supplementary Data 1). A ‘retention bias’ towards healthier participants was evident in the Longitudinal Participants, who—compared to the Cross-sectional Participants—had significantly lower BMIs at baseline assessment (26.6 ± 0.02 kg/m^2^ vs 27.5 ± 0.007 kg/m^2^; *P* < 0.001; FPR < 0.001) and significantly fewer self-reported cancer diagnoses at baseline assessment (0.065 ± 0.001 vs 0.091 ± 0.0005; *P* < 0.001; FPR < 0.001). Longitudinal Participants were also significantly less socio-economically deprived than Cross-sectional Participants—as determined using the Townsend Deprivation Score [TDS] at baseline assessment (−1.89 ± 0.01 vs −1.27 vs 0.005 [higher scores indicate higher deprivation]; *P* < 0.001; FPR < 0.001)—and TDS correlates strongly with overall health^117^. These differences persisted after controlling for systematic age and sex differences between the two participant groups (*p*-values <0.001; FPRs <0.001—data not shown).

A ‘Menopause Cohort’ of 365,895 Cross-sectional Participants was also created, which included 170,101 females of known menopause status (premenopausal, N = 53,840; postmenopausal, N = 116,261) and all 195,794 males (Supplementary Data 1). Females who reported having undergone menopause outside of the typical ‘menopause window’ of 45–60 yr^76^ were removed from the participant group. Stratification of postmenopausal females by HRT usage history within this participant group was based on response to the question “Have you ever used hormone replacement therapy (HRT)?”: those who answered “Yes” (N = 49,699) or “No” (66,291) where included in HRT analyses (N = 115,990), whereas those who answered “Prefer not to answer” (N = 46) or “Do not know” (N = 225) were excluded.

### Data visualisation

All data were visualised using the ‘ggplot2’ R package^118^.

### Hand grip strength

Maximum hand grip strength readings in whole kilogram force units (kgf) were taken from seated participants using an adjustable Jamar J00105 hydraulic hand dynamometer (Lafayette Instrument Company, Lafayette, IN, USA), which measures force isometrically^119^. Hand grip strength readings were adjusted to account for observed dynamometer miscalibration and measurement drift, and a number of readings found to have been taken under extreme drift or miscalibration were excluded from the final analyses (Supplementary Data 4). Analyses were performed using the mean grip strength of both hands, as left and right hand showed similar trends (Supplementary Data 4).

### Muscle mass

The majority of muscle mass estimates presented in this study derive from BIA, a widely employed method of estimating body composition using electrical conductance between skin-surface electrodes^120^ that is considered sufficiently accurate for muscle mass measurements in large cohorts^3^. All BIA readings in the UKB dataset were taken with a Tanita Body Composition Analyser BC-418MA (Tanita Corporation, Tokyo, Japan), which uses an undisclosed, proprietary algorithm calibrated DXA scans^108^ to generate limb and whole-body estimates of muscle mass by combining observed impedance values with information on the age, sex, ethnicity, height, and body mass of the participant^109^ (also see Tanita Body Composition Analyser BC-418MA Instruction Manual).

DXA scan estimates of arm muscle mass were also available for a subset of 4,253 Longitudinal Participants (‘DXA-scanned participants’). All DXA scans were conducted at the Cheadle Assessment Centre on a GE-Lunar iDXA machine (GE Healthcare, Madison, Wisconsin, USA). DXA uses the differential attenuation of x-ray beams at two different energies to generate compartmental estimates of body composition^121^. Although DXA technically returns compartmental estimates of lean soft tissue (LST) rather than muscle mass, a large majority of LST is muscle, with only small amounts of skin and connective tissue present in LST^87^. It is therefore considered justified in practice to treat DXA LST readings as muscle mass estimates^87,122^. A 2018 study of 4,753 DXA-scanned UKB participants found excellent accordance between leg muscle mass estimates produced by DXA and MRI (R^2^ = 0.97), the latter of which is considered a ‘gold standard’ for regional body composition analysis^88^. DXA has also been shown to produce fat-free mass (i.e. LST, organs, and bone^87^) estimates that are more similar (i.e. show higher correlation and lower mean bias) to those produced by CT scans (another standard in regional body composition analysis^88^) than are those derived from BIA^123^.

We used the DXA-scanned participants to compare DXA mean arm muscle mass estimates with three sets of BIA-derived muscle mass estimates:

1. direct outputs from the Tanita Body Composition Analyser BC-418MA (‘Tanita’);
2. estimates generated using the equations of Powell et al. (2020) ^72^ (‘Powell’), which predict mean arm muscle mass using impedance values, age, sex, and anthropometric data readily available in the UKB (e.g. standing height and body mass; see Model C: Arms Lean Mass (g) in Supplementary Table 3 of Powell et al.);
3. estimates generated using our own novel equations (‘novel’), which were themselves based on the Powell equations (Supplementary Data 5).

The muscle mass estimates produced by our novel equations proved to be significantly more similar to the DXA-derived muscle mass estimates than did either the Powell or Tanita estimates (Supplementary Data 5). We therefore used our novel estimates of muscle mass when performing our primary analyses.

### Muscle quality

Muscle quality can be defined either as strength divided by muscle cross-sectional area (CSA) ^16^, or strength divided by muscle mass^7,45^. We therefore derived two measures of mean arm muscle quality: MQ, calculated as *(mean HGS)/(mean arm muscle CSA)*; and MQ’, calculated as *(mean HGS)/(mean arm MM*) (Supplementary Data 6).

Direct measurements of arm muscle CSA are not available for UKB participants, so CSA was instead modelled by dividing muscle mass by ulna length (UL). As an individual’s standing height can be predicted from their UL with high accuracy^124,125^, we reasoned that the reverse must also be true, and so rearranged equations originally developed to predict standing height from UL^124,125^, to instead estimate UL from standing height.

Standing height typically declines with age^38^, but an analogous decrease in UL is considered improbable^125^, suggesting that the standing height:UL ratio is unlikely to remain constant throughout life. If not controlled for, this phenomenon might result in an artefactual age-related decline in MQ (Supplementary Data 6). To account for this, we calculated UL using participants’ predicted maximum standing heights (PMSHs), rather than their observed standing heights. PMSHs of participants were predicted using previously published equations^38^ that are proven to accurately predict maximum standing height from observed standing height, sex, and age.

The six sex- and ethnicity-specific equations^125^ used to predict UL from PMSH were:

Asian Females: UL = (PMSH [cm] − 77.62) / 3.26

Asian Males: UL = (PMSH [cm] − 83.58) / 3.26

Black Females: UL = (PMSH [cm] − 79.55) / 3.14

Black Males: UL = (PMSH [cm] − 85.8) / 3.14

White Females: UL = (PMSH [cm] − 95.6) / 2.77

White Males: UL = (PMSH [cm] − 90.92) / 3.14

All equations except for the ‘White Females’ equation are re-arranged versions of equations originally published in Madden et al. (2020) ^125^. These equations were highly applicable to the UKB dataset, having been generated using a multi-ethnic British cohort comprising both sexes and spanning a wide age range (21–82 yr). The White Females equation was originally published in Elia (2003) ^124^, and is recommended for use for white females by Madden et al..

To avoid bias, participants in the ‘Other’ ethnic group category were randomly assigned one of the three sex-specific UL calculation equations at a ratio of 1:1:1.

A substantially greater age-related decline in female MQ persisted under various calculations of UL (Supplementary Data 6), indicating that our MQ measure was not compromised by the use of estimated or predicted values for UL, CSA, and maximum standing height.

### Participant matching

The *matchit* function of the ‘MatchIt’ R package^126^ was used to match female:male pairs based on their similarity in age, standing height, and body mass at baseline assessment. Nearest neighbour matching without replacement was performed, with Mahalanobis distances used as the measure of similarity. Males were arbitrarily designated as the ‘Treatment’ group—i.e. for each male a matching female was sought, rather than vice versa— due to there being fewer males than female Cross-sectional Participants and Longitudinal Participants (Table 1). Designating females as the Treatment group instead of males did not return meaningfully different results (data not shown). The *caliper* argument was used to prevent matching of participants whose values for a given criterion differed by more than a predefined amount (age at baseline assessment ≤ 0.5 yr; standing height at baseline assessment ≤ 1 cm; body mass at baseline assessment ≤ 1 kg). *matchit* returned 61,683 female:male pairs drawn from the Cross-sectional Participants (i.e. Matched Cross-sectional

Participants, N = 123,366, representing ∼29% of this participant group) and 5,435 female:male pairs drawn from the Longitudinal Participants (i.e. Matched Longitudinal Participants, N = 10,870; ∼24% of this participant group) that proved to be exceptionally well-matched based on recommended assessment criteria^126,127^ (Supplementary Data 7). Ethnicity was not included as a matching criterion, as inclusion further reduced the overall diversity of the matched participant groups (data not shown). Although participant matching is susceptible to an element of measurement error bias (e.g. potentially selecting some larger, under-measured males and smaller, over-measured females, thereby diminishing the precision of the matches), the effect is expected to be small given the relatively high accuracy of standing height and body mass measures^128^, their large male and female mean difference and variance, and the presumed larger measurement errors on HGS and BIA. Indeed, matching did little to diminish the sexual dimorphism observed in difference of HGS, MM, or MQ with age, either within the cross-sectional or longitudinal participant groups (Tables 2–4).

### Calculations and analyses

Comparison of means for two independent samples was performed using either Student’s *t*-test (when variances were equal) or Welch’s *t*-test (when variances were unequal); equality of variance was established beforehand using an *F*-test. False Positive Risk (FPR) scores were calculated using the methods of Sellke et al. (2001) ^129^, as discussed by Colquhoun (2019) ^130^, and implemented using the *pcal* function in the ‘pcal’ R package^131^. Skewness and kurtosis were tested using the ‘moments’ R package^132^, with skewness values <|2| and kurtosis values <|7| taken to indicate adequately normal distributions^133^ (also see Supplementary Data 6). Total percentage differences/changes were calculated as *((second measure − first measure)/first measure)·100*, and annual percentage differences/changes were calculated as *(second measure / first measure)^(^*^1^ *^/Interval)^ − 1*, where *Interval* is the recorded time (in years) between the two measures. α was set at 0.05 for significance tests.

### Regression analysis

#### Multiple linear regression

Multiple linear regression (MLR) models were fitted using the *lm* base R function, regressing a muscle trait (HGS, MM, or MQ) on age, sex, and an age-sex interaction term. 95% CIs for differences in slopes (β) were calculated as β ± 1.96·standard error (Supplementary Data 2).

#### Generalised additive mixed models

The relationship between each muscle trait (HGS, MM, or MQ) and age was modelled using generalised additive mixed models (GAMMs), with the muscle trait as the dependent variable. The models included smooth functions of age to allow complex ageing trajectories to be inferred (Supplementary Data 2). Sexual dimorphism in ageing trajectories was assessed by comparing pairs of GAMMs^134^: the first with a single (‘global’) age term that could not vary by sex, and the second with sex-specific splines. GAMMs were fitted using the *bam* function of the ‘mgcv’ R package^135^, with the fast restricted maximum likelihood (fREML) method utilised for smoothing parameter estimation.

Longitudinal models were fitted using data from two assessments per participant, which were explicitly linked within the models by including a random intercept for each participant. Random intercepts were used instead of random slopes, as the latter could not be estimated with only two datapoints per participant.

Sex, self-reported ethnicity, and season of attendance were included as fixed effect covariates in all models. UKB Assessment Centres and individual measurement devices were included in models as random effects unless otherwise specified, reflecting the fact that they represented realised instances of many possible Assessment Centres/devices. Standing height and body mass were also included in some models, to control for the potential contribution of body size to muscle trait differences (Supplementary Data 7).

To determine whether males and females exhibit distinct ageing trajectories of muscle traits, the relative quality of global and sex-specific models was assessed by calculating the difference between the Akaike Information Criterion (AIC) scores of the two models (‘ΔAIC’), with smaller AIC scores indicating better model quality (in the sense of minimising Kullback-Liebler information loss), and larger ΔAICs indicating larger differences in model quality^136,137^. ΔAICs were then converted to Akaike weights (AWs) for the two models using the *akaike.weights* function of the ‘qpcR’ R package^138^. Since each individual AW is always within the interval [0, 1] and AWs always sum to 1 for a given set of candidate models^136^, an AW is directly interpretable as the probability of the corresponding model being the ‘best’ approximating model within a candidate set^136,137,139^. AWs were therefore converted to percentage probabilities (denoted ‘prob_AW_’) for presentation purposes. For reference, 95%, 99%, and 99.9% probabilities of the lower-AIC model being the better approximating model in a two-model comparison correspond to ΔAICs of 5.89 (prob_AW_ = 95%), 9.19 (prob_AW_ = 99%), and 13.81 (prob_AW_ = 99.9%).

A similar approach was used to analyse the cross-sectional Menopause Participants dataset, but with ‘group’—i.e. ‘premenopausal females’, ‘postmenopausal females’, and ‘males’—replacing sex in the GAMMs. We fitted HGS, MM, and MQ models for each two-group combination in turn, and again interpreted low support for a global model (relative to a model with group-specific splines) as evidence of dimorphism in muscle ageing trajectories. This approach was deemed preferable to including all three groups within a single model, because inclusion of group-specific splines could potentially improve fit in such a model, even if trajectories were identical for two of the three groups.

The GAMM-fitted values presented in the Figures and Tables were estimated using the *predict.bam* function of the ‘mgcv’ R package, and unless otherwise specified include sex- or group-specific splines (e.g. HGS model 1_SEX-SPECIFIC_ [Supplementary Data 2] in Fig. 1a and Table 2), based on a White participant attending their UK Biobank assessment(s) during Spring. White and Spring were the most frequent factor levels for ‘ethnicity’ and ‘season of attendance’ within the Cross-sectional Participants, and so for consistency were used for all Figures; selecting other values for these variables would shift male and female curves on by an equal amount on the *y*-axis without changing the age trajectory. The King’s Computational Research, Engineering And Technology Environment (CREATE) ^140^ was used to generate some of the models included in this study. Full details of models and model comparisons are provided in Supplementary Data 2.

## Ethics statement

The UK Biobank study holds ethical approval from the North West Multi-centre Research Ethics Committee (REC reference 21/NW/0157) ^141^. All UK Biobank participants gave informed written consent at recruitment^142^. This study was performed under Research Project Number 67736 and complies with all relevant ethical regulations.

## Data Availability

All data analysed in this study are available from the UK Biobank^70^. The adjusted hand grip strength readings and novel estimates of muscle mass and muscle quality generated as part of this study will be returned to the UK Biobank for dissemination to other users.

## Code Availability

Identical copies of the complete code used in this study will be deposited with the UK Data Service (https://ukdataservice.ac.uk/) and the King’s Open Research Data System repository (https://kcl.figshare.com/) upon final acceptance of the manuscript.

## Acknowledgements

We thank Michael Simpson, Aleksandar Dimitrov, Neelanksha Garg, Alix Hughes, Marc Österdahl, and Richard Powell for their advice and input. SMH is a UKRI/Medical Research Council Scientist with MRC Programme Grant MR/N021231/1 support.

## Author Contributions

N.D. and S.M.H. conceived the study. N.D., C.R.O’N., T.W.F., and A.S. performed initial analyses. Final analyses were by N.D. and T.W.F.. Statistical and modelling approaches were developed by N.D., T.W.F., S.M.H., and H.E.O., and performed by T.W.F.. Manuscript writing was led by T.W.F., with input from all other authors. N.D. and S.M.H. jointly supervised the study.

## Competing Interests

The authors declare no competing interests.

## Supplementary Information

### Supplementary Figures

**Supplementary Figure 1.**
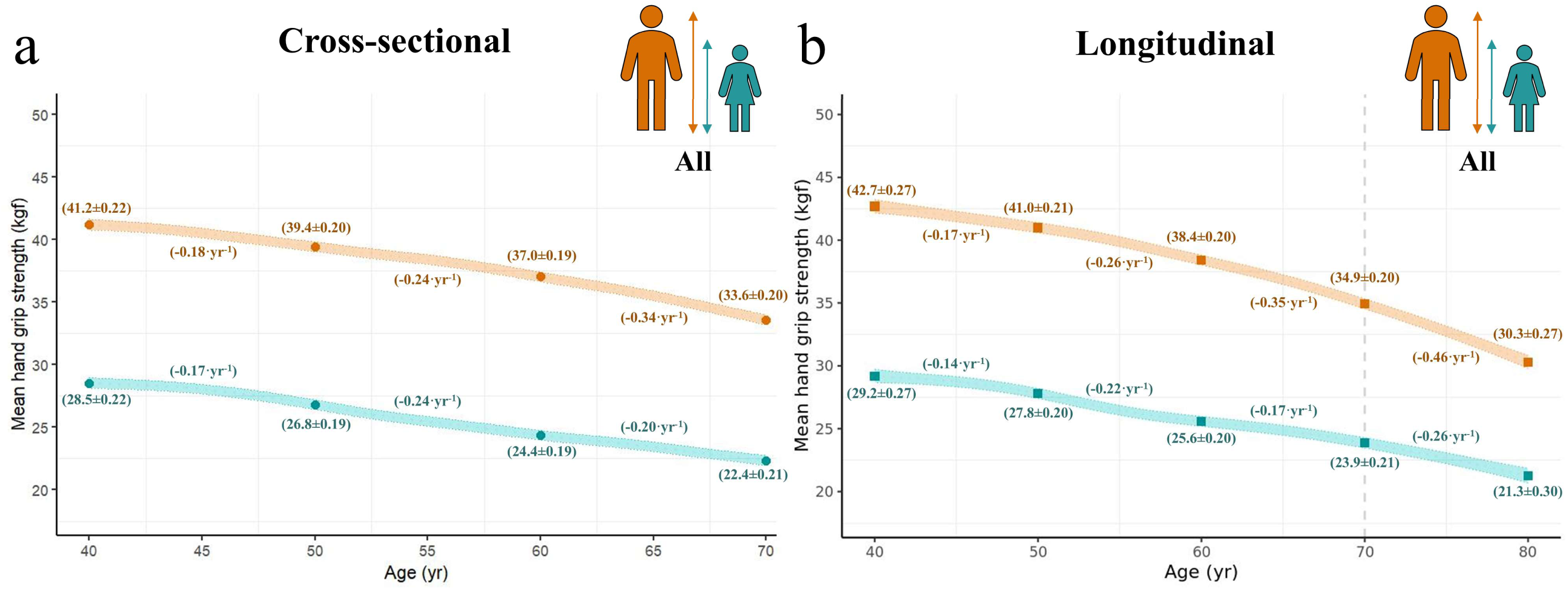
Modest sexual dimorphism in age-related strength decline persists after accounting for sex differences in body size. Generalised additive mixed model (GAMM)-fitted relationships between mean hand grip strength (HGS) and age are shown for **a** Cross-sectional Participants and **b** Longitudinal Participants (GAMM accounts for repeated measures, and therefore estimates true longitudinal change in HGS). Coloured bands (females, teal; males, orange) represent the 95% confidence intervals for the GAMM-fitted relationships (plotted with covariates fixed at ‘White’ for ethnicity and ‘Spring’ for season of assessment—Methods). Large points and values in parentheses are fitted values (± SEM) at 40, 50, 60, 69 (Cross-sectional Participants only), 70 (Longitudinal Participants only), and 80 (Longitudinal Participants only) yr, and inferred annual change over the intervening period. The models include standing height and body mass as covariates, to control for the potential contribution to HGS of body size differences. Standing height and body mass are fixed for plotting purposes at 168.97 cm and 78.85 kg, respectively, values equal to *(mean female value + mean male value)/2* in the Cross-sectional Participants. Complete model specifications are provided in Supplementary Data 2 under headings ‘HGS Model 3’ (a) and ‘HGS Model 4’ (b). The general trends in age-related HGS decline presented in the main manuscript therefore persist after accounting for body size differences by age and sex.

**Supplementary Figure 2.**
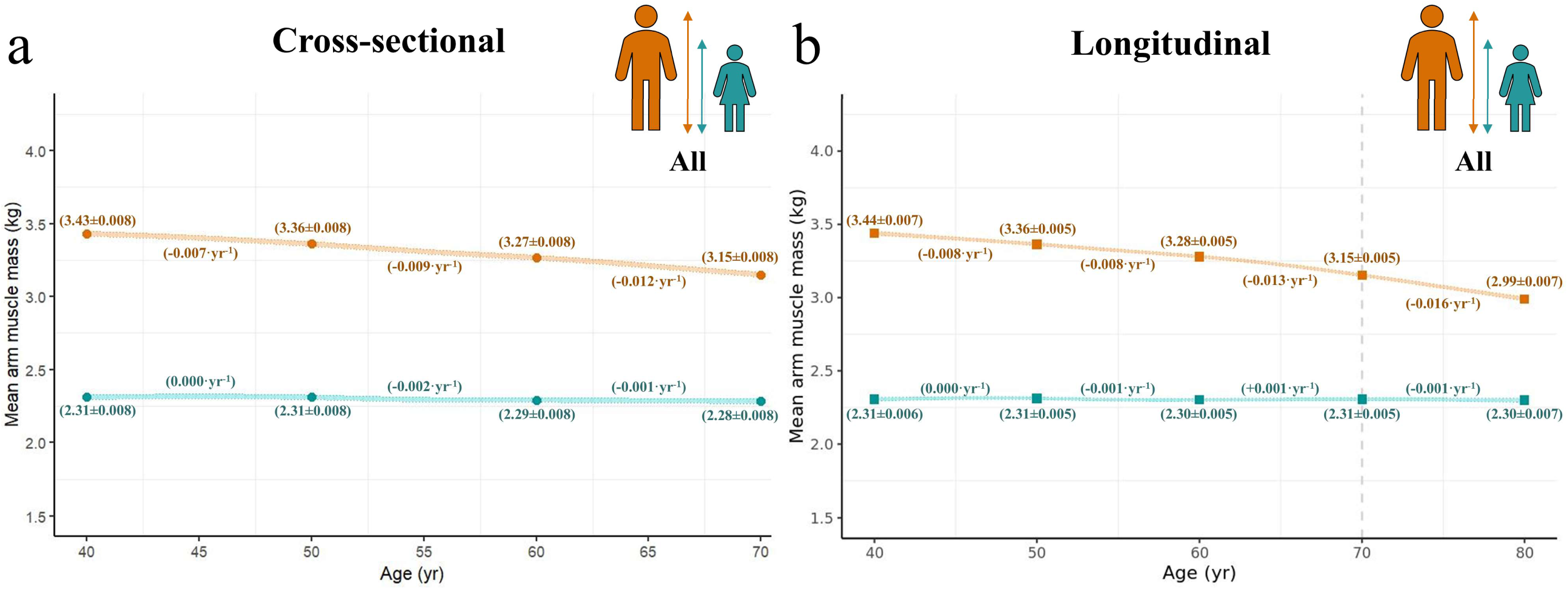
Age-related muscle mass decline remains substantially greater in males than females after accounting for sex differences in body size. Generalised additive mixed model (GAMM)-fitted relationships between mean arm muscle mass (MM) and age are shown for **a** Cross-sectional Participants and **b** Longitudinal Participants (GAMM accounts for repeated measures, and therefore estimates true longitudinal change in MM). Coloured bands (females, teal; males, orange) represent the 95% confidence intervals for the GAMM-fitted relationships (plotted with covariates fixed at ‘White’ for ethnicity and ‘Spring’ for season of assessment—Methods). Large points and values in parentheses are fitted values (± SEM) at 40, 50, 60, 69 (Cross-sectional Participants only), 70 (Longitudinal Participants only), and 80 (Longitudinal Participants only) yr, and inferred annual change over the intervening period. The models include standing height and body mass as covariates, to control for the potential contribution to MM of body size differences. Standing height and body mass are fixed for plotting purposes at 168.97 cm and 78.85 kg, respectively, values equal to *(mean female value + mean male value)/2* in the Cross-sectional Participants. Complete model specifications are provided in Supplementary Data 2 under headings ‘MM Model 3’ (a) and ‘MM Model 4’ (b). Intriguingly, female MM remains almost constant with age, in contrast to the modest age-related MM decline in females over ∼50 yr identified in our primary analyses (Fig. 2). Whereas participant matching allows for a relationship between age and body size (e.g. approx. −0.15 cm⋅yr^−1^ standing height and −0.07 kg⋅yr^−1^ body mass in female Matched Cross-sectional Participants), the modelling approach utilised here does not; the discordant findings of the two approaches suggest that age-related female MM decline reflects the inverse correlation between age and body size.

**Supplementary Figure 3.**
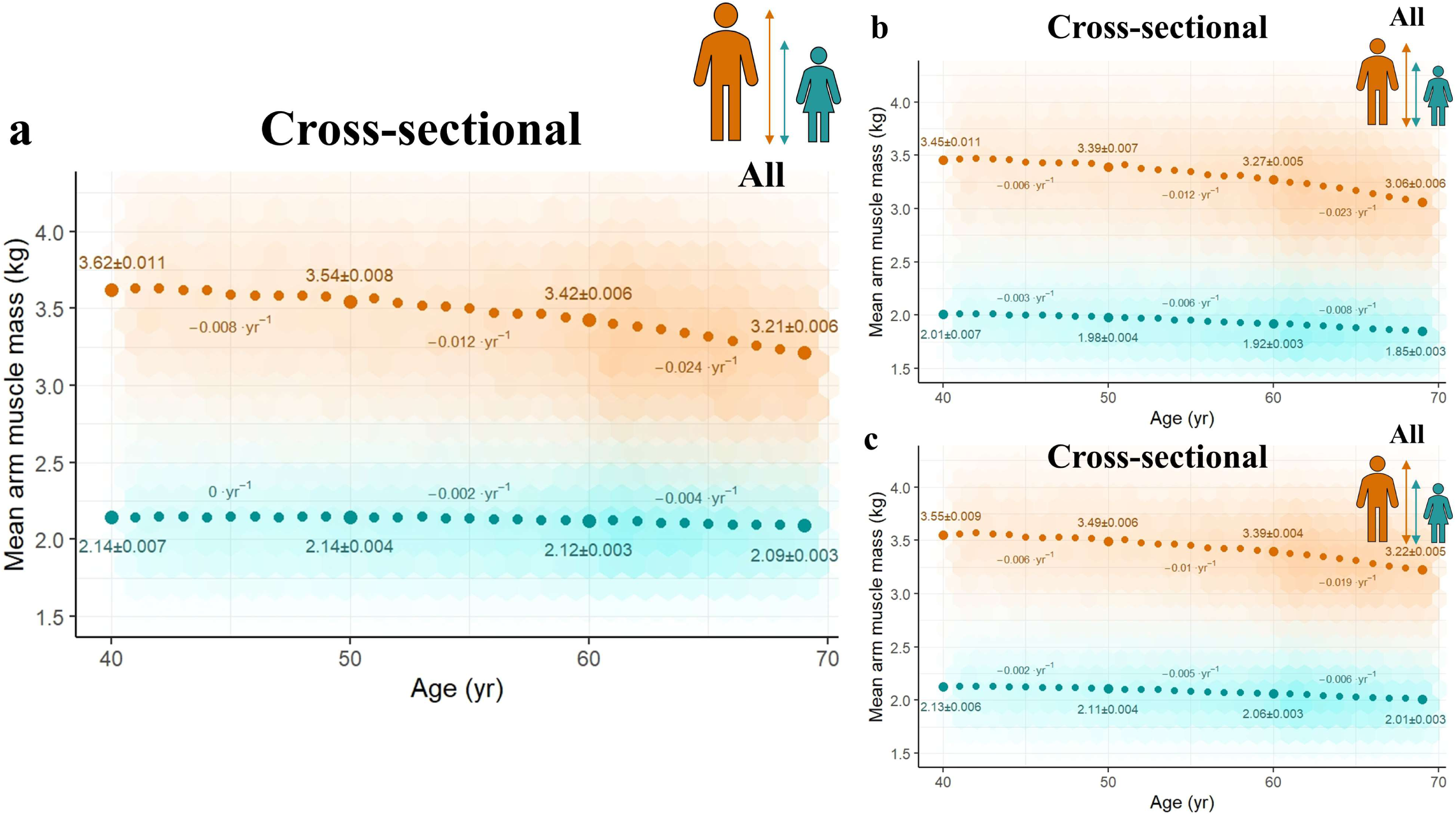
Patterns of age-related muscle mass decline are similar across three alternative sets of muscle mass estimates derived from bioelectrical impedance analysis. Sex-specific relationships between mean arm muscle mass and age within the Cross-sectional Participants (females, teal; males, orange). The observed data are plotted using hexagonal bin plotting, with greater opacity indicating higher density of participant measures. Large points and values are observed mean values (± SEM) at 40, 50, 60, and 69 yr, and inferred annual change over the intervening period. *y*-axes are truncated to better visualise the general trends in the data. **a** Direct outputs from a Tanita Body Composition Analyser BC-418MA. **b** Estimates generated using the sex-specific equations of Powell et al. (2020) (referenced in main manuscript). **c** Estimates generated using our novel sex-specific equations, derived using unadjusted impedance (Ω) values (Supplementary Data 5). Although the magnitude of the sexual dimorphism differs slightly between the datasets, the primary trends of accelerating decline and substantially greater age-related muscle mass decline in males hold across all three sets of muscle mass estimates.

**Supplementary Figure 4.**
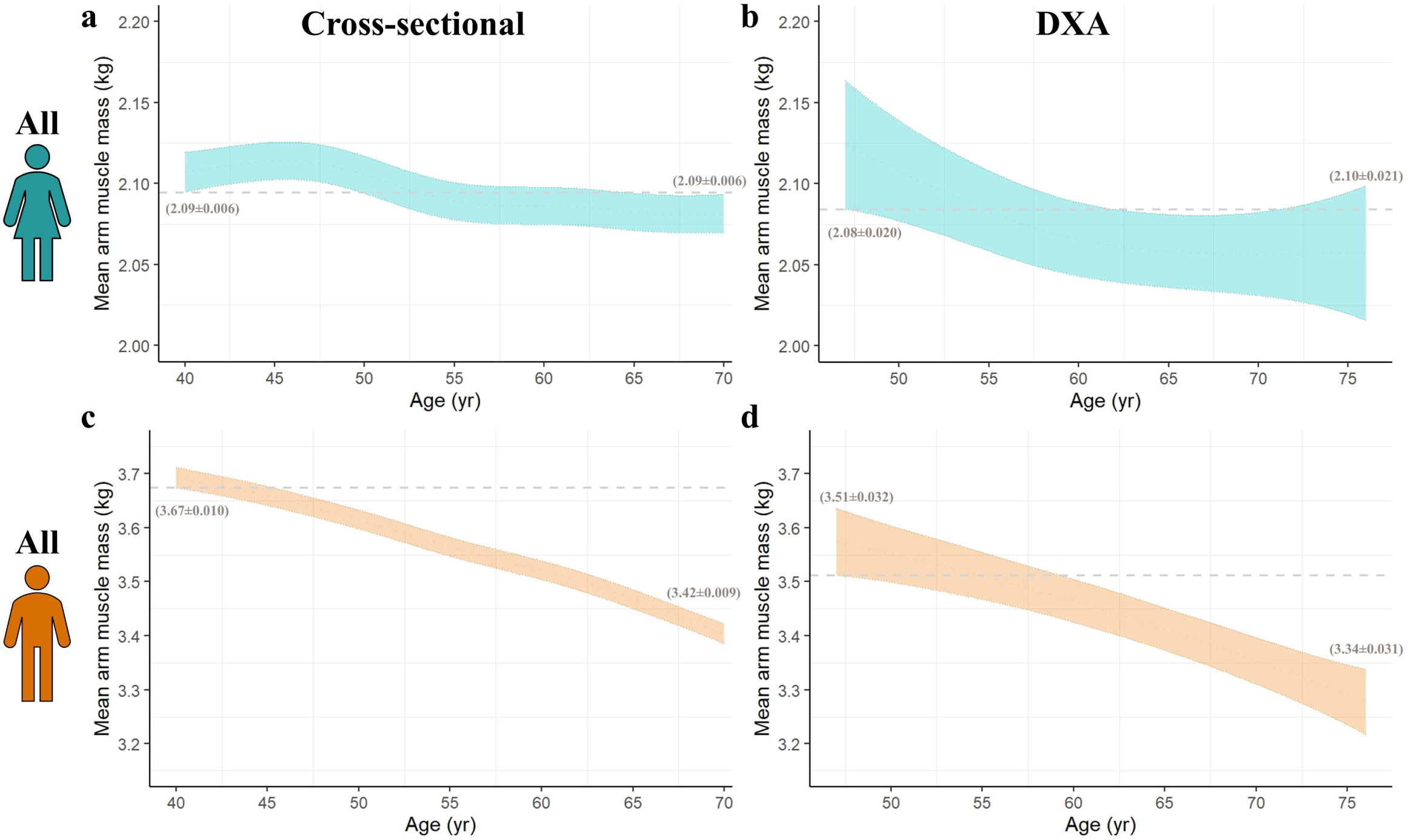
Female arm muscle mass exhibits minimal decline with age after accounting for age-related body-size decline. Generalised additive mixed model (GAMM)-fitted relationships between mean arm muscle mass and age. **a,c** bioelectrical impedance analysis (BIA)-derived measures from Cross-sectional Participants. **b,d** Dual x-ray absorptiometry (DXA)-derived measures from a subset of 4,253 DXA-scanned Longitudinal Participants ≥44.5 and ≤78 yr, analysed cross-sectionally. Coloured bands (females, teal; males, orange) represent the 95% confidence intervals (CIs) for the GAMM-fitted relationships (plotted with covariates fixed at ‘White’ for ethnicity and ‘Spring’ for season of assessment—Methods). Values (mean ± SEM) are the lower bound of the CI at 40 yr (BIA) or 47 yr (DXA), and the upper bound of the CI at 70 yr (BIA) or 76 yr (DXA). The models include standing height and body mass as covariates, to control for the potential contribution to muscle mass of body size differences. Standing height and body mass are fixed for plotting purposes at sex-specific means for Cross-sectional Participants (females: 162.38 cm and 71.63 kg; males: 175.55 cm and 86.07 kg). In both female datasets the lower bound of the CI at the lowest age (dashed grey line) overlaps the upper bound at the greatest age, indicating minimal difference in fitted mean arm muscle mass values at 40 vs 70 yr (a) or 47 vs 76 yr (b) in the absence of age-related body-size decline. In contrast, substantial age-related male mean arm muscle mass decline is indicated in both datasets (c,d). Complete model specifications are provided in Supplementary Data 2 under heading ‘Supplementary Fig. 4’.

**Supplementary Figure 5.**
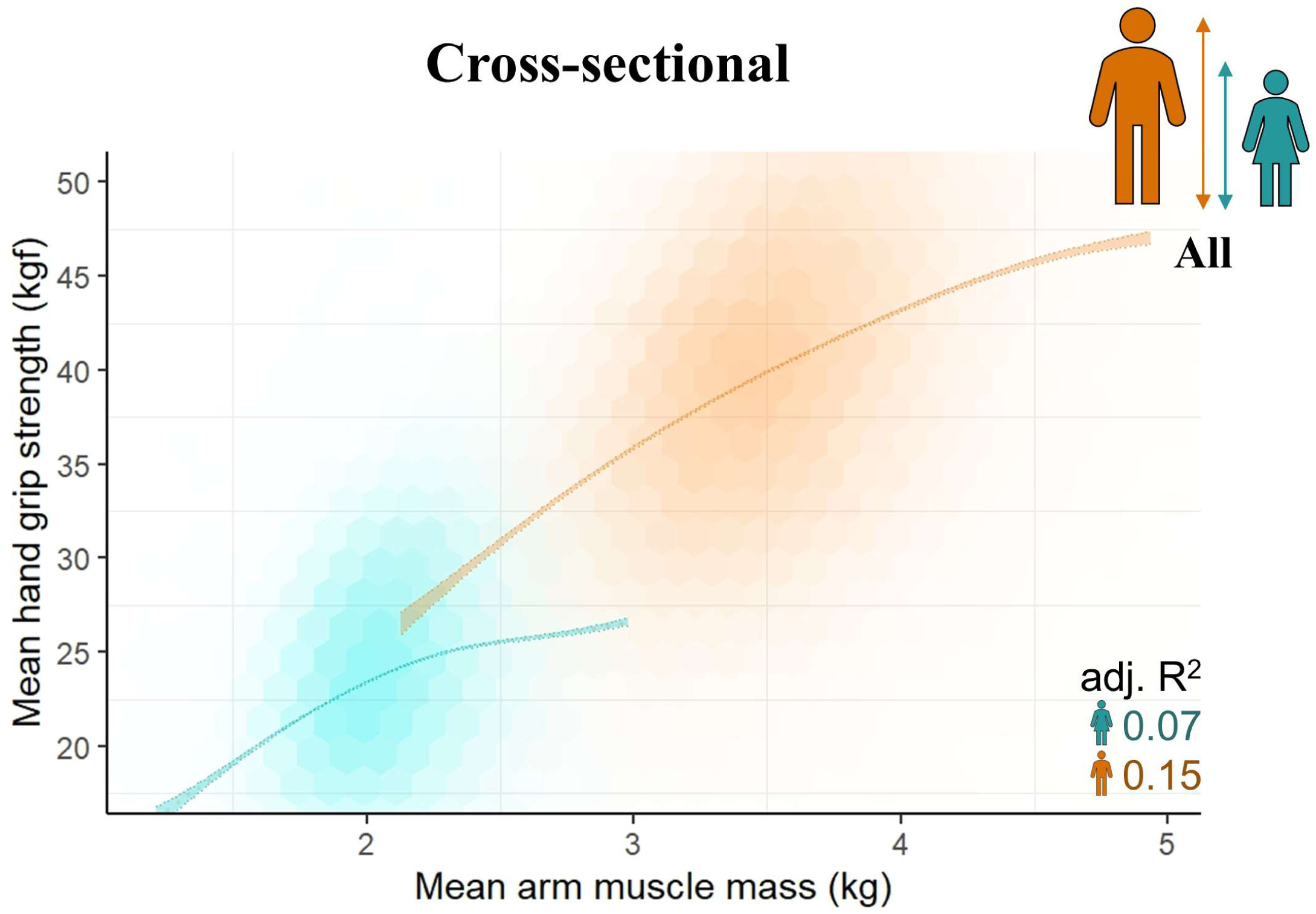
Variance in arm muscle mass does not explain most of the variance in hand grip strength, and the strength-mass correlation is stonger in males. The relationship between mean hand grip strength (HGS) and mean arm muscle mass (MM) within the Cross-sectional Participants (females, teal; males, orange). The observed data are plotted using hexagonal bin plotting, with greater opacity indicating higher density of participant measures. Coloured bands represent the 95% confidence intervals for the sex-specific generalised additive mixed model (GAMM)-fitted relationships *HGS ∼ s(MM),* where *s(MM)* denotes a spline fitted to MM (see Methods and Supplementary Data 2 for more details on GAMM-fitting procedures). Adjusted R^2^ values indicate that most of the variance in HGS is not explained by variance in MM, and that the correlation between HGS and MM is around twice as strong in males (adj. R^2^ = 0.15) as in females (adj. R^2^ = 0.07). These preliminary findings prompted us to investigate further the concept of muscle ‘quality’, including the potential for systematic age-sex muscle quality differences. See Supplementary Data 6 for more rigorous analysis of our muscle quality measures (note that this figure is also presented in Supplementary Data 6 as Figure SD6.Ib).

**Supplementary Figure 6.**
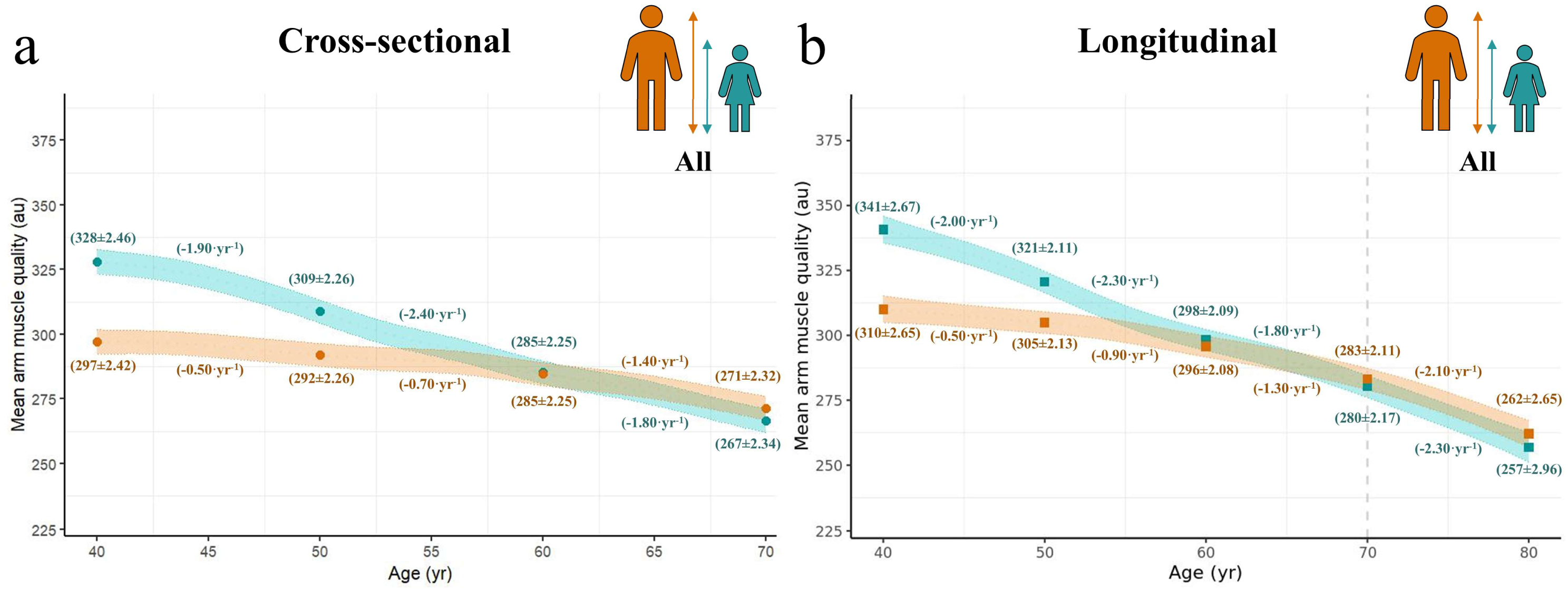
Age-related muscle quality decline remains substantially greater in females than males after accounting for sex differences in body size. Generalised additive mixed model (GAMM)-fitted relationships between mean arm muscle quality (MQ) (measured in arbitrary units [au]) and age are shown for **a** Cross-sectional Participants and **b** Longitudinal Participants (GAMM accounts for repeated measures, and therefore estimates true longitudinal change in MQ). Coloured bands (females, teal; males, orange) represent the 95% confidence intervals for the GAMM-fitted relationships (plotted with covariates fixed at ‘White’ for ethnicity and ‘Spring’ for season of assessment—Methods). Large points and values in parentheses are fitted values (± SEM) at 40, 50, 60, 69 (Cross-sectional Participants only), 70 (Longitudinal Participants only), and 80 (Longitudinal Participants only) yr, and inferred annual change over the intervening period. The models include standing height and body mass as covariates, to control for the potential contribution to MQ of body size differences. Standing height and body mass are fixed for plotting at 168.97 cm and 78.85 kg, respectively, values equal to *(mean female value + mean male value)/2* in the Cross-sectional Participants. Complete model specifications are provided in Supplementary Data 2 under headings ‘MQ Model 3’ (a) and ‘MQ Model 4’ (b).

**Supplementary Figure 7.**
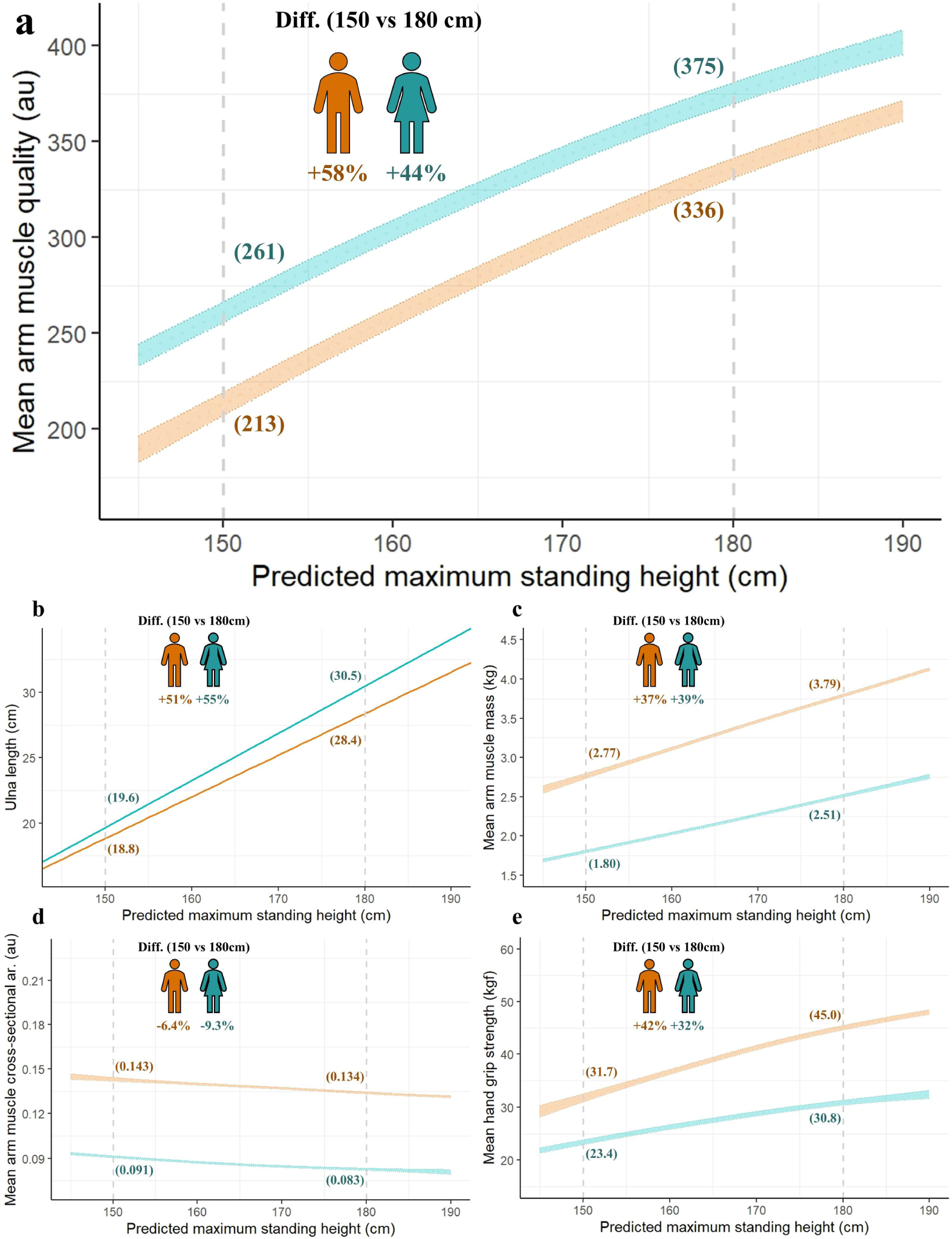
Muscle quality is influenced by complex and sex-specific allometric relationships between standing height, ulna length, arm muscle mass, and arm muscle cross-sectional area. Relationships are shown for Cross-sectional Participants (females, teal; males, orange). Values in parentheses are fitted values at a predicted maximum standing height (PMSH—Methods) of 150 cm or 180 cm (dashed vertical lines). Coloured bands in panels a and c–e represent 95% confidence intervals for generalised additive mixed model (GAMM)-fitted relationships (plotted with covariates fixed at ‘White’ for ethnicity, ‘Spring’ for season of assessment, and 40 yr for age—Methods). Coloured lines in panel b represent the fitted relationships between ulna length (UL) and PMSH for white females (UL = (PMSH [cm] − 95.6) / 2.77) and white males (UL = (PMSH [cm] − 90.92) / 3.14) (Methods). **a** Mean arm muscle quality (MQ) is positively correlated with PMSH, such that MQ at a PMSH of 180 cm is 44% greater (females) or 58% greater (males) than at 150 cm. **b–d** Ulna length (b) scales more strongly with PMSH than does mean arm muscle mass (MM) (c), leading to an inverse relationship between mean arm muscle cross-sectional area (CSA)—defined as *MM/UL*—and PMSH (d), and implying non-isometric scaling of arm muscle. **e** Mean hand grip strength (HGS) correlates positively with PMSH; as *MQ = HGS/CSA*, the positive correlation between MQ and PMSH arises because greater PMSHs are generally associated with both higher HGS (numerator) and lower CSA (denominator). Also see Supplementary Discussion. Complete model specifications are provided in Supplementary Data 2 under heading ‘Supplementary Fig. 7’.

**Supplementary Figure 8.**
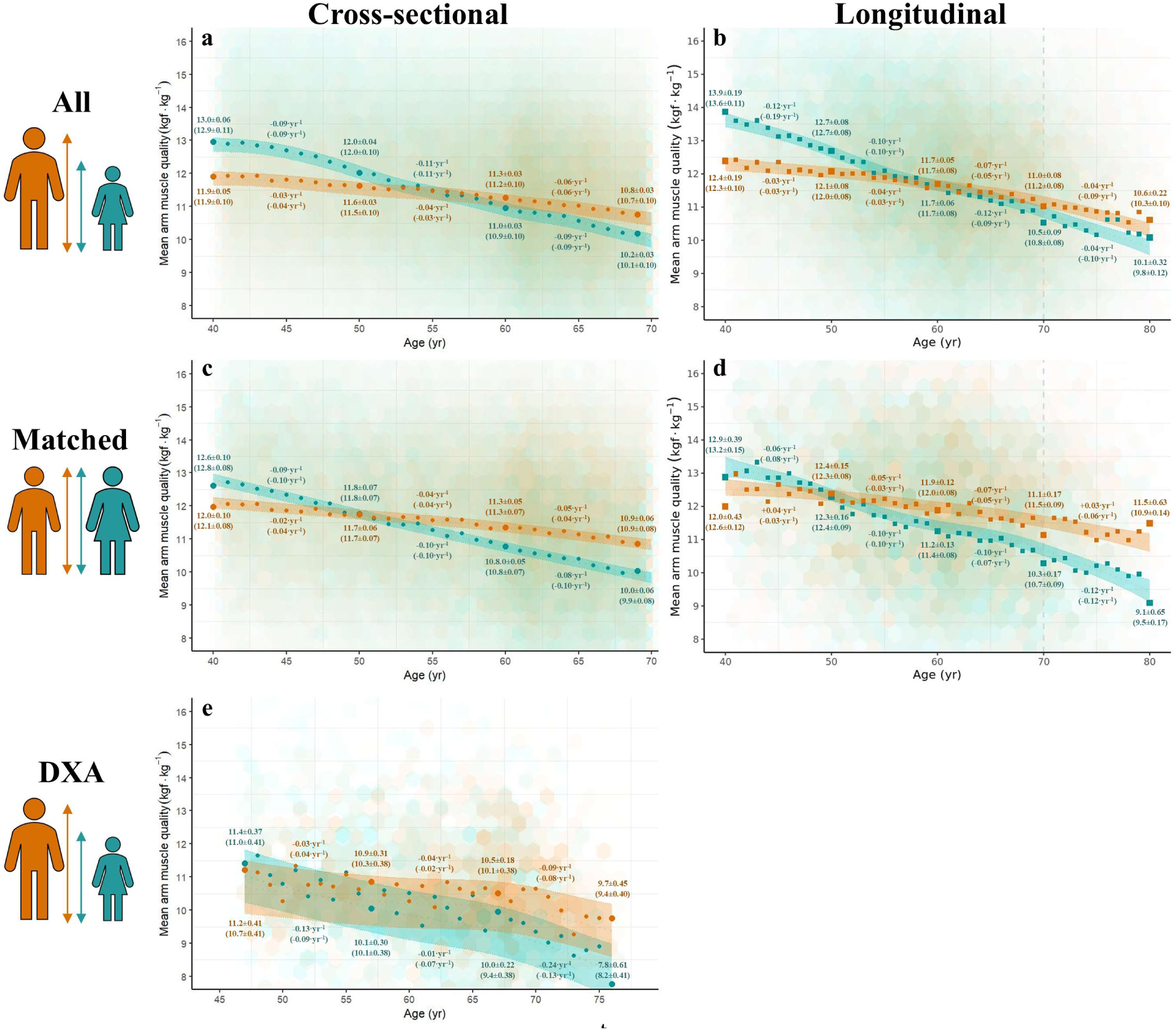
A second measure of muscle quality (MQ’) corroborates greater age-related muscle quality decline in females than males. Sex-specific relationships between between mean arm muscle quality (MQ’)—calculated as mean hand grip strength (HGS) divided by mean arm muscle mass (MM)—and age for UK Biobank participants (females, teal; males, orange). **a** Cross-sectional participants (N = 368,336). **b** Longitudinal participants (N = 32,663). **c** Matched Cross-sectional Participants (N = 104,820). **d** Matched Longitudinal Participants (N = 7,778). **e** Dual X-ray Absorptiometry (DXA)-derived MQ’ estimates for a subset of Longitudinal Participants ≥44.5 and ≤78 yr (one observation per participant) (N = 3,972). Hexagonal bins: observed data, with greater opacity indicating higher density of participant measures. Coloured bands show 95% confidence interval for the MQ’-age relationship fitted by a generalised additive mixed model (GAMM) (plotted with covariates fixed at ‘White’ for ethnicity and ‘Spring’ for season of assessment—Methods). Complete specifications for all models are provided in Supplementary Data 2, with fitted relationships in a, c, and e based on models MQ’ models 1, 3, and 5 respectively. Fitted relationships in b and d are based on MQ’ models 2 and 4, respectively, that account for repeated measures (and therefore estimate true longitudinal change in MQ’). Points represent observed mean values for each year age-group. Large points and values without parentheses are observed mean values (± SEM) at 40, 50, 60, and 69 yr (a,c), 40, 50, 60, 70 and 80 yr (b,d), or 47, 57, 67, and 76 yr (e), and inferred annual change (kgf·kg^−1^·yr^−1^) over the intervening period. The equivalent fitted values are given within parentheses. Sexual dimorphism is strongly supported for all models (each prob_AW_ >99.9%).

**Supplementary Figure 9.**
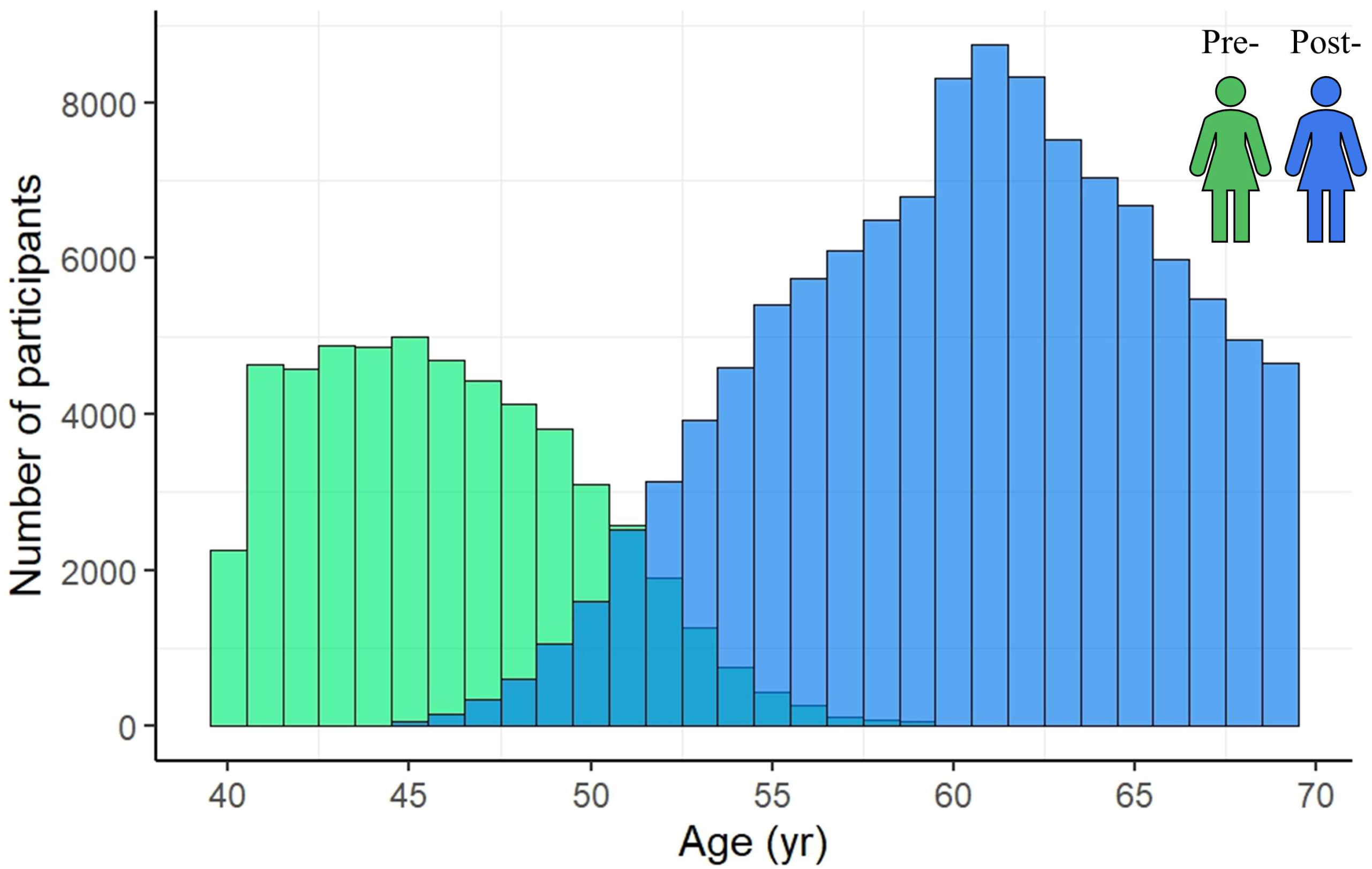
Age distribution of females in the Menopause Participants group, stratified by menopause status. Green and blue represent premenopausal (N = 53,840) and postmenopausal (N = 116,261) females, respectively.

**Supplementary Figure 10.**
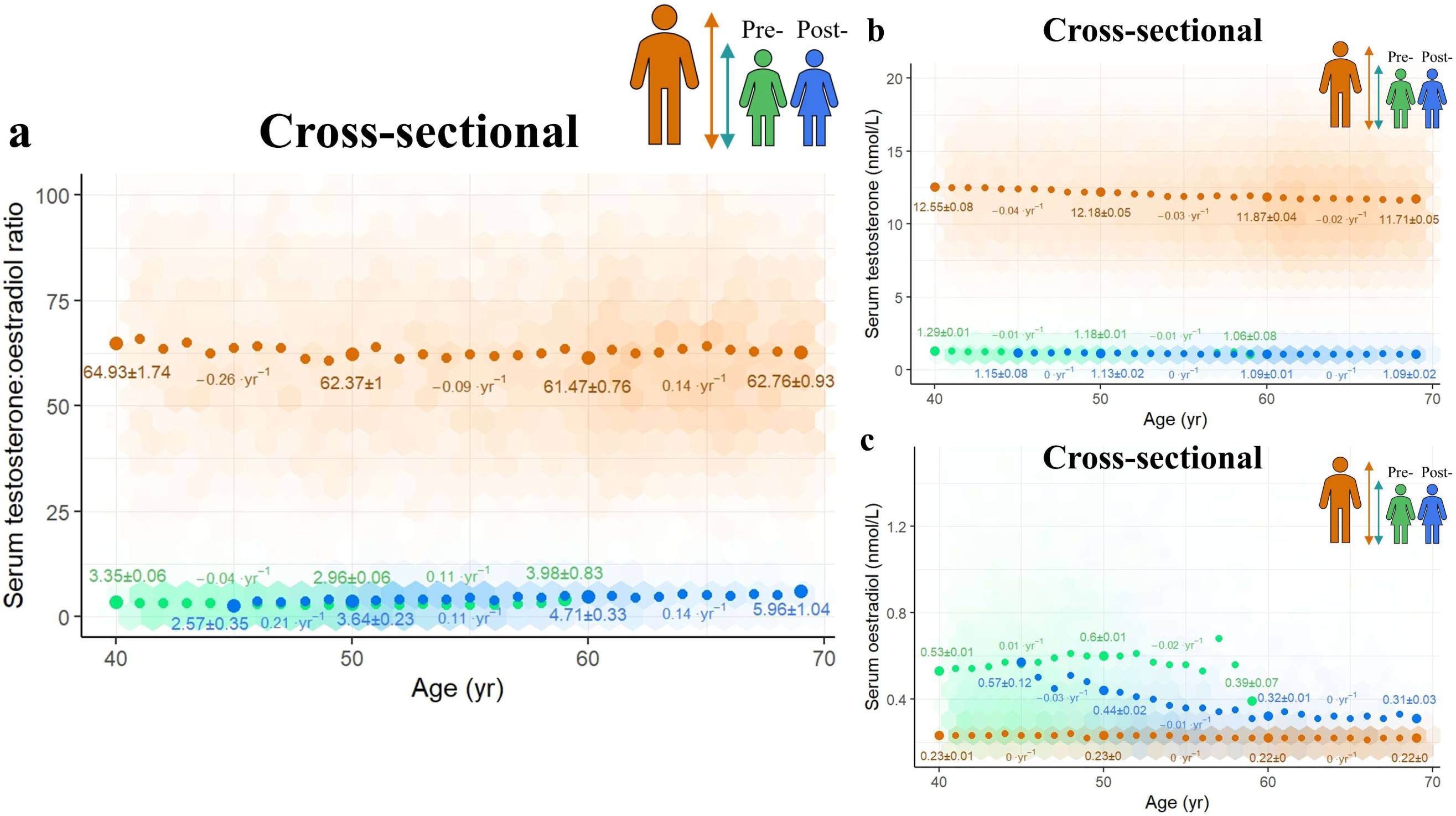
Modest age-related convergence of the sex hormone milieus of females and males is driven by declining female oestradiol levels. Group-specific relationships between sex hormone levels and age within the Menopause Participants, a participant group comprising all male Cross-sectional Participants and a subset of female Cross-sectional Participants (Supplementary Data 1). Females are stratified by menopause status, with green and blue representing pre- and postmenopausal females, respectively; orange represents males. The observed data are plotted using hexagonal bin plotting, with greater opacity indicating higher density of participant measures. Large points and values are observed mean values (± SEM) at 40, 45 (postmenopausal females only), 50, 60, and 69 (males and postmenopausal females only) yr, and inferred annual change over the intervening period. *y*-axes are truncated to better visualise the general trends in the data. **a** The serum testosterone:oestradiol ratio exhibits modest convergence between 40–69 yr. This convergence is apparently driven by an age-related decline in female serum oestradiol levels, because 1) sex differences in serum testosterone levels arguably become more pronounced with age (e.g. mean testosterone levels are 9.7-fold greater in males than females at 40 yr, rising to 10.7-fold greater at 69 yr) (**b**), and 2) male serum oestradiol levels exhibit no relationship with age (**c**).

**Supplementary Figure 11.**
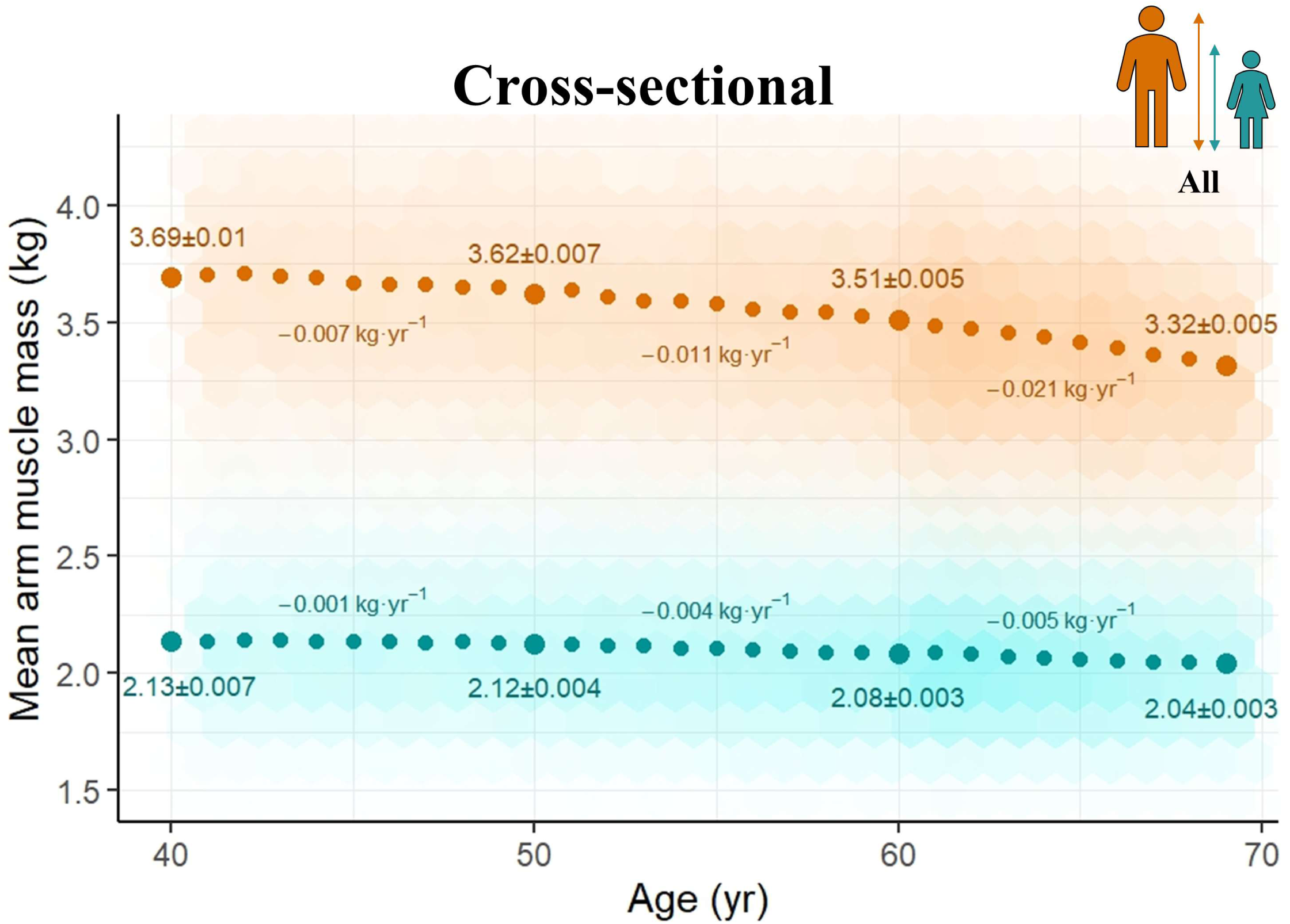
Excluding participants with self-reported history of cancer has little effect on population-level arm muscle mass trends. Scatterplot of the relationship between mean arm muscle mass (MM) and age in 396,177 Cross-sectional Participants with no self-reported history of cancer (i.e. those with a value of ‘0’ for ‘Number of self-reported cancers’— Data-Field 134). Teal and orange represent females (N = 213,212) and males (N = 182,965), respectively. Large points and values are observed mean values (± SEM) at 40, 50, 60, and 69 yr, and inferred annual change over the intervening period. Observations are plotted using hexagonal bin plotting, with greater opacity indicating higher density of participant measures. *y*-axes are truncated to visualise better the general trends in the data. Note that the results are essentially identical to those of the Cross-sectional Participants as a whole (main manuscript, Fig. 2a).

## Supplementary Discussion

### Muscle architecture

Muscle architecture is a potential mediator of age-sex differences in MQ. The positive correlation between standing height and MQ apparent in both sexes in our study reflects the fact that greater standing heights are typically associated with both higher HGS and lower CSA, i.e. the numerator and denominator of our MQ measure, respectively. Both MM and UL scaled positively allometrically with standing height in our dataset; UL scaled more strongly with standing height than with MM, meaning that taller individuals were expected to have more elongate arm muscles—and thus smaller CSAs—despite having greater MM (Supplementary Fig. 7).

The positive allometric scaling between UL and standing height in our data ultimately derives from the predictive equations that we used to estimate the former from the latter (Methods). However, positive allometric scaling between arm length and standing height—including forearm (radius) length—has been reported elsewhere^1,2^. An inverse correlation between muscle quality and CSA (Supplementary Data 6, Figure SD6.Ie) has also been observed in at least one study^3^. Both the increase in MQ with standing height and the inverse correlation between MQ and CSA are consistent with a pennate muscle architecture model under which elongated forearm muscles in taller individuals contain more muscle fibres, thereby increasing physiological cross-sectional area (PCSA)—i.e. the summed component of force deriving from the cross-sectional areas of all fibres within a muscle—at a given CSA^4^. There is evidence that muscle fibre pennation angles are typically greater in males^5–10^: while greater pennation angles reduce the specific force of individual muscle fibres, this loss can be more-than offset by the denser muscle fibre packing that is possible at greater pennation angles^11^, suggesting that denser muscle fibre packing in males may lead to a relative increase in male PCSA, and thus greater MQ at a given CSA. Interestingly, vastus lateralis muscle fibre pennation angles have been shown to decrease with age in both females and males^5^. If this reduction in pennation angle occurs to compensate for age-related loss of muscle fibres^12,13^, then it would be expected to lead to age-related MQ decline in both sexes, as observed in our study.

We were concerned that two aspects of using muscle CSA to determine MQ could introduce artefactual sexual dimorphism. Firstly, due to the oblique pennation angle of their fibres, some muscles generate force greater than that predicted by their CSA, and this angle could potentially show changing sexual dimorphism with age. Secondly, CSA is calculated from standing height, but males and females may lose standing height differently with age, thereby representing another potential source of artefactual sexual dimorphism. However, our MQ’ (*HGS/MM*) measures were not calculated using either standing height or CSA, yet also supported a greater age-related muscle quality decline in females than males, including when DXA-derived muscle mass measures (which are not calculated using any body size terms^14^) we used to derive muscle quality. We are therefore confident that our key finding of greater age-related muscle quality decline in females reflects a biological truth, although the degree to which age-sex differences in muscle architecture contribute to this sexual dimorphism remains an open question warranting further investigation.

### Sarcopenia thresholds

Amara et al. (2003)^15^ suggested that muscle-group-specific minimum strength thresholds might exist, below which day-to-day living may be hindered. These they estimated at ∼150 N (∼15.3 kgf) for hand grip strength and ∼400 N (∼40.8 kgf) for the plantar flexor muscles. Interestingly, the hand grip strength threshold of ∼15.3 kgf is similar to the 16 kgf threshold for females given in the most-recent European Working Group on Sarcopenia in Older People (EWGSOP2) guidelines, but well below the 27 kgf threshold for males^16^. In fact, of 12,823 Longitudinal Participants aged ≥70 yr at repeat assessment in our study, 662 of the 688 participants with a HGS reading <150 N are female (96.2%). As females comprise only 44.5% of Longitudinal Participants ≥70 yr at repeat assessment (N = 5,700), females ≥70 yr are almost 32 times as likely as males ≥70 yr to fall below the 150 N / 15.3 kgf threshold in this simple analysis (11.6% of females vs 0.4% of males). Additionally, HGS readings of 11.2% of males ≥70 yr (799/7,123) are ≥150 N but <27 kgf, meaning that these participants meet the sarcopenia criterion of EWGSOP2, yet remain above the functional threshold suggested by Amara et al.. The decision of whether a sex-specific or muscle-group-specific strength threshold is used should ultimately depend on the muscle under assessment, and its particular function; for instance, the substantially greater average body weight of males suggests that sex-specific thresholds might be appropriate for muscles used for supporting body weight. More broadly, our findings indicate that measures of muscle functional capacity (e.g. absolute strength) are likely to be more appropriate diagnostic criteria for sarcopenia than are measures of muscle size, and also highlight the importance of explicitly considering sex when defining and diagnosing sarcopenia.

## Supplementary Files

### Supplementary Data 1 Excluded participants

#### Participant exclusion flowcharts

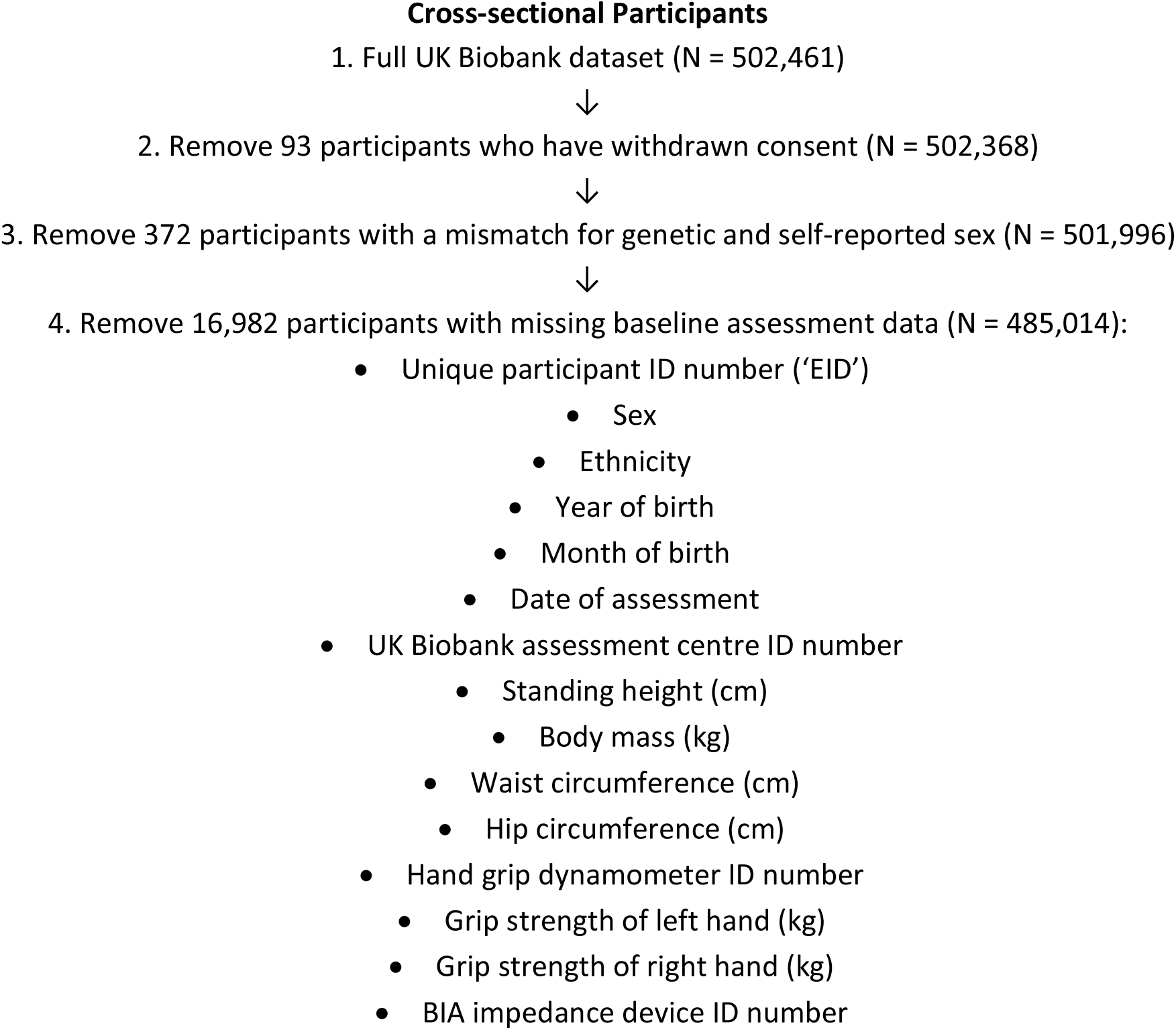
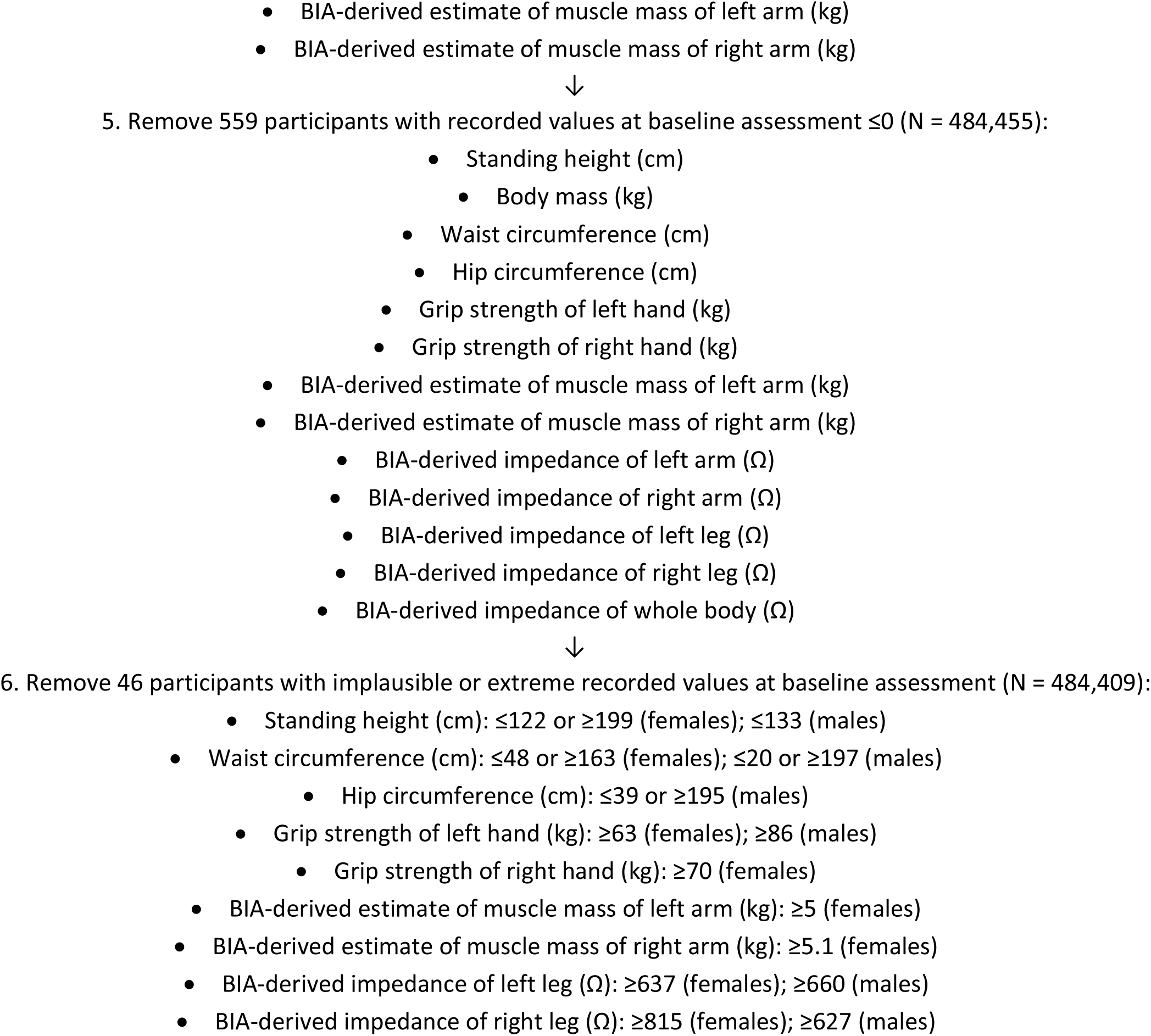
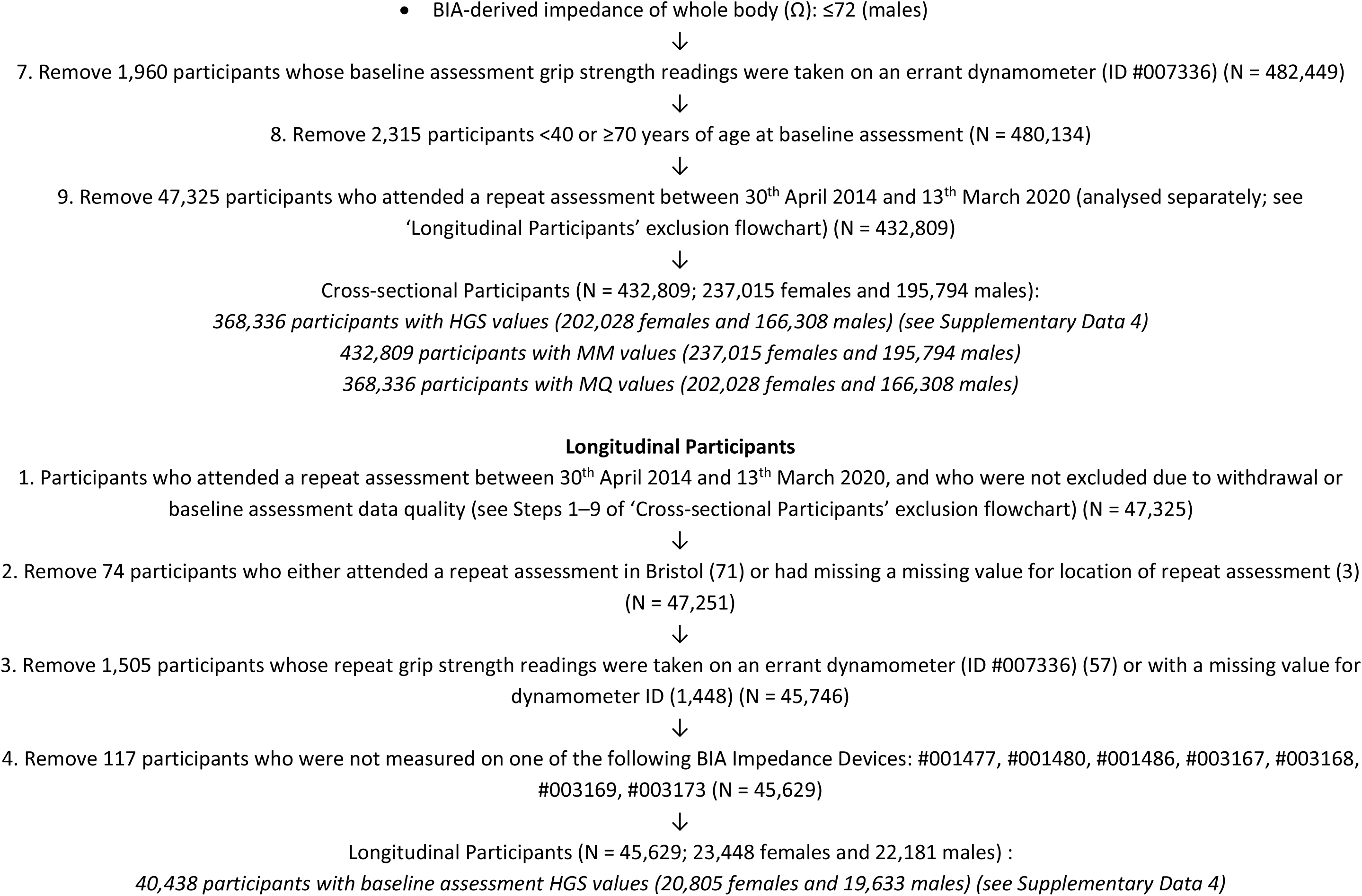
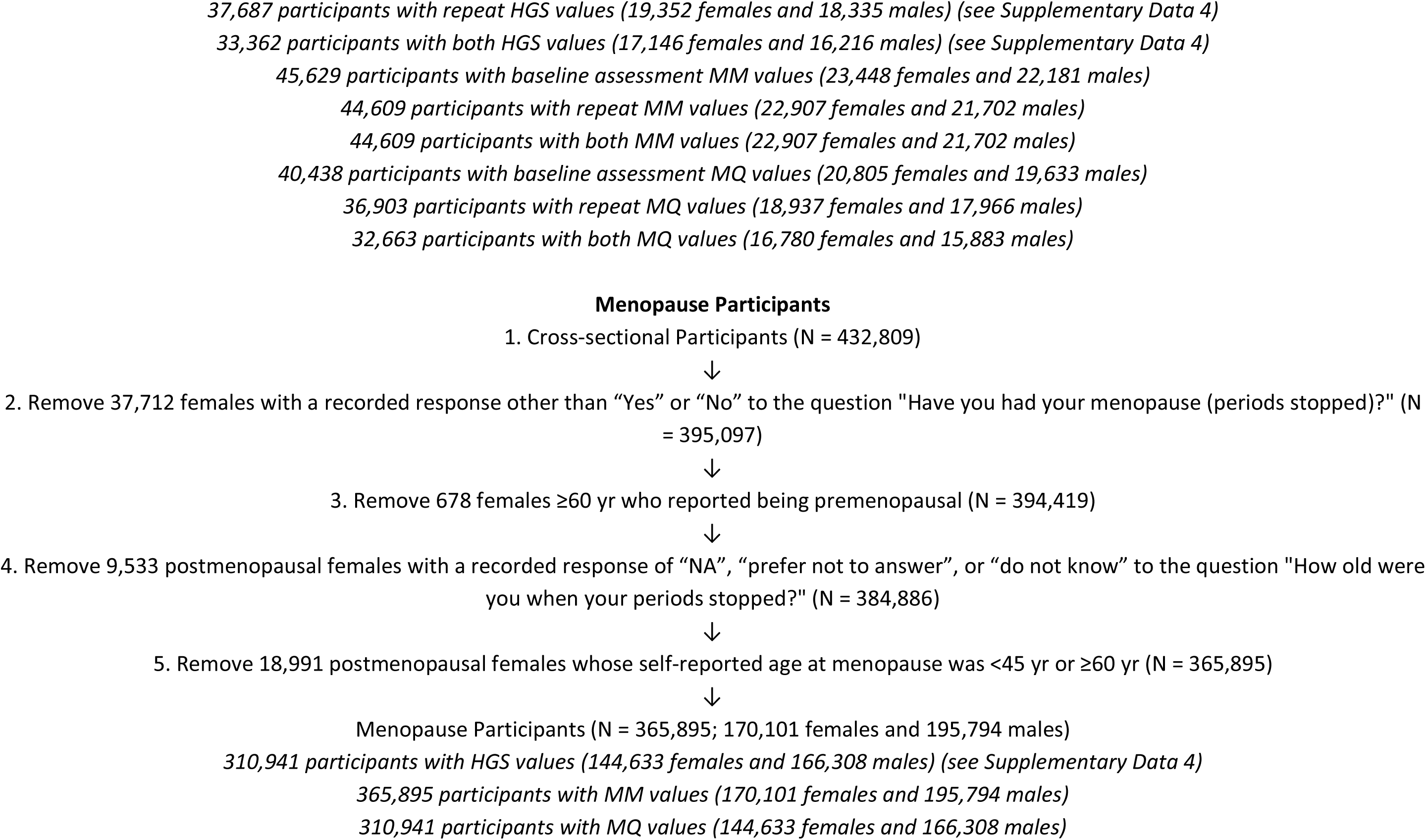

#### Characteristics of excluded participants

**Table SD1.I.**
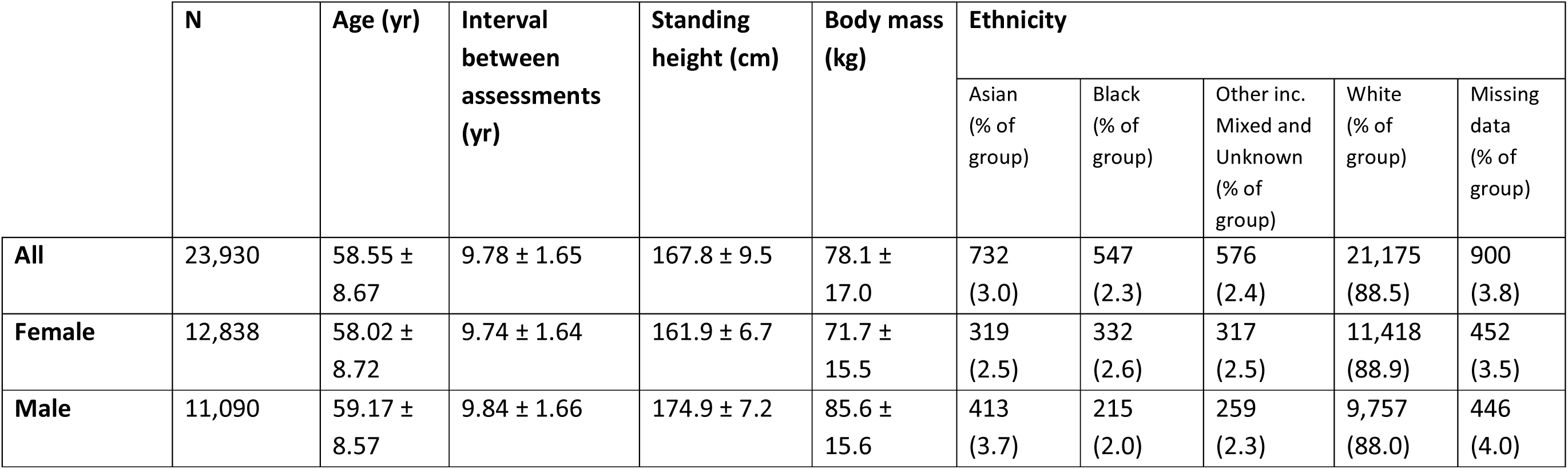
Characteristics of the 23,930 participants excluded from this study. Values are from baseline assessment, with the exception of ‘Interval between assessments’. Data are either simple counts (‘N’ and ‘Ethnicity’), or mean ± the standard deviation. Discrepancies in simple counts of All participants vs sex-stratified counts result from missing ‘sex’ values for two participants.

### Supplementary Data 2 Regression analysis

#### Multiple linear regression

Total and annualised changes/differences (in unit or percentage terms) can be calculated using the models reported below and the formulae described in the ‘Calculations and analyses’ section of the Methods. *P* values are those of age-sex interaction terms, with *P* < 0.05 taken as evidence of a sex difference. ‘Difference in slopes’ (β) is calculated as *female – male*, with negative values therefore indicating a greater decline with age in males.

**HGS ∼ age + sex + age*sex**

1. Cross-sectional Participants: Females: *HGS = 38.338 − 0.258·age* 40 yr: 28.0 kgf; 69 yr: 20.6 kgf; difference (40 vs 69 yr): *−*7.4 kgf (−26.4%); inferred annual change (40–69 yr): −0.26 kgf·yr^−1^ (−1.05 %·yr^−1^) Males: HGS = *57.026 − 0.301·age* 40 yr: 45.0 kgf; 69 yr: 36.3 kgf; difference (40 vs 69 yr): *−*8.7 kgf (−19.3%); inferred annual change (40–69 yr): −0.30 kgf·yr^−1^ (−0.74 %·yr^−1^) β: −0.043 ± 0.006 kgf·yr^−1^ (*P* < 0.001)
2. Longitudinal Participants: Females: *HGS = 38.905 − 0.248·age* 40 yr: 29.0 kgf; 80 yr: 19.1 kgf; difference (40 vs 80 yr): *−*9.9 kgf (−34.1%); inferred annual change (40–80 yr): −0.25 kgf·yr^−1^ (−1.04 %·yr^−1^) Males: *HGS = 60.426 – 0.338·age* 40 yr: 46.9 kgf; 80 yr: 33.4 kgf; difference (40 vs 80 yr): *−*13.5 kgf (−28.8%); inferred annual change (40–80 yr): −0.34 kgf·yr^−1^ (−0.85 %·yr^−1^) β: −0.090 ± 0.011 kgf·yr^−1^ (*P* < 0.001)
3. Matched Cross-sectional Participants: Females: *HGS = 40.959 – 0.274·age* 40 yr: 30.0 kgf; 69 yr: 22.0 kgf; difference (40 vs 69 yr): *−*8.0 kgf (−26.7%); inferred annual change (40–69 yr): −0.28 kgf·yr^−1^ (−1.06 %·yr^−1^) Males: *HGS = 53.041 – 0.280·age* 40 yr: 41.9 kgf; 69 yr: 33.7 kgf; difference (40 vs 69 yr): *−*8.2 kgf (−19.6%); inferred annual change (40–69 yr): −0.28 kgf·yr^−1^ (−0.75 %·yr^−1^) β: −0.006 ± 0.010 kgf·yr^−1^ (*P* = 0.29)
4. Matched Longitudinal Participants: Females: *HGS = 41.230 – 0.262·age* 40 yr: 30.8 kgf; 80 yr: 20.3 kgf; difference (40 vs 80 yr): *−*10.5 kgf (−34.1%); inferred annual change (40–80 yr): −0.26 kgf·yr^−1^ (−1.04 %·yr^−1^) Males: *HGS = 55.684 – 0.301·age* 40 yr: 43.7 kgf; 80 yr: 31.6 kgf; difference (40 vs 80 yr): *−*12.1 kgf (−27.7%); inferred annual change (40–80 yr): −0.30 kgf·yr^−1^ (−0.81 %·yr^−1^) β: −0.039 ± 0.022 kgf·yr^−1^ (*P* < 0.001)

**MM ∼ age + sex + age*sex**

1. Cross-sectional Participants: Females: *MM = 2.306 – 0.004·age* 40 yr: 2.16 kg; 69 yr: 2.05 kg; difference (40 vs 69 yr): *−*0.11 kg (−5.1%); inferred annual change (40–69 yr): −0.004 kg·yr^−1^ (−0.18 %·yr^−1^) Males: *MM = 4.303 – 0.013·age* 40 yr: 3.76 kg; 69 yr: 3.37 kg; difference (40 vs 69 yr): *−*0.39 kg (−10.4%); inferred annual change (40–69 yr): −0.013 kg·yr^−1^ (−0.38 %·yr^−1^) β: −0.010 ± 0.0003 kg·yr^−1^ (*P* < 0.001)
2. Longitudinal Participants: Females: *MM = 2.333 – 0.004·age* 40 yr: 2.16 kg; 80 yr: 1.98 kg; difference (40 vs 80 yr): *−*0.18 kg (−8.3%); inferred annual change (40–80 yr): −0.005 kg·yr^−1^ (−0.22 %·yr^−1^) Males: *MM = 4.420 – 0.016·age* 40 yr: 3.79 kg; 80 yr: 3.15 kg; difference (40 vs 80 yr): *−*0.64 kg (−16.9%); inferred annual change (40–80 yr): −0.016 kg·yr^−1^ (−0.46 %·yr^−1^) β: −0.011 ± 0.0005 kg·yr^−1^ (*P* < 0.001)
3. Matched Cross-sectional Participants Females: *MM = 2.540 – 0.004·age* 40 yr: 2.36 kg; 69 yr: 2.24 kg; difference (40 vs 69 yr): *−*0.12 kg (−5.1%); inferred annual change (40–69 yr): −0.004 kg·yr^−1^ (−0.18 %·yr^−1^) Males: *MM = 3.989 – 0.013·age* 40 yr: 3.47 kg; 69 yr: 3.10 kg; difference (40 vs 69 yr): *−*0.37 kg (−10.7%); inferred annual change (40–69 yr): −0.013 kg·yr^−1^ (−0.39 %·yr^−1^) β: −0.009 ± 0.0005 kg·yr^−1^ (*P* < 0.001)
4. Matched Longitudinal Participants: Females: *MM = 2.576 – 0.005·age* 40 yr: 2.36 kg; 80 yr: 2.14 kg; difference (40 vs 80 yr): *−*0.22 kg (−9.3%); inferred annual change (40–80 yr): −0.006 kg·yr^−1^ (−0.24 %·yr^−1^) Males: *MM = 4.049 – 0.015·age* 40 yr: 3.46 kg; 80 yr: 2.88 kg; difference (40 vs 80 yr): *−*0.58 kg (−16.8%); inferred annual change (40–80 yr): −0.015 kg·yr^−1^ (−0.46 %·yr^−1^) β: −0.009 ± 0.0009 kg·yr^−1^ (*P* < 0.001)

**MQ ∼ age + sex + age*sex**

1. Cross-sectional Participants: Females: *MQ = 447 – 2.947·age* 40 yr: 329 au; 69 yr: 244 au; difference (40 vs 69 yr): *−*85 au (−25.8%); inferred annual change (40–69 yr): −2.9 au·yr^−1^ (−1.03 %·yr^−1^) Males: *MQ = 391 – 1.419·age* 40 yr: 334 au; 69 yr: 293 au; difference (40 vs 69 yr): *−*41 au (−12.3%); inferred annual change (40–69 yr): −1.4 au·yr^−1^ (−0.45 %·yr^−1^) β: 1.528 ± 0.057 au·yr^−1^ (*P* < 0.001)
2. Longitudinal Participants: *MQ = 447 – 2.626·age* 40 yr: 342 au; 80 yr: 237 au; difference (40 vs 80 yr): *−*105 au (−30.7%); inferred annual change (40–80 yr): −2.6 au·yr^−1^ (−0.91 %·yr^−1^) Males: *MQ = 405 – 1.417·age* 40 yr: 349 au; 80 yr: 292 au; difference (40 vs 80 yr): *−*57 au (−16.3%); inferred annual change (40–80 yr): −1.4 au·yr^−1^ (−0.44 %·yr^−1^) β: 1.208 ± 0.112 au·yr^−1^ (*P* < 0.001)
3. Matched Cross-sectional Participants: Females: *MQ = 469 – 2.991·age* 40 yr: 349 au; 69 yr: 262 au; difference (40 vs 69 yr): *−*87 au (−24.9%); inferred annual change (40–69 yr): −3.0 au·yr^−1^ (−0.98 %·yr^−1^) Males: *MQ = 368 – 1.371·age* 40 yr: 313 au; 69 yr: 273 au; difference (40 vs 69 yr): *−*40 au (−12.8%); inferred annual change (40–69 yr): −1.4 au·yr^−1^ (−0.47 %·yr^−1^) β : 1.620 ± 0.103 au·yr^−1^ (*P* < 0.001)
4. Matched Longitudinal Participants: Females: *MQ = 470 – 2.706·age* 40 yr: 361 au; 80 yr: 253 au; difference (40 vs 80 yr): *−*108 au (−29.9%); inferred annual change (40–80 yr): −2.7 au·yr^−1^ (−0.88 %·yr^−1^) Matched Longitudinal Participants (males): *MQ = 381 – 1.309·age* 40 yr: 329 au; 80 yr: 277 au; difference (40 vs 80 yr): *−*52 au (−15.8%); inferred annual change (40–80 yr): −1.3 au·yr^−1^ (−0.43 %·yr^−1^) β : 1.396 ± 0.221 au·yr^−1^ (*P* < 0.001)

### Generalised additive mixed models

The generalised additive mixed models (GAMMs) below are presented in the syntax of the ‘mgcv’ R package’s *bam* function (see main manuscript for references). <u>Underlining</u> denotes terms included in sex- or group-specific models only, while the remaining terms are included in both global and sex- or group-specific models. ‘Dynamometer instance’ denotes a specific dynamometer over a pre-determined usage period: for example, four discrete usage periods (‘instances’) were identified for dynamometer #000194, each of which was then coded as a unique ‘device’ (i.e. ‘#000194_0’–’#000194_3’) for modelling purposes (Supplementary Data 4). ‘Device day of use’ is a value that equals zero on the earliest assessment date of a BIA impedance device/dynamometer instance, and increments by one for each subsequent calendar day (Supplementary Data 4). *s(x, bs = "re")* indicates that covariate *x* was modelled as a random effect. Lower AIC scores indicate better model fit, and larger ΔAICs therefore indicate stronger support for the model with the lower AIC score. Each ΔAIC was converted to a Akaike weight-derived percentage probability (‘prob_AW_’) of the lower-AIC model being the better of the two approximating models within the candidate set (Methods). Units are as follows: age—yr; CSA—au; HGS—kgf; MM—kg; MQ—au; PMSH—cm. Units for DXA-derived estimates are kg for muscle mass and au for muscle quality. See Supplementary Data 7 for more information on the model selection procedure for controlling for body size within GAMMs. Note that a small number of models do not incorporate random effects (e.g. MM Model 7; Supplementary Fig. 5a–f), and are thus technically generalised additive models (GAMs) rather than GAMMs; however, the term ‘GAMM’ is employed throughout this study for the sake of consistency and simplicity.

**HGS model 1—Cross-sectional Participants (Fig. 1a)**

*HGS ∼ sex + s(age) + <u>s(age, by = sex)</u> + ethnicity + season of attendance + s(UKB Assessment Centre, bs = "re") + s(dynamometer instance, bs = "re") + s(dynamometer instance, device day of use, bs = "re")*

**AIC_GLOBAL_** 2,441,316; **AIC_SEX-SPECIFIC_** 2,440,783; **ΔAIC** 533; **prob_AW_** >99.9%

**HGS model 2—Longitudinal Participants (Fig. 1b)**

*HGS ∼ sex + s(age) + <u>s(age, by = sex)</u> + ethnicity + season of attendance + s(UKB Assessment Centre, bs = "re") + s(dynamometer instance, bs = “re”) + s(dynamometer instance, device day of use, bs = "re")* + *s(participant ID, bs = "re")*

**AIC_GLOBAL_** 439,223; **AIC_SEX-SPECIFIC_** 437,287; **ΔAIC** 1,936; **prob_AW_** >99.9%

**HGS model 3—Cross-sectional Participants (controlling for body size) (Supplementary Fig. 1a)**

*HGS ∼ sex + s(age) + <u>s(age, by = sex)</u> + ethnicity + season of attendance + s(UKB Assessment Centre, bs = "re") + s(dynamometer instance, bs = "re") + s(dynamometer instance, device day of use, bs = "re") + s(standing height) + s(body mass) + s(standing height, by = sex) + s(body mass, by = sex)*

**AIC_GLOBAL_** 2,403,705; **AIC_SEX-SPECIFIC_** 2,403,315; **ΔAIC** 390; **prob_AW_** >99.9%

**HGS model 4—Longitudinal Participants (controlling for body size) (Supplementary Fig. 1b)**

*HGS ∼ sex + s(age) + <u>s(age, by = sex)</u> + ethnicity + season of attendance + s(UKB Assessment Centre, bs = "re") + s(dynamometer instance, bs = "re") + s(dynamometer instance, device day of use, bs = "re") + s(participant ID, bs = "re") + s(standing height) + s(body mass) + s(standing height, by = sex) + s(body mass, by = sex)*

**AIC_GLOBAL_** 438,383; **AIC_SEX-SPECIFIC_** 436,735; **ΔAIC** 1,648; **prob_AW_** >99.9%

**HGS model 5—Matched Cross-sectional Participants (Fig. 1c)**

*HGS ∼ sex + s(age) + <u>s(age, by = sex)</u> + ethnicity + season of attendance + s(UKB Assessment Centre, bs = "re") + s(dynamometer instance, bs = “re”) + s(dynamometer instance, device day of use, bs = "re")*

**AIC_GLOBAL_** 690,961; **AIC_SEX-SPECIFIC_** 690,900; **ΔAIC** 61; **prob_AW_** >99.9%

**HGS model 6—Matched Longitudinal Participants (Fig. 1d)**

*HGS ∼ sex + s(age) + <u>s(age, by = sex)</u> + ethnicity + season of attendance + s(UKB Assessment Centre, bs = "re") + s(dynamometer instance, bs = “re”) + s(dynamometer instance, device day of use, bs = "re")* + *s(participant ID, bs = "re")*

**AIC_GLOBAL_** 103,641; **AIC_SEX-SPECIFIC_** 103,412; **ΔAIC** 229; **prob_AW_** >99.9%

**MM model 1—Cross-sectional Participants (Fig. 2a)**

*MM ∼ sex + s(age) + <u>s(age, by = sex)</u> + ethnicity + season of attendance + s(UKB Assessment Centre, bs = "re") + s(BIA impedance device, bs = “re”) + s(BIA impedance device, device day of use, bs = "re")*

**AIC_GLOBAL_** 373,361; **AIC_SEX-SPECIFIC_** 368,198; **ΔAIC** 5,163; **prob_AW_** >99.9%

**MM model 2—Longitudinal Participants (Fig. 2b)**

*MM ∼ sex + s(age) + <u>s(age, by = sex)</u> + ethnicity + season of attendance + s(UKB Assessment Centre, bs = "re") + s(BIA impedance device, bs = “re”) + s(BIA impedance device, device day of use, bs = "re")* + *s(participant ID, bs = "re")*

**AICGLOBAL** −90,132; **AIC_SEX-SPECIFIC_** −100,374.; **ΔAIC** 10,242; **prob_AW_** >99.9%

**MM model 3—Cross-sectional Participants (controlling for body size) (Supplementary Fig. 2a)**

*MM ∼ sex + s(age) + <u>s(age, by = sex)</u> + ethnicity + season of attendance + s(UKB Assessment Centre, bs = "re") + s(BIA impedance device, bs = “re”) + s(BIA impedance device, device day of use, bs = "re") + s(standing height) + s(body mass) + s(standing height, by = sex) + s(body mass, by = sex)*

**AIC_GLOBAL_** –162,851; **AIC_SEX-SPECIFIC_** −174,605; **ΔAIC** 11,754; **prob_AW_** >99.9%

**MM model 4—Longitudinal Participants (controlling for body size) (Supplementary Fig. 2b)**

*MM ∼ sex + s(age) + <u>s(age, by = sex)</u>* + *ethnicity + season of attendance + s(UKB Assessment Centre, bs = "re") + s(BIA impedance device, bs = “re”) + s(BIA impedance device, device day of use, bs = "re")* + *s(participant ID, bs = "re") + s(standing height) + s(body mass) + s(standing height, by = sex) + s(body mass, by = sex)*

**AIC_GLOBAL_** −121,199; **AIC_SEX-SPECIFIC_** −132,636; **ΔAIC** 11,437; **prob_AW_** >99.9%

**MM model 5—Matched Cross-sectional Participants (Fig. 2c)**

*MM ∼ sex + s(age) + <u>s(age, by = sex)</u> + ethnicity + season of attendance + s(UKB Assessment Centre, bs = "re") + s(BIA impedance device, bs = “re”) + s(BIA impedance device, device day of use, bs = "re")*

**AIC_GLOBAL_** 80,146; **AIC_SEX-SPECIFIC_** 78,585; **ΔAIC** 1,561; **prob_AW_** >99.9%

**MM model 6—Matched Longitudinal Participants (Fig. 2d)**

*MM ∼ sex + s(age) + <u>s(age, by = sex)</u> + ethnicity + season of attendance + s(UKB Assessment Centre, bs = "re") + s(BIA impedance device, bs = “re”) + s(BIA impedance device, device day of use, bs = "re")* + *s(participant ID, bs = "re")*

**AIC_GLOBAL_** −23,390; **AIC_SEX-SPECIFIC_** −25,342; **ΔAIC** 1,952; **prob_AW_** >99.9%

**MM model 7—4,253 Longitudinal Participants ≥44.5 and ≤78 yr subjected to a DXA scan (N = 2,224 [F], 2,029 [M]) (Fig. 2e)**

(NB DXA measurement device and UKB Assessment Centre were not included as covariates in this model, as all DXA scans were performed at the Cheadle Assessment Centre, and no information was available on whether multiple DXA devices were utilised)

*DXA-derived estimate of mean arm muscle mass ∼ sex + s(age) + <u>s(age, by = sex)</u> + ethnicity + season of attendance*

**AIC_GLOBAL_** 4,651.8; **AIC_SEX-SPECIFIC_** 4,597.5; **ΔAIC** 54.3; **prob_AW_** >99.9%

**MQ model 1—Cross-sectional Participants (Fig. 3a)**

*MQ ∼ sex + s(age) + s(age, by = sex) + ethnicity + season of attendance + s(UKB Assessment Centre, bs = "re") + s(dynamometer instance, bs = "re") + s(dynamometer instance, device day of use, bs = "re") + s(BIA impedance device, bs = “re”) + s(BIA impedance device, device day of use, bs = "re")* **AIC_GLOBAL_** 4,145,095; **AIC_SEX-SPECIFIC_** 4,141,887; **ΔAIC** 3,208; **probAW** >99.9%

**MQ model 2—Longitudinal Participants (Fig. 3b)**

*MQ ∼ sex + s(age) + <u>s(age, by = sex)</u> + ethnicity + season of attendance + s(UKB Assessment Centre, bs = "re") + s(dynamometer instance, bs = “re”) + s(dynamometer instance, device day of use, bs = "re") + s(BIA impedance device, bs = “re”) + s(BIA impedance device, device day of use, bs = "re")* + *s(participant ID, bs = "re")*

**AIC_GLOBAL_** 794,360; **AIC_SEX-SPECIFIC_** 793,770; **ΔAIC** 590; **prob_AW_** >99.9%

**MQ model 3—Cross-sectional Participants (controlling for body size) (Supplementary Fig. 6a)**

*MQ ∼ sex + s(age) + <u>s(age, by = sex)</u> + ethnicity + season of attendance + s(UKB Assessment Centre, bs = "re") + s(dynamometer instance, bs = “re”) + s(dynamometer instance, device day of use, bs = "re") + s(BIA impedance device, bs = “re”) + s(BIA impedance device, device day of use, bs = "re") + s(standing height) + s(body mass) + s(standing height, by = sex) + s(body mass, by = sex)*

**AIC_GLOBAL_** 4,036,496; **AIC_SEX-SPECIFIC_** 4,033,275; **ΔAIC** 3,221; **prob_AW_** >99.9%

**MQ model 4—Longitudinal Participants (controlling for body size) (Supplementary Fig. 6b)**

*MQ ∼ sex + s(age) + <u>s(age, by = sex)</u> + ethnicity + season of attendance + s(UKB Assessment Centre, bs = "re") + s(dynamometer instance, bs = “re”) + s(dynamometer instance, device day of use, bs = "re") + s(BIA impedance device, bs = “re”) + s(BIA impedance device, device day of use, bs = "re")* + *s(participant ID, bs = "re") + s(standing height) + s(body mass) + s(standing height, by = sex) + s(body mass, by = sex)*

**AIC_GLOBAL_** 793,296; **AIC_SEX-SPECIFIC_** 792,871; **ΔAIC** 425; **prob_AW_** >99.9%\

**MQ model 5—Matched Cross-sectional Participants (Fig. 3c)**

*MQ ∼ sex + s(age) + s(age, by = sex) + ethnicity + season of attendance + s(UKB Assessment Centre, bs = "re") + s(dynamometer instance, bs = “re”) + s(dynamometer instance, device day of use, bs = "re") + s(BIA impedance device, bs = “re”) + s(BIA impedance device, device day of use, bs = "re")* **AIC_GLOBAL_** 1,174,546; **AIC_SEX-SPECIFIC_** 1,173,498; **ΔAIC** 1,048; **prob_AW_** >99.9%\**MQ model 6—Matched Longitudinal Participants (Fig. 3d)**

*MQ ∼ sex + s(age) + <u>s(age, by = sex)</u> + ethnicity + season of attendance + s(UKB Assessment Centre, bs = "re") + s(dynamometer instance, bs = “re”) + s(dynamometer instance, device day of use, bs = "re") + s(BIA impedance device, bs = “re”) + s(BIA impedance device, device day of use, bs = "re") + s(participant ID, bs = "re")*

**AIC_GLOBAL_** 188,917; **AIC_SEX-SPECIFIC_** 188,701; **ΔAIC** 216; **prob_AW_** >99.9%

**MQ model 7—3,972 Longitudinal Participants ≥45.4 and ≤78 yr subjected to a DXA scan (N = 2,061 [F], 1,911 [M]) (Fig. 3e)**

(NB DXA measurement device and UKB Assessment Centre were not included as covariates in this model, as all DXA scans were performed at the Cheadle Assessment Centre, and no information was available on whether multiple DXA devices were utilised)

*DXA-derived estimate of mean arm muscle quality ∼ sex + s(age) + s(age, by = sex) + ethnicity + season of attendance + s(dynamometer instance, bs = "re") + s(dynamometer instance, device day of use, bs = "re")*

**AIC_GLOBAL_** 44,188; **AIC_SEX-SPECIFIC_** 44,165; **ΔAIC** 23; **prob_AW_** >99.9%

**MQ’ model 1—Cross-sectional Participants (Supplementary Fig. 8a)**

*MQ’ ∼ sex + s(age) + s(age, by = sex) + ethnicity + season of attendance + s(UKB Assessment Centre, bs = "re") + s(dynamometer instance, bs = "re") + s(dynamometer instance, device day of use, bs = "re") + s(BIA impedance device, bs = “re”) + s(BIA impedance device, device day of use, bs = "re")* **AIC_GLOBAL_** 1,712,989; **AIC_SEX-SPECIFIC_** 1,708,939; **ΔAIC** 4,050; **prob_AW_** >99.9%

**MQ’ model 2—Longitudinal Participants (Supplementary Fig. 8b)**

*MQ’ ∼ sex + s(age) + <u>s(age, by = sex)</u> + ethnicity + season of attendance + s(UKB Assessment Centre, bs = "re") + s(dynamometer instance, bs = “re”) + s(dynamometer instance, device day of use, bs = "re") + s(BIA impedance device, bs = “re”) + s(BIA impedance device, device day of use, bs = "re")* + *s(participant ID, bs = "re")*

**AIC_GLOBAL_** 292,466; **AIC_SEX-SPECIFIC_** 291,631; **ΔAIC** 835; **prob_AW_** >99.9%

**MQ’ model 3—Matched Cross-sectional Participants (Supplementary Fig. 8c)**

*MQ’ ∼ sex + s(age) + s(age, by = sex) + ethnicity + season of attendance + s(UKB Assessment Centre, bs = "re") + s(dynamometer instance, bs = “re”) + s(dynamometer instance, device day of use, bs = "re") + s(BIA impedance device, bs = “re”) + s(BIA impedance device, device day of use, bs = "re")* **AIC_GLOBAL_** 481,540; **AIC_SEX-SPECIFIC_** 480,540; **ΔAIC** 1,000; **prob_AW_** >99.9%

**MQ’ model 4—Matched Longitudinal Participants (Supplementary Fig. 8d)**

*MQ’ ∼ sex + s(age) + <u>s(age, by = sex)</u> + ethnicity + season of attendance + s(UKB Assessment Centre, bs = "re") + s(dynamometer instance, bs = “re”) + s(dynamometer instance, device day of use, bs = "re") + s(BIA impedance device, bs = “re”) + s(BIA impedance device, device day of use, bs = "re") + s(participant ID, bs = "re")*

**AIC_GLOBAL_** 68,418; **AIC_SEX-SPECIFIC_** 68,235; **ΔAIC** 183; **prob_AW_** >99.9%

**MQ’ model 5—3,972 Longitudinal Participants ≥45.4 and ≤78 yr subjected to a DXA scan (N = 2,061 [F], 1,911 [M]) (Supplementary Fig. 8e)**

(NB DXA measurement device and UKB Assessment Centre were not included as covariates in this model, as all DXA scans were performed at the Cheadle Assessment Centre, and no information was available on whether multiple DXA devices were utilised. Dynamometer instance was modelled as a fixed effect rather than a random effect)

*DXA-derived estimate of mean arm muscle quality ∼ sex + s(age) + s(age, by = sex) + ethnicity + season of attendance + dynamometer instance + s(device day of use, by=dynamometer instance)*

**AIC_GLOBAL_** 17,817; **AIC_SEX-SPECIFIC_** 17,789; **ΔAIC** 28; **prob_AW_** >99.9%\

**HGS menopause models 1–6—Menopause Participants (controlling for body size)**

*HGS ∼ group* + s(age) + <u>s(age, by = group)</u>* + *ethnicity + season of attendance + s(UKB Assessment Centre, bs = "re") + s(dynamometer instance, bs = "re") + s(dynamometer instance, device day of use, bs = "re") + s(standing height) + s(body mass) + s(standing height, by = group) + s(body mass, by = group)*

*1. Premenopausal females vs Postmenopausal females: **AIC_GLOBAL_** 875,663.5; **AIC_GROUP-SPECIFIC_** 875,654.8; **ΔAIC** 8.7; **prob_AW_** 98.7%

2. Postmenopausal females vs Males: **AIC_GLOBAL_** 1,761,165; **AIC_GROUP-SPECIFIC_** 1,760,775; **ΔAIC** 390; **prob_AW_** >99.9%

3. Premenopausal females vs Males: **AIC_GLOBAL_** 1,431,264; **AIC_GROUP-SPECIFIC_** 1,431,249; **ΔAIC** 15; **prob_AW_** >99.9%

4. Postmenopausal females (HRT) vs Postmenopausal females (no HRT): **AIC_GLOBAL_** 594,546; **AIC_GROUP-SPECIFIC_** 594,524; **ΔAIC** 22 ; **prob_AW_** >99.9% **(Fig. 4d)**

5. Postmenopausal females (HRT) vs Males: **AIC_GLOBAL_** 1,409,692; **AIC_GROUP-SPECIFIC_** 1,409,536; **ΔAIC** 156; **prob_AW_** >99.9%

6. Postmenopausal females (no HRT) vs Males: **AIC_GLOBAL_** 1,496,109; **AIC_GROUP-SPECIFIC_** 1,495,770; **ΔAIC** 339; **prob_AW_** >99.9%

**MM menopause models 1–6—Menopause Participants (controlling for body size)**

*MM ∼ group* + s(age) + <u>s(age, by = group)</u> + ethnicity + season of attendance + s(UKB Assessment Centre, bs = "re") + s(BIA impedance device, bs = “re”) + s(BIA impedance device, device day of use, bs = "re") + s(standing height) + s(body mass) + s(standing height, by = group) + s(body mass, by = group)*

*1. Premenopausal females vs Postmenopausal females: **AIC_GLOBAL_** −195,858.47; **AIC_GROUP-SPECIFIC_** −195,858.12; **ΔAIC** 0.35 ; **prob_AW_** 54.4%

2. Postmenopausal females vs Males: **AIC_GLOBAL_** −65,034; **AIC_GROUP-SPECIFIC_** −69,127; **ΔAIC** 4,093; **prob_AW_** >99.9%

3. Premenopausal females vs Males: **AIC_GLOBAL_** −19,588; **AIC_GROUP-SPECIFIC_** −20,322; **ΔAIC** 734; **prob_AW_** >99.9%

4. Postmenopausal females (HRT) vs Postmenopausal females (no HRT): **AIC_GLOBAL_** −133,509; **AIC_GROUP-SPECIFIC_** −133,531; **ΔAIC** 22; **prob_AW_** >99.9% **(Fig. 4e)**

5. Postmenopausal females (HRT) vs Males: **AIC_GLOBAL_** −16,191; **AIC_GROUP-SPECIFIC_** −17,597; **ΔAIC** 1,406; **probAW** >99.9%

6. Postmenopausal females (no HRT) vs Males: **AIC_GLOBAL_** −26,729; **AIC_GROUP-SPECIFIC_** −29,497; **ΔAIC** 2,768; **prob_AW_** >99.9%

**MQ menopause models 1–6—Menopause Participants (controlling for body size)**

*MQ ∼ group* + s(age) + <u>s(age, by = group)</u> + ethnicity + season of attendance + s(UKB Assessment Centre, bs = "re") + s(dynamometer instance, bs = "re") + s(dynamometer instance, device day of use, bs = "re") + s(BIA impedance device, bs = “re”) + s(BIA impedance device, device day of use, bs = "re") + s(standing height) + s(body mass) + s(standing height, by = group) + s(body mass, by = group)*

*1. Premenopausal females vs Postmenopausal females: **AIC_GLOBAL_** 1,582,313.9; **AIC_GROUP-SPECIFIC_** 1,582,309.0; **ΔAIC** 4.9; **prob_AW_** 92.1%

2. Postmenopausal females vs Males: **AIC_GLOBAL_** 2,897,669; **AIC_GROUP-SPECIFIC_** 2,897,331; **ΔAIC** 338; **prob_AW_** >99.9%

3. Premenopausal females vs Males: **AIC_GLOBAL_** 2,315,464; **AIC_GROUP-SPECIFIC_** 2,315,274; **ΔAIC** 190; **probAW** >99.9%

4. Postmenopausal females (HRT) vs Postmenopausal females (no HRT): **AIC_GLOBAL_** 1,079,157; **AIC_GROUP-SPECIFIC_** 1,079,142; **ΔAIC** 15; **prob_AW_** >99.9% **(Fig. 4f)**

5. Postmenopausal females (HRT) vs Males: **AIC_GLOBAL_** 2,279,056; **AIC_GROUP-SPECIFIC_** 2,278,906; **ΔAIC** 150; **prob_AW_** >99.9%

6. Postmenopausal females (no HRT) vs Males: **AIC_GLOBAL_** 2,429,909; **AIC_GROUP-SPECIFIC_** 2,429,767; **ΔAIC** 142; **prob_AW_** >99.9%

**Fig. 4a—Menopause Participants (HGS, controlling for body size)**

*HGS ∼ group* + s(age) + s(age, by = group)* + *ethnicity + season of attendance + s(UKB Assessment Centre, bs = "re") + s(dynamometer instance, bs = "re") + s(dynamometer instance, device day of use, bs = "re") + s(standing height) + s(body mass) + s(standing height, by = group) + s(body mass, by = group)*

*‘Premenopausal females’, ‘Postmenopausal females’, and ‘Males’

**Fig. 4b—Menopause Participants (MM, controlling for body size)**

*MM ∼ group* + s(age) + s(age, by = group) + ethnicity + season of attendance + s(UKB Assessment Centre, bs = "re") + s(BIA impedance device, bs = “re”) + s(BIA impedance device, device day of use, bs = "re") + s(standing height) + s(body mass) + s(standing height, by = group) + s(body mass, by = group)*

*‘Premenopausal females’, ‘Postmenopausal females’, and ‘Males’

**Fig. 4c—Menopause Participants (MQ, controlling for body size)**

*MQ ∼ group* + s(age) + s(age, by = group) + ethnicity + season of attendance + s(UKB Assessment Centre, bs = "re") + s(dynamometer instance, bs = "re") + s(dynamometer instance, device day of use, bs = "re") + s(BIA impedance device, bs = “re”) + s(BIA impedance device, device day of use, bs = "re") + s(standing height) + s(body mass) + s(standing height, by = group) + s(body mass, by = group)*

*‘Premenopausal females’, ‘Postmenopausal females’, and ‘Males’

**Supplementary Fig. 4 a,c—a) Cross-sectional Participants (females only); c) Cross-sectional Participants (males only)**

*MM ∼ s(age)* + *ethnicity + season of attendance + s(UKB Assessment Centre, bs = "re") + s(BIA impedance device, bs = “re”) + s(BIA impedance device, device day of use, bs = "re") + s(standing height) + s(body mass) + ti(standing height, age) + ti(body mass, age)*

**Supplementary Fig. 4 b,d—b) 2,224 female Longitudinal Participants ≥44.5 and ≤78 yr subjected to a DXA scan; d) 2,029 male Longitudinal Participants ≥44.5 and ≤78 yr subjected to a DXA scan**

(NB DXA measurement device and UKB Assessment Centre were not included as covariates in this model, as all DXA scans were performed at the Cheadle Assessment Centre, and no information was available on whether multiple DXA devices were utilised)

*DXA-derived estimate of mean arm muscle mass ∼ s(age) + ethnicity + season of attendance + s(standing height) + s(body mass) + ti(standing height, age) + ti(body mass, age)*

**Supplementary Fig. 7a—Cross-sectional Participants**

*MQ ∼ s(PMSH) + sex + s(age) + s(PMSH, by = sex) + s(age, by = sex) + ethnicity + season of attendance + s(UKB Assessment Centre, bs = "re") + s(dynamometer instance, bs = "re") + s(dynamometer instance, device day of use, bs = "re") + s(BIA impedance device, bs = “re”) + s(BIA impedance device, device day of use, bs = "re")*

**Supplementary Fig. 7c—Cross-sectional Participants**

*MM ∼ s(PMSH) + sex + s(age) + s(PMSH, by = sex) + s(age, by = sex) + ethnicity + season of attendance + s(UKB Assessment Centre, bs = "re") + s(BIA impedance device, bs = “re”) + s(BIA impedance device, device day of use, bs = "re")*

**Supplementary Fig. 7d—Cross-sectional Participants**

*CSA ∼ s(PMSH) + sex + s(age) + s(PMSH, by = sex) + s(age, by = sex) + ethnicity + season of attendance + s(UKB Assessment Centre, bs = "re") + s(BIA impedance device, bs = “re”) + s(BIA impedance device, device day of use, bs = "re")*

**Supplementary Fig. 7e—Cross-sectional Participants**

*HGS ∼ s(PMSH) + sex + s(age) + s(PMSH, by = sex) + s(age, by = sex) + ethnicity + season of attendance + s(UKB Assessment Centre, bs = "re") + s(dynamometer instance, bs = "re") + s(dynamometer instance, device day of use, bs = "re")*

### Supplementary Data 3 UK Biobank Data-Fields included in this study

eid; sex_f31; year_of_birth_f34; hand_grip_dynamometer_device_id_f38; impedance_device_id_f43; hand_grip_strength_left_f46; hand_grip_strength_right_f47; waist_circumference_f48; hip_circumference_f49; standing_height_f50; month_of_birth_f52; date_of_attending_assessment_centre_f53; uk_biobank_assessment_centre_f54; number_of_selfreported_cancers_f134; townsend_deprivation_index_at_recruitment_f189; number_of_daysweek_of_moderate_physical_activity_10_minutes_f884; number_of_daysweek_of_vigorous_physical_activity_10_minutes_f904; handedness_chiralitylaterality_f1707; had_menopause_f2724; ever_used_hormonereplacement_therapy_hrt_f2814; age_at_menopause_last_menstrual_period_f3581; ethnic_background_f21000; body_mass_index_bmi_f21001_0_0; weight_f21002; genetic_sex_f22001; impedance_of_whole_body_f23106; impedance_of_leg_right_f23107; impedance_of_leg_left_f23108; impedance_of_arm_right_f23109; impedance_of_arm_left_f23110; arm_predicted_mass_right_f23122; arm_fat_mass_left_f23124; arm_lean_mass_left_f23250; arm_lean_mass_right_f23254; oestradiol_f30800; testosterone_f30850

### Supplementary Data 4 Adjustment of hand grip strength readings

When plotting average HGS values against date of assessment for individual dynamometers, we observed evidence of drift in HGS readings for many devices (Fig. SD4.I). We therefore undertook to identify and remove readings taken under extreme drift, and to implement a HGS correction to minimise the impact of device drift on the remaining readings. Here, and throughout this Supplementary Data section, ‘average HGS’ refers to the average (mean) of left and right hand readings taken at a single assessment.

UK Biobank have provided dates at which dynamometers were recalibrated (Resources 2431–2433). For the purposes of quality control (QC) and adjustment, we treated measurements taken prior to a device’s earliest recalibration date, between consecutive recalibration dates, and after its latest recalibration date as separate device ‘instances’.

We first performed QC and adjustment of baseline assessment HGS readings. We excluded readings taken on devices used for <500 participants (6,840 [1.4%] of 496,905 participants with baseline assessment readings), and participants without HGS measures for both hands. We manually inspected plots for each dynamometer instance and identified clear temporal outliers and errant readings for exclusion (covering 10,495 participants in total) (e.g. Fig. SD4.I). Next, we excluded readings taken on dynamometer instances used for <250 participants (total 5,556 participants).

We quantified the drift exhibited by each dynamometer instance by fitting a regression model for average HGS that included linear, quadratic, and cubic terms for ‘device day of use’ (a value that equals zero on the earliest assessment date of a dynamometer instance, and increments by one for each subsequent calendar day). To control for participant-level differences that may influence HGS independently of systematic dynamometer effects we included sex, standing height (cm), body mass (kg), age (yr), age^2^ (yr^2^), and self-reported ethnic background ^[a]^ as covariates. We could not include Assessment Centre or season of attendance as covariates, since both are correlated with device day of use (dynamometers were periodically moved between Assessment Centres) and lead to volatile cubic fits. A small number of dynamometer instances (‘000193’, ‘000194_1’ and ‘000196_0’) were highly unbalanced in terms of sex; for these dynamometer instances the cubic model was fitted to male data only.

Average HGS values were adjusted in a two-stage process that will be described in more detail below: (i) by removing readings taken under the most extreme drift, and (ii) by re-fitting the cubic models for drift, and correcting the remaining readings (Fig. SD4.II).

i. A preliminary adjustment was made to average HGS values using the estimated model parameters. Namely, the fitted parameter values for device day of use, device day of use^2^, and device day of use^3^ were used to calculate the contribution of device drift to each individual observation; this contribution was then subtracted from the observed value to give a preliminary adjusted value. Note that this process makes an explicit assumption that dynamometers are correctly calibrated at the beginning of each dynamometer instance. Subsequently, we identified and removed the 5% of all remaining 471,611 participants subject to the largest preliminary adjustments (i.e. absolute value of adjustment). We recorded the threshold adjustment value for exclusion (5.51 kgf). The preliminary adjustment values were then discarded.
ii. We excluded any dynamometer instances with <250 observations remaining after step (i), and re-fitted the cubic regression model (identical in form to that described above) to the remaining readings for each dynamometer instance. This final model was used to generate refined adjustments for each observed average HGS reading. Finally, we performed post-adjustment filtering. First, we calculated the mean HGS across participants for each dynamometer instance, both pre- and post-adjustment. To identify and mitigate the most problematic dynamometers, we excluded dynamometer instances with the largest absolute change in mean value until 5% of participants had been excluded (10 of 204 dynamometer instances; implied threshold for dynamometer instance mean adjustment of 2.71 kgf). Second, we removed individual participants whose average HGS reading was adjusted by at least 5.51 kgf (i.e. the previously identified threshold adjustment value for exclusion).

This process of QC and adjustment resulted in adjusted baseline assessment readings for 422,364 participants. The distribution of adjustments is shown in Fig. SD4.III. The average adjustment was +0.33 kgf. In total, 168,862 participants (40.0%) had their HGS reading adjusted to a lower value and 249,847 (59.2%) had their reading adjusted to a higher value. 383,721 participants (90.9%) had their reading adjusted by <2.5 kgf.

HGS readings for the repeat assessment were adjusted using an identical process. However, due to the larger sample size at baseline assessment, we applied the participant-specific and device instance-specific adjustment thresholds calculated using the baseline assessment values (5.51 kgf and 2.71 kgf, respectively).

Note that the process of adjusting and excluding HGS readings described in this Supplementary Data was performed prior to the creation of the participant groups for our primary analyses (Supplementary Data 1). The substantial number of participants with missing adjusted HGS readings (e.g. ∼15% of Cross-sectional Participants) were retained for the MM analyses where MM estimates were available (Supplementary Data 1).

While adjustment of HGS readings inevitably altered individual HGS-age trajectories (i.e. those of most Longitudinal Participants), Figs SD4.IV and SD4.V show that cohort-level HGS-age trajectories are very consistent between the unadjusted and adjusted HGS datasets, when assessed in the Cross-sectional Participants. We conclude that the general cohort-level trends in HGS and MQ reported in this study are not biased by our HGS adjustments.

**Figure SD4.I.**
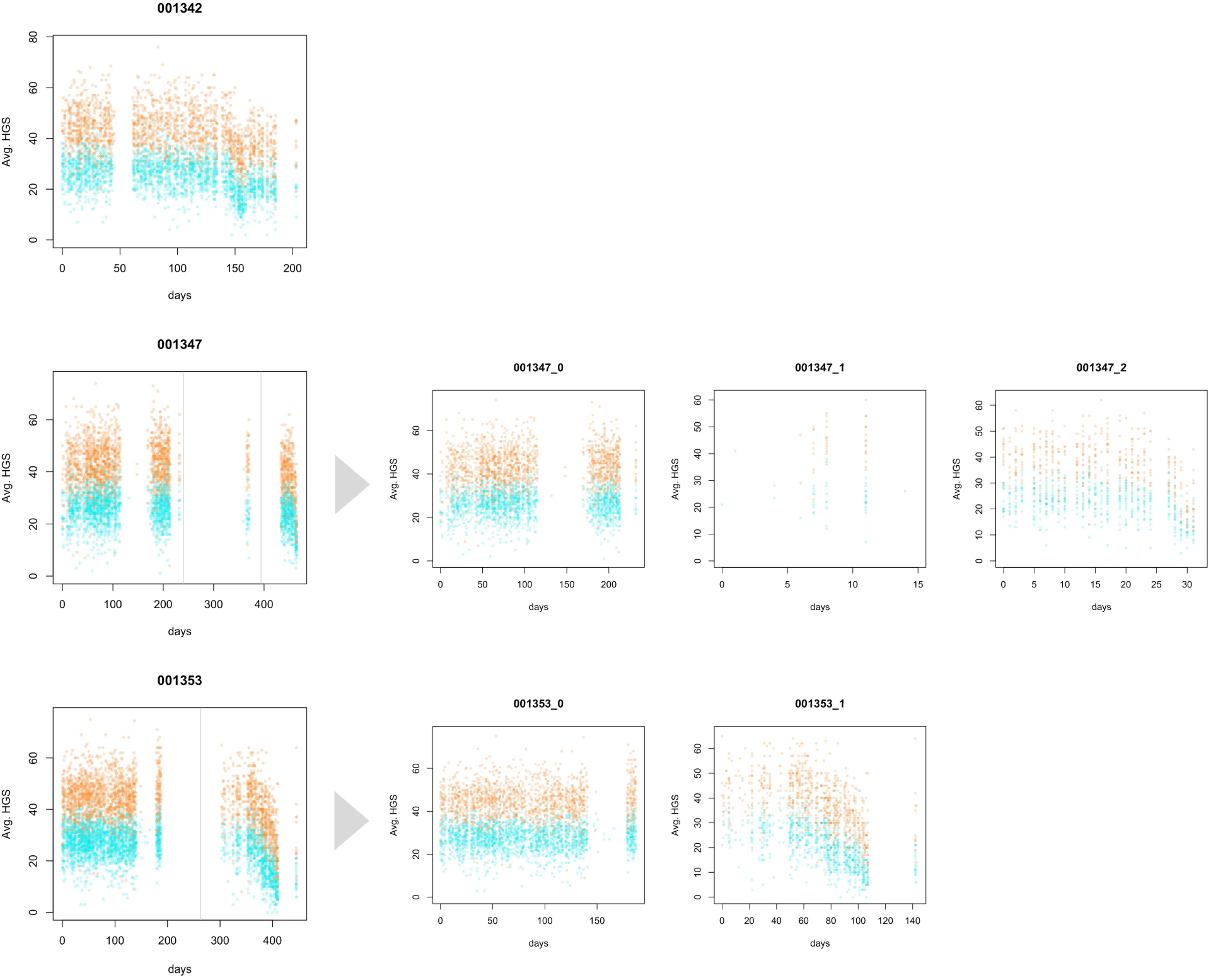
Example of dynamometer drift. Plots of unadjusted average hand grip strength (HGS) values for three illustrative dynamometer devices (#001342, #001347, and #001353) are shown in the left column. Each point represents a participant; *y*-axis, average HGS in kgf; *x*-axis, ‘device day of use’ (see Supplementary Data 2); point colour, sex (female = teal; male = orange). Vertical grey lines indicate dates on which UK Biobank recorded a calibration check (and if necessary, recalibration) of the device. Where calibration checks occurred, we split the data into ‘dynamometer instances’ with independent ‘device days of use’. This is illustrated by the grey arrows and the plots on their right, which display the same data, but as separate dynamometer instances. Note that for device #001342 there were no recorded calibration checks, and all data is therefore treated as a single dynamometer instance. Dynamometer instances #001347_1 and #001353_1 were excluded from subsequent analysis (as described in detail in the text of this Supplementary Data).

**Figure SD4.II.**
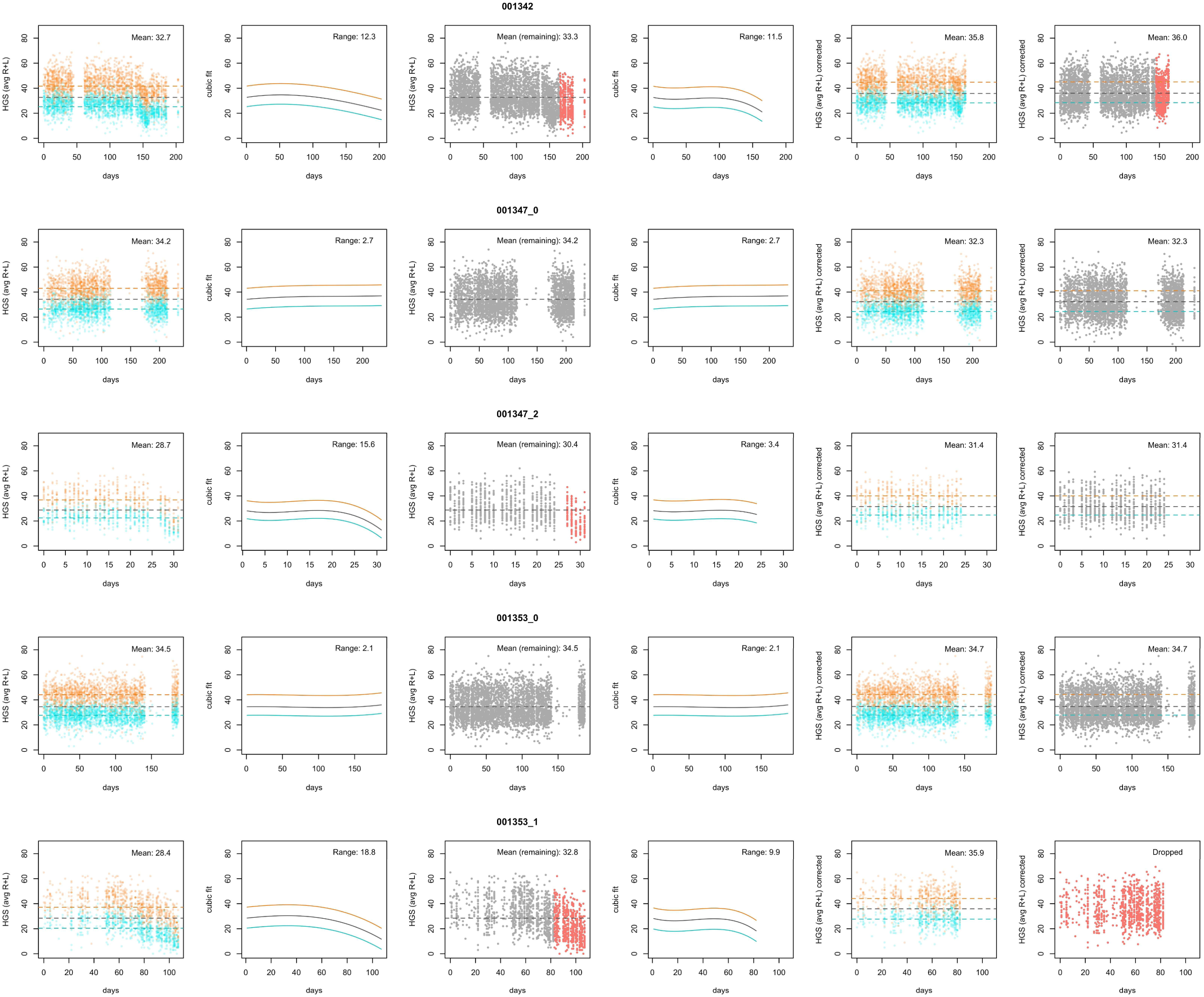
Illustration of hand grip strength quality control and adjustment. These plots illustrate the subsequent steps of the quality control (QC) and adjustment process for each of the dynamometer instances shown in Fig. SD4.I (excluding dynamometer instance #001347_1, which was excluded due to low numbers). **First column** Unadjusted data, plotted as described for Fig. SD4.I. Dashed lines indicate mean observed average hand grip strength (HGS) values (female = teal; male = orange; grey = all). **Second column** Preliminary cubic fit to the observed values. Indicative lines are shown for males (orange), females (teal), and all (grey), but since a single cubic model is fitted per dynamometer instance, these represent the same line shifted in position on the *y*-axis. **Third column** Unadjusted average HGS values are plotted again, coloured according to whether they are taken forward to the final cubic model; red points are excluded due to being among the 5% of participants (across all dynamometer instances) with the largest absolute preliminary adjustment value. **Fourth column** Refined cubic fit to the observed values, based on remaining participants only. **Fifth column** Average HGS values plotted after adjustment, but before the final filtering step. **Sixth column** Adjusted average HGS values are plotted again, coloured according to whether they are retained in the final dataset; red points are excluded due to being measured in a dynamometer instance among the top 5% according to overall mean adjustment (e.g. #001353_1 here) or due to exhibiting individual adjustments larger than the threshold identified from the preliminary fit (e.g. #001342 here). A more detailed description is provided in the text of this Supplementary Data.

**Figure SD4.III.**
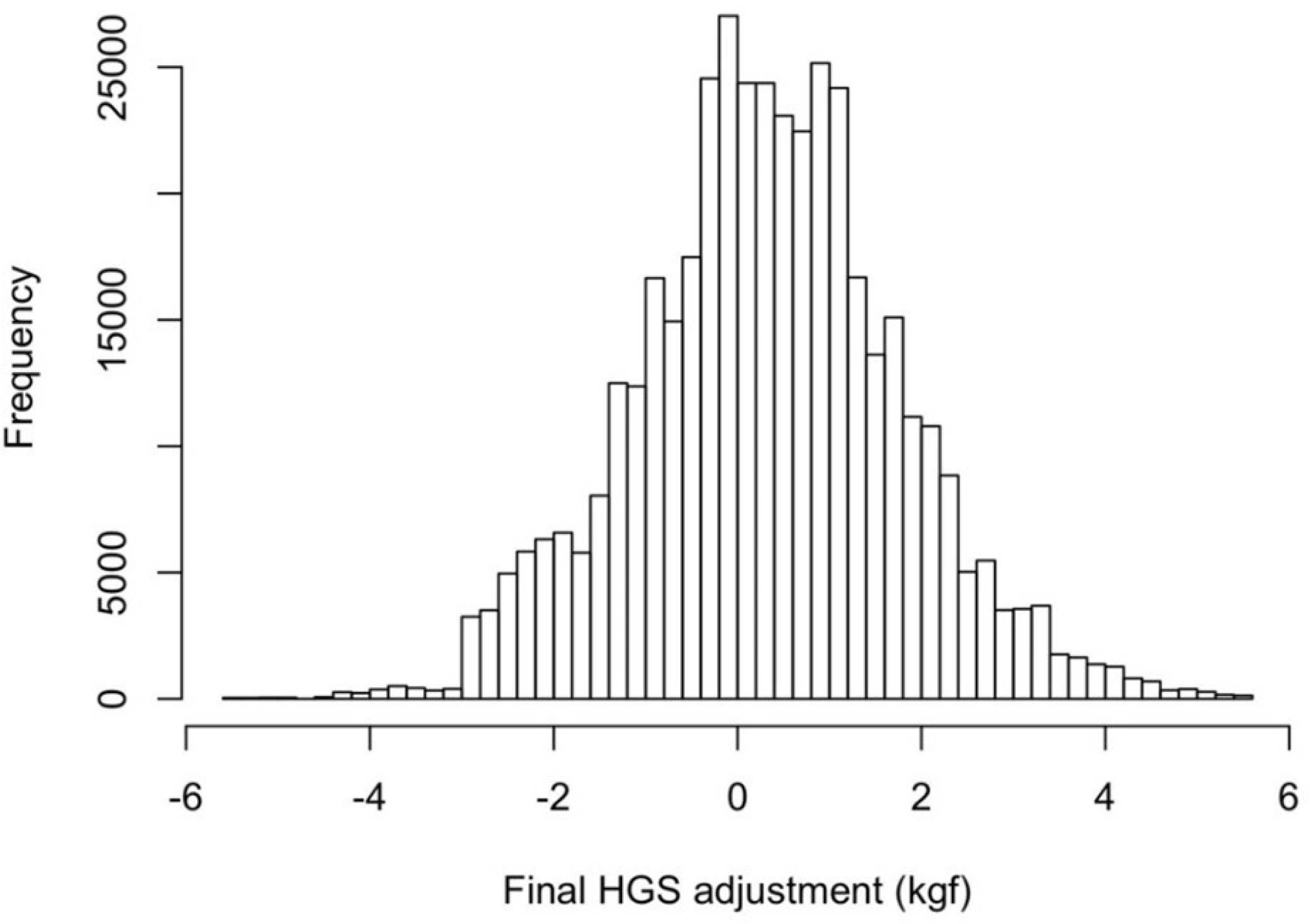
Distribution of hand grip strength adjustments. Histogram of adjustment values for the average hand grip strength (HGS) measures of 422,364 UKB participants at baseline assessment.

**Figure SD4.IV.**
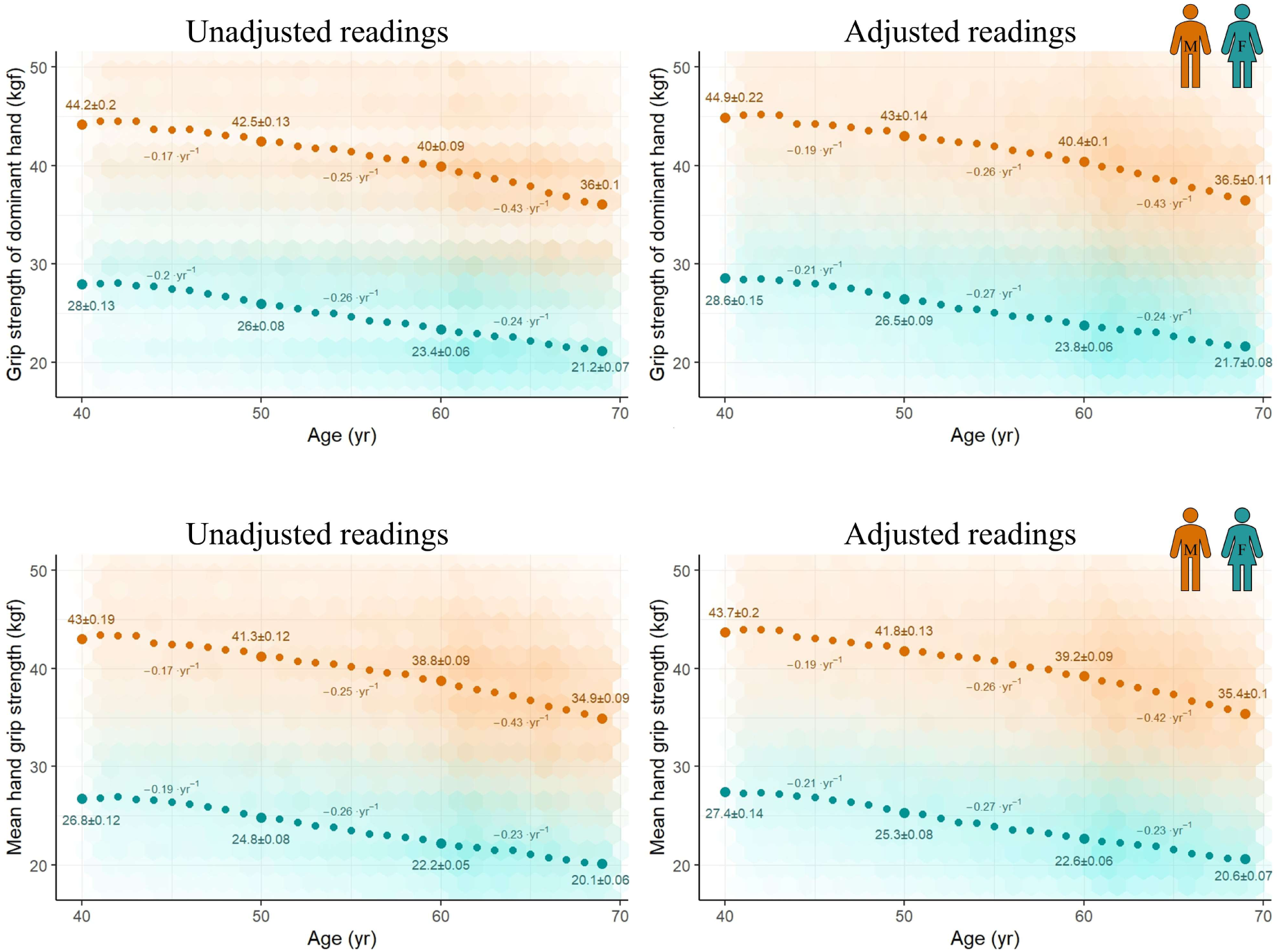
Age-related trajectories are similar for unadjusted and adjusted hand grip strength readings (left and right hand measures). Sex-specific relationships between grip strength of the left (top row) and right (bottom row) hand and age in the Cross-sectional Participants. Teal and orange represent females and males, respectively. Large points and values (mean ± SEM) are observed values at 40, 50, 60, and 69 yr, and inferred annual change over the corresponding 9–10 year period. Observations are plotted using hexagonal bin plotting, with greater opacity indicating higher density of participant measures. *y*-axes are truncated to visualise better the general trends in the data.

**Figure SD4.V.**
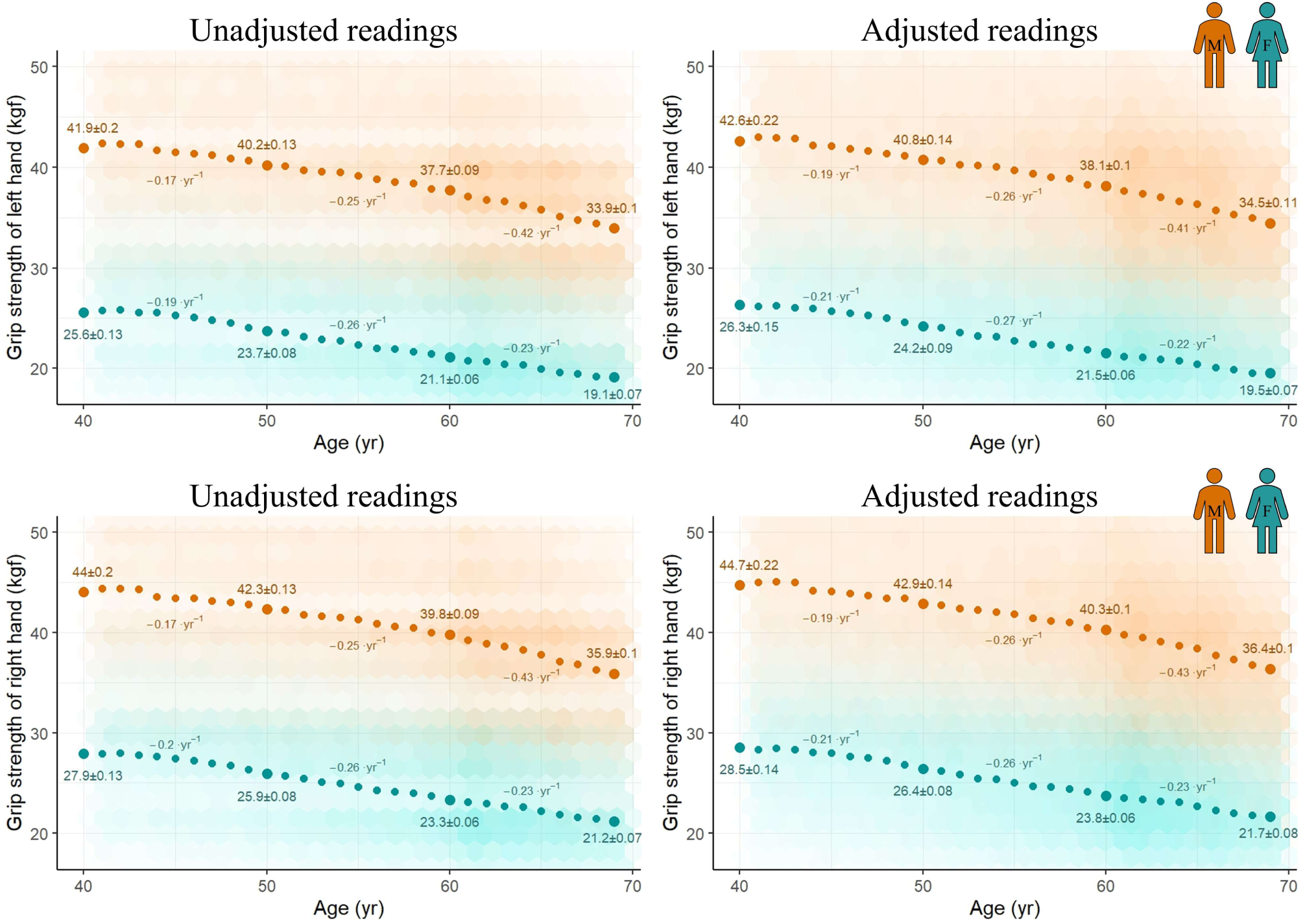
Age-related trajectories are similar for unadjusted and adjusted hand grip strength readings (dominant hand and mean measures). Sex-specific relationships between grip strength of the dominant hand (top row) and mean hand grip strength (HGS) (bottom row) and age in the Cross-sectional Participants. Dominant hand was determined by participants’ responses to the question “Are you right or left handed?” (Data-Field 1707); participants who responded “Use both right and left hands equally” or “Prefer not to answer” were excluded from the dominant hand plots. Teal and orange represent females and males, respectively. Large points and values (mean ± SEM) are observed values at 40, 50, 60, and 69 yr, and inferred annual change over the corresponding 9–10 year period. Observations are plotted using hexagonal bin plotting, with greater opacity indicating higher density of participant measures. *y*-axes are truncated to visualise better the general trends in the data.

### Supplementary Data 5 Muscle mass adjustment and estimation

#### Adjustment of BIA impedance values

A preliminary analysis of 45,686 participants who attended a repeat assessment revealed an apparent MM increase in an unexpectedly large proportion (∼37.3%) of the 44,702 participants for whom two Tanita MM estimates were available. This phenomenon was particularly pronounced in females (∼45.2%), especially those who were younger at baseline assessment (Fig. SD5.I). Because a widespread trend of age-related MM increase seemed biologically implausible—and also contradicted our cross-sectional finding of age-related MM decline—we decided to investigate further.

We first stratified the 44,702 participants by the UKB Assessment Centre at which their repeat assessment was performed. Age-related MM decline was observed on average for males at all three Assessment Centres. Female age-related MM decline was also observed on average in females assessed at the Reading (N = 6,288) and Newcastle (N = 11,292) centres, whereas females who attended the Cheadle (N = 27,122) centre tended to exhibit age-related MM increase on average (Fig. SD5.IIa). Two of the seven BIA impedance devices used at repeat assessments were based at Reading (#001480 and #003169), two at Newcastle (#003167 and #003168), and three at Cheadle (#001477, #001486, and #003173). None of these devices was transferred between Assessment Centres during the assessment period. Females assessed at Cheadle were reported to have—on average—greater arm MM at repeat than at baseline assessment, regardless of the BIA impedance device used at repeat assessment; in contrast, all four of the devices based at Reading and Newcastle were associated with age-related female MM decline (Fig. SD5.IIb). Male age-related MM decline held on average across all seven BIA impedance devices, but was markedly less pronounced in participants assessed at Cheadle (Fig. SD5.IIb). The MM estimates accorded with the lower arm impedance values (Ω) generally observed in the Cheadle data (Fig. SD5.IIc).

**Figure SD5.I.**
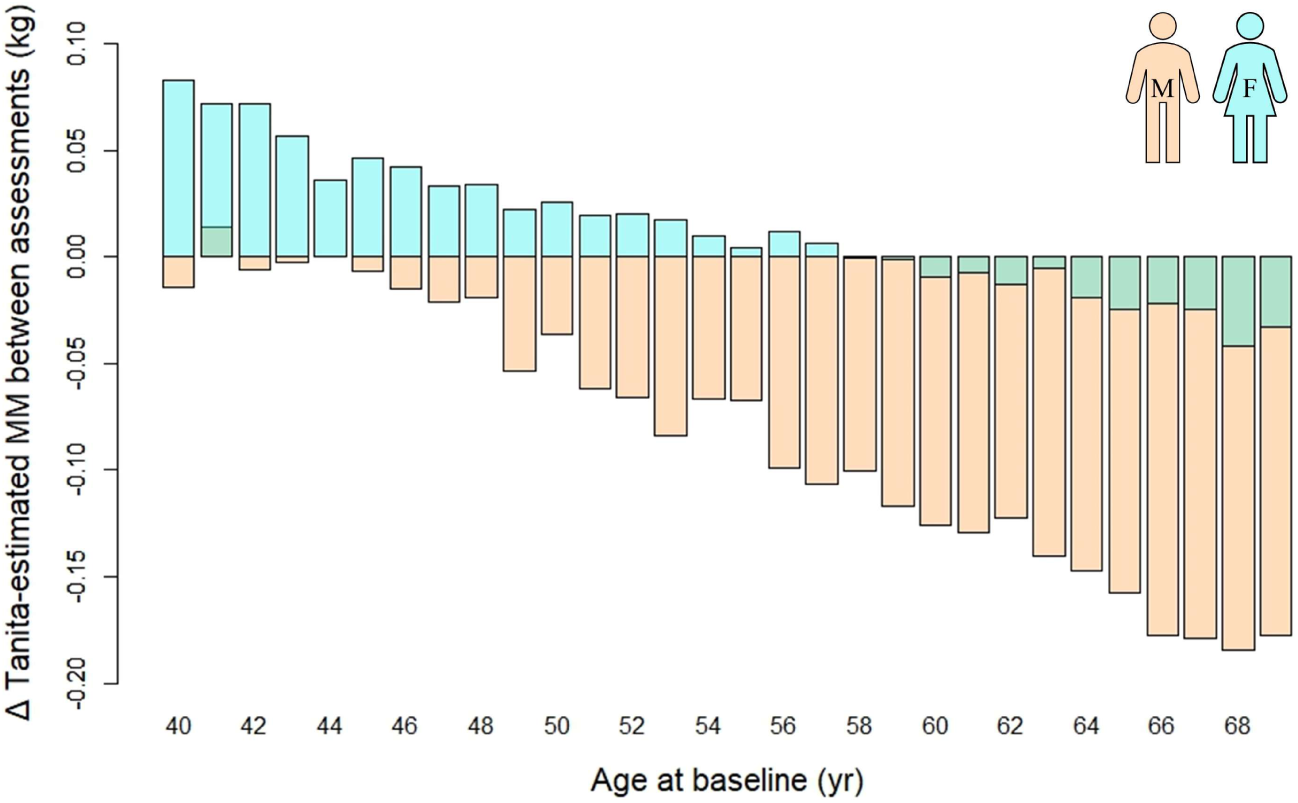
Apparent age-related arm muscle mass increase in over 1/3^rd^ of participants. N = 44,702. Bars indicate average change in mean arm muscle mass (MM) between baseline and repeat assessment for participants grouped by whole year of age at baseline assessment. Teal and orange represent females (N = 22,967) and males (21,735), respectively, with green indicating areas of overlap. Average interval between assessments was 8.92 ± 1.76 (SD) yr. MM estimates were generated by a Tanita Body Composition Analyser BC-418MA.

**Figure SD5.II.**
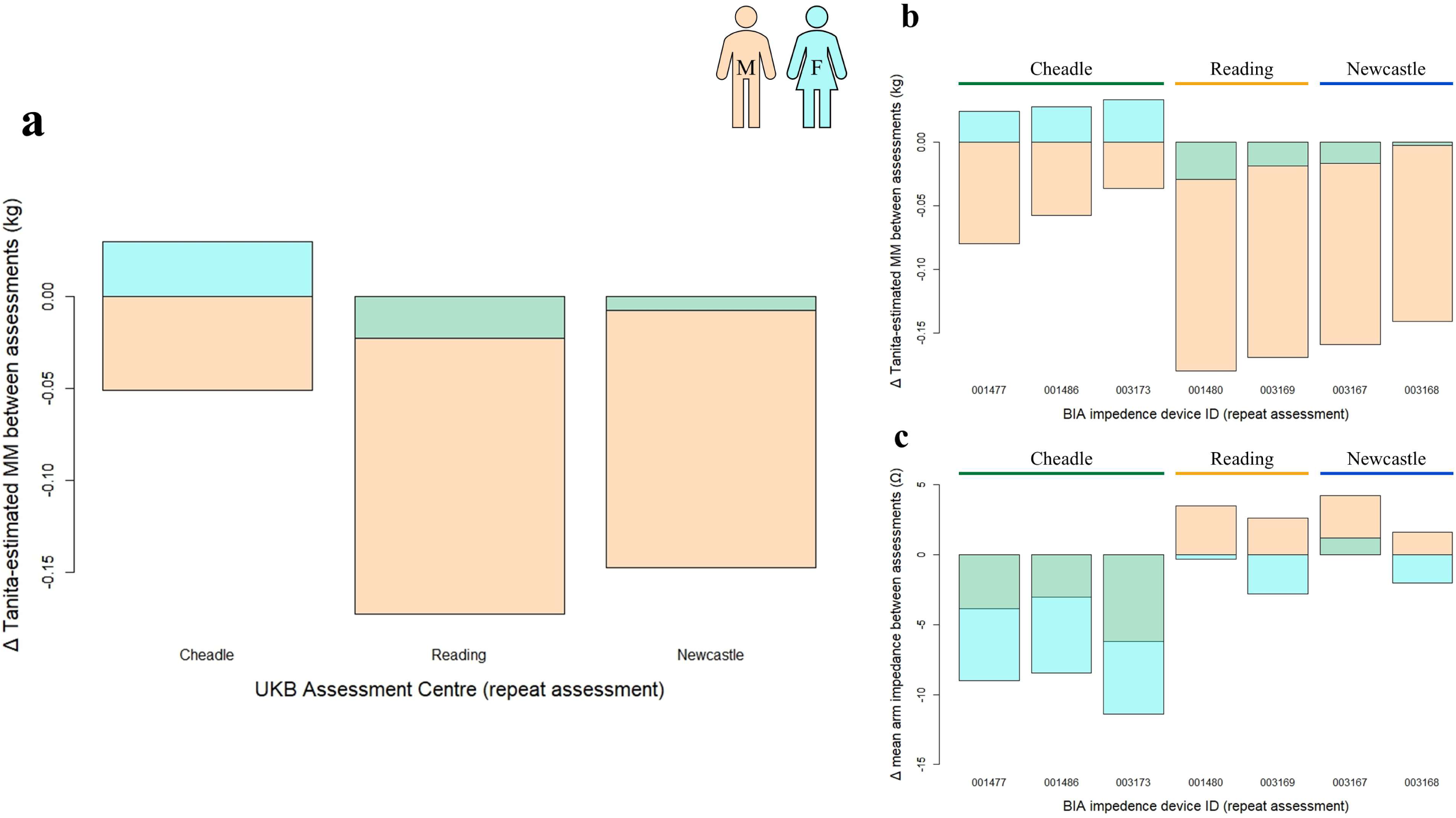
Tanita bioelectrical impedance analysis devices based in Cheadle give aberrant readings. 44,702 participants assessed longitudinally, with an average of 8.92 ± 1.76 (SD) yr between baseline and repeat assessment. Teal and orange represent females (N = 22,967) and males (21,735), respectively, with green indicating areas of overlap. **a,b** Females whose repeat assessment was conducted in Cheadle were reported to have—on average—greater mean arm muscle mass (MM) at repeat than at baseline assessment, regardless of the bioelectrical impedance analysis (BIA) impedance device used at repeat assessment; in contrast, average female MM was lower at repeat assessment across all four BIA impedance devices based in Reading and Newcastle. Reported loss of MM at repeat assessment in males was also markedly less pronounced in Cheadle. **c** Arm impedance data accord with the MM estimates, as lower impedance values are associated with higher estimated MM, due to the relatively low impedance of muscle tissue compared with fat and bone.

We next checked whether differences between Cheadle and the other two Assessment Centres remained after controlling for potential demographic differences between them. The *matchit* function of the ‘MatchIt’ R package (referenced in main manuscript) was used to match Cheadle participants with participants of the same sex who were assessed at either Reading *or* Newcastle (‘Other’), based on similarity across predefined criteria, namely age at baseline assessment; interval between baseline and repeat assessment (‘Interval’); standing height at baseline assessment; and body mass at baseline assessment. Nearest neighbour matching without replacement was performed, with Mahalanobis distances used as the measure of similarity. The *caliper* argument was used to prevent matching of participants whose values for a given criterion differed by more than a predefined amount (age at baseline assessment: ≤0.5 yr; Interval: ≤0.45 yr; standing height at baseline assessment: ≤1 cm; body mass at baseline assessment: ≤1 kg). This matching procedure returned 4,722 same-sex Cheadle:Other participant pairs (N = 9,444) that were well-matched across all matching criteria (all Standard Mean Differences [SMDs] ≥−0.0017 and ≤0.0267; all Variance Ratios [VRs] ≥0.9852 and ≤1.0008; see Griefer [2023]—referenced in main manuscript). Nevertheless, a clear difference was still apparent between Cheadle and the other two Assessment Centres (Fig. SD5.III), confirming that the discrepancy was not the result of demographic differences in the matching criteria between Assessment Centres. We thus concluded that BIA measurement error in Cheadle was likely the cause of the discrepancy, and that adjustment of the Cheadle BIA impedance values was therefore warranted. As no information was available on BIA impedance device performance, accuracy, or calibration history, it was necessary for us to develop our own procedure for adjusting impedance values from the BIA impedance devices based at Cheadle.

**Figure SD5.III.**
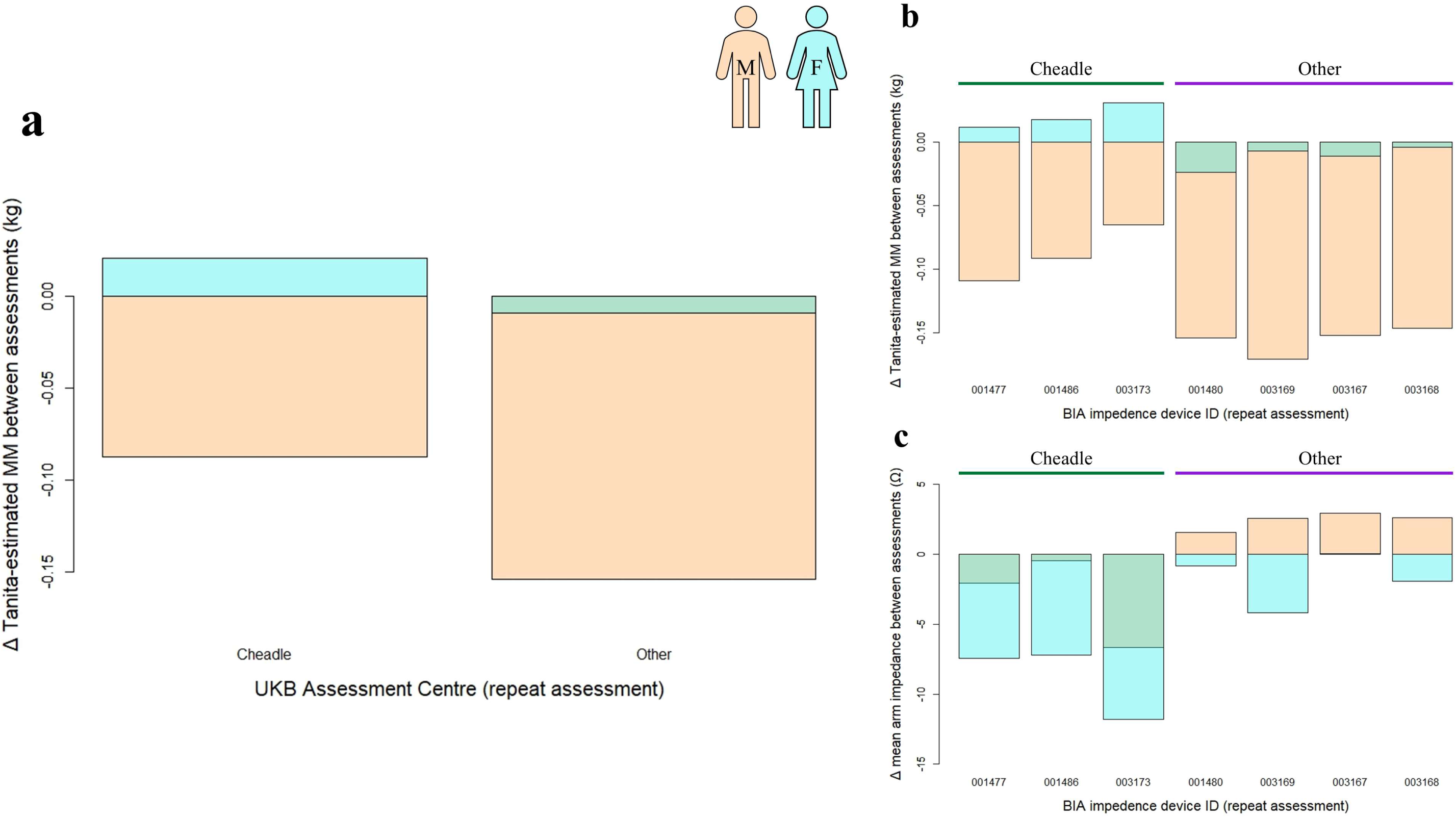
Discrepancies between UK Biobank Assessment Centres persist after controlling for demographic differences. N = 9,444, comprising 4,722 same-sex Cheadle:Other participant pairs matched on similarity in age and body size at baseline assessment, as well as interval (yr) between baseline and repeat assessment. ‘Other’ denotes participants whose repeat assessment was conducted at either Reading *or* Newcastle. Teal and orange represent females (N = 5,110; 2,555 pairs) and males (N = 4,334; 2,167 pairs), respectively, with green indicating areas of overlap. The figure panels are analogous to those in Fig. SD5.IIa–c, and so demonstrate that the aberrant Cheadle data are not explicable by demographic differences in the matching criteria between Cheadle and the other two Assessment Centres.

We first formally assessed whether the data supported the necessity of adjustments at the level of BIA impedance device and/or Assessment Centre. To do so, we used the *lm* base R function to fit multiple linear regression models to various subsets of the repeat assessment data, with mean arm impedance (Ω) as the dependent variable, and age and sex as covariates. We then tested whether including BIA impedance device *or* Assessment Centre as an additional covariate in these models improved their performance relative to their respective, simpler (nested) models which did not include them. Likelihood-ratio tests— as performed using the *lrtest* function of the ‘lmtest’ R package (version 0.9-40: Hothorn et al. 2015)—were used for comparative assessment of model performance. Inclusion of Assessment Centre as a covariate significantly improved model performance in the full participant dataset (*P* <0.001; FPR <0.001), but not in a model fitted to Reading and Newcastle data only (*P* = 0.58; FPR = 0.5). Furthermore, inclusion of BIA impedance device as a covariate did not improve performance in this model (*P* = 0.02; FPR = 0.18). We therefore concluded that the four BIA impedance devices based at Reading and Newcastle behaved in an essentially similar manner. We found that including BIA impedance device as a covariate significantly improved performance in a model fitted to Cheadle data only (*P* <0.001; FPR <0.001), but not when readings from BIA impedance device #003173 were omitted (i.e. with only readings from devices #001477 and #001486 included; *P* = 0.01; FPR = 0.11). We therefore deemed it appropriate to make two separate adjustments to the Cheadle impedance values: one to impedances values from device #003173, and a second to impedance values from devices #001477 and #001486.

Calculation of MM (see ‘Generating novel muscle mass estimates’ below), required several impedance measures, namely: whole-body impedance, ‘arms impedance index’ (i.e. height^2^ [in metres^2^] divided by the sum of the left and right arm impedance values), and ‘legs impedance index’ (i.e. height^2^ [in metres^2^] divided by the sum of the left and right leg impedance values). It was therefore necessary to calculate separate adjusted values of summed arm impedance, summed leg impedance, and whole-body impedance. For each of these impedance measures, we sought to calculate two simple scalar adjustment values for the correction of values ascertained at Cheadle (i.e. independently of age, sex, ethnicity, or other covariates): one for participants assessed using impedance device #003173, and one for participants assessed using either #001477 or #001486.

To calculate adjustment values for the summed arm impedance data, we again performed participant matching on the initial pool of 45,686 participants, in the manner described above. ‘Cheadle’ and ‘Other’ participants of the same sex were matched by age at baseline assessment (≤ 0.5 yr), Interval (≤ 0.45 yr), summed arm impedance at baseline assessment (≤ 2 Ω), and Townsend Deprivation Score (a measure of material deprivation: min. −6.26; max. 10.1; mean −1.89) at recruitment (≤ 1). The matching procedure returned 3,746 Cheadle:Other participant pairs. Our approach implicitly assumed that any within-pair discrepancy in summed arm impedance at repeat assessment was the result of measurement error in Cheadle. Therefore, the summed arm impedance values of each pair participant at repeat assessment were essentially treated as ‘observed’ (Cheadle participant) and ‘expected’ (Other participant) values. The average difference between these observed and expected values was then calculated across participant pairs, with the two mean differences—i.e. one mean for the Cheadle participants assessed on impedance device #003173, and a second for those assessed on either #001477 or #001486—subsequently used as the final adjustment values. A comparable procedure was performed to calculate adjustment values for the summed leg impedance data (2,385 Cheadle:Other pairs), but with summed leg impedance (≤ 2 Ω) replacing summed arm impedance, and with number of days per week of vigorous physical activity (10+ minutes) (‘vigorous physical activity’) at baseline assessment (≤ 1 day) included as an additional matching parameter. This procedure was then performed for a third and final time to calculate adjustment values for whole-body impedance data (2,883 Cheadle:Other pairs), with whole-body impedance at baseline assessment (≤ 1 Ω) replacing summed arm/leg impedance as the impedance-related matching parameter, and without the inclusion of vigorous physical activity at baseline assessment as a matching parameter. Note that a new matched subset was generated in each case (e.g. the ‘leg’ subset was not generated from the ‘arm’ subset), and that these subsets were used only to calculate adjustment values (i.e. none of these three matched subsets was included in any subsequent analyses).

All three subsets were well-matched across their respective matching criteria (all SMDs ≥−0.0058 and ≤0.0353; all VRs ≥0.9514 and ≤1.0045). Significance tests (Cheadle vs Other)—on age at baseline assessment, Interval, number of days per week of moderate physical activity (10+ minutes) (‘moderate physical activity’) at baseline assessment, vigorous physical activity at baseline assessment, Townsend Deprivation Score at recruitment, and the relevant impedance value at baseline assessment—were all unequivocally non-significant in the three matched subsets (all *P*-values and FPRs >0.09), with the exception of moderate physical activity at baseline assessment in the ‘legs’ subset (*P* = 0.01; FPR = 0.12). Underlying demographic differences between the Cheadle and Other participant groups were therefore considered sufficiently minor to justify treating the impedance values of Other participants as the ‘expected’ impedance values of their Cheadle counterparts. The final adjustment values—as subsequently applied to the respective impedance values—are given in Table SD5.I.

**Table SD5.I.**
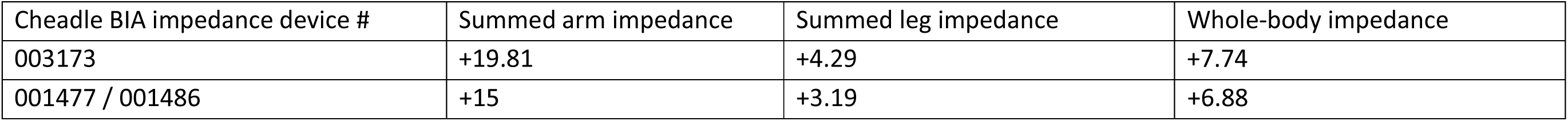
Adjustment values (Ω) for impedance values from the three bioelectrical impedance analysis (BIA) impedance devices based in Cheadle.

| Cheadle BIA impedance device # | Summed arm impedance | Summed leg impedance | Whole-body impedance |
| --- | --- | --- | --- |
| 003173 | +19.81 | +4.29 | +7.74 |
| 001477 / 001486 | +15 | +3.19 | +6.88 |

Once adjusted, the Cheadle impedance data proved to be broadly similar to the Reading and Newcastle impedance data (Fig. SD5.IV), which we interpreted as evidence of a successful and appropriate adjustment. These adjusted impedance data were then used in the generation of novel estimates of mean arm muscle mass.

**Figure SD5.IV.**
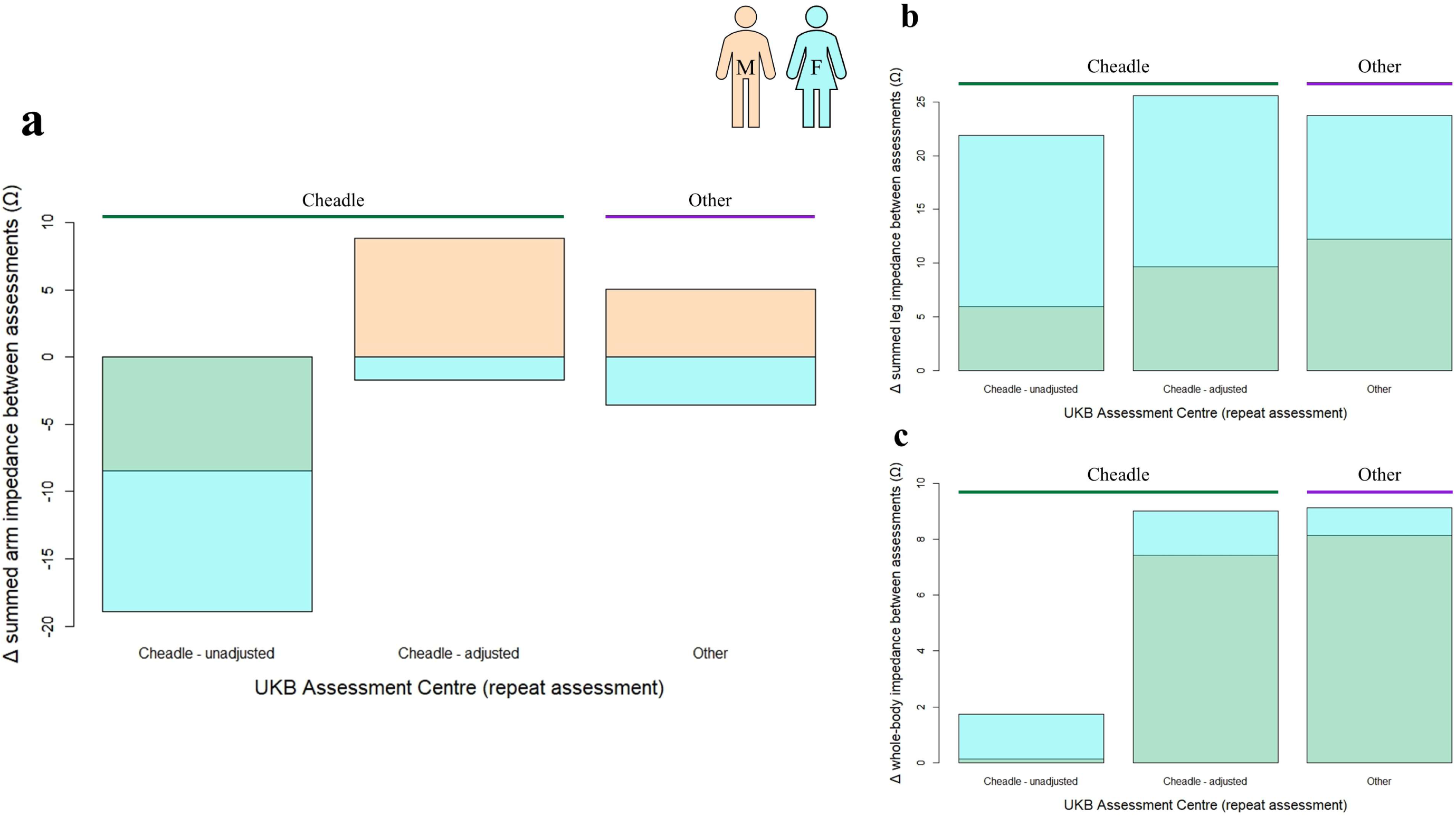
Adjustment of bioelectrical impedance device impedance values minimises differences between UK Biobank Assessment Centres. N = 9,444. These participants are the same as those shown in Fig. SD5.III, i.e. 4,722 same-sex Cheadle:Other participant pairs matched on similarity in age and body size at baseline assessment, as well as interval (yr) between baseline and repeat assessment. ‘Other’ denotes participants whose repeat assessment was conducted at either Reading *or* Newcastle. Teal and orange represent females (N = 5,110; 2,555 pairs) and males (N = 4,334; 2,167 pairs), respectively, with green indicating areas of overlap. The adjusted Cheadle bioelectrical impedance analysis (BIA) impedance values accord more closely with the Other values than do the unadjusted Cheadle BIA impedance values, particularly in the case of summed arm (**a**) and whole-body (**c**) impedance. The difference in summed leg impedance values between Cheadle and the other two Assessment Centres was much more modest (**b**).

#### Generating novel muscle mass estimates

We generated novel MM estimates using sex-specific equations based on those of Powell et al. (2020) (‘Powell’ equations; see main manuscript). Sex-specific equations produced substantially better MM estimates in preliminary analysis than did a single equation with sex included as a covariate (data not shown). Derivation and validation of equations was performed in a subset of 4,182 participants who were subjected to both a BIA reading and a DXA scan during their repeat assessment in Cheadle, and for whom all other necessary impedance and anthropometric measures were available. DXA-derived MM estimates were used as reference values against which to develop the predictive equations.

To develop a female-specific equation, we first split the female participants (N = 2,179) 1:1 into Derivation and Validation groups. The following linear model was then fitted to the Derivation group using the *lm* base R function:

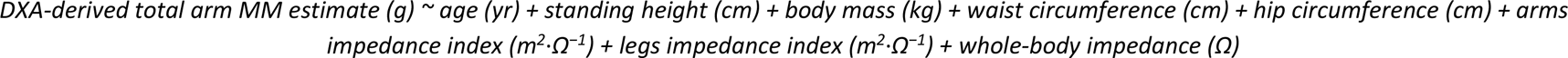

where ‘arms impedance index’ = (height^2^ [in metres^2^] / summed arm impedance [Ω]), and ‘legs impedance index’ = (height^2^ [in metres^2^] / summed leg impedance [Ω]). The *ols_step_best_subset* function of the ‘olsrr’ R package (version 0.5.3: Hebbali 2020) was then used to determine which set of independent variables to retain in the final model, with AIC values as the objective function for selection of model terms. The *ols_step_best_subset* function works by fitting all possible models given the independent variables supplied, and returning the optimal model (in this case, the model with the lowest AIC value) for each number of independent variables (i.e. for models fitted using 1, 2 … *k* independent variables, where *k* = no. independent variables). This final model was then used to estimate total arm MM (g) in the Validation group via the *predict* function in the R ‘stats’ base package. Finally, both DXA-derived total arm MM (g) estimates and our own total arm MM (g) estimates were divided by 2,000, to convert values to mean arm MM (kg).

This modelling procedure was performed twice in the females: once using unadjusted Ω values, and once using adjusted Ω values (see previous section ‘Adjustment of BIA impedance values’). Analogous analyses were also performed for males (N = 2,003), resulting in a total of four models, the coefficients of which are given in the Table SD5.II.

**Table SD5.II.** Coefficients in final models. Covariates that were dropped from final models are indicated by a coefficient of zero. The coefficients of the Powell equations are also provided for reference.

| | Females<br>(unadjusted $\Omega$<br>values) | Females<br>(adjusted $\Omega$<br>values) | Reference model:<br>females<br>(Powell) | Males<br>(unadjusted $\Omega$<br>values) | Males<br>(adjusted $\Omega$<br>values) | Reference<br>model: males<br>(Powell) |
| --- | --- | --- | --- | --- | --- | --- |
| Intercept | 614.1 | 355.7 | 2,479 | 2,012 | 2,491 | 6,738 |
| Age (yr) | -4.425 | -2.374 | -7.9 | -14.19 | -17.95 | -15.47 |
| Standing height (cm) | 15.31 | 15.21 | 0 | 21.26 | 0 | 0 |
| Body mass (kg) | 22.51 | 22.28 | 34.5 | 20.97 | 18.56 | 62.5 |
| Waist circumference<br>(cm) | 4.353 | 4.25 | 2.008 | -12.63 | -9.963 | -18.93 |
| Hip circumference<br>(cm) | -13.6 | -13.28 | -19.87 | 10.04 | 15.15 | -31.58 |
| Arms impedance<br>index ($\text{m}^2 \cdot \Omega^{-1}$) | 478,800 | 525,600 | 724,356 | 747,600 | 969,500 | 929,870 |
| Legs impedance<br>index ($\text{m}^2 \cdot \Omega^{-1}$) | 0 | 0 | -118,528 | -222,000 | -135,300 | -388,873 |
| Whole-body<br>impedance ($\Omega$) | -1.573 | -1.538 | -1.196 | -4.332 | -1.876 | -3.742 |

#### Comparative analysis of muscle mass estimates

To determine which set of BIA-derived MM estimates was most similar to the DXA MM estimates, we calculated mean prediction error, maximum prediction error, and root mean square error (RMSE—calculated using the *rsme* function of the ‘Metrics’ R package [version 0.1.4: Hamner et al. 2022]) for the Tanita estimates, the Powell estimates (with unadjusted and adjusted Ω values), and our novel estimates (with unadjusted and adjusted Ω values in the corresponding derived models) when compared with the Validation group (Table SD5.III). We also performed pairwise model comparisons of predictive accuracy with Diebold-Mariano tests using the *dm.test* function of the ‘forecast’ package (version 8.21: Hyndman et al. 2020). Our novel MM estimates on the Validation group proved to be significantly more similar to the DXA MM measurements than did the Powell and Tanita MM estimates, regardless of whether unadjusted or adjusted impedance values were used for model derivation; however, no difference in predictive accuracy was observed between these two models (Tables SD5.III and IV). Nevertheless, we opted to use the MM estimates generated using adjusted impedance values for our primary analyses, due to the superior mean prediction error, maximum prediction error, and RMSE values, along with the strong prior evidence that adjustment of the Cheadle BIA impedance data was warranted (see ‘Adjustment of BIA impedance values’). Supplementary Fig. 3 shows the baseline assessment data for a) the Tanita estimates, b) the Powell estimates (note that there is no distinction between the ‘unadjusted’ and ‘adjusted’ Powell estimates at baseline assessment, because i) impedance values from baseline assessments were not adjusted, and ii) the Powell coefficients were not derived using the repeat assessment data), and c) novel estimates generated using unadjusted impedance values. Each method showed clear sexual dimorphism, with greater muscle mass decline with age in males than females, in both absolute and percentage terms.

**Table SD5.III.** Analysis of similarity of dual x-ray absorptiometry-derived mean arm muscle mass estimates to five other sets of mean arm muscle mass estimates. ‘Error’ was defined as a participant’s dual x-ray absorptiometry (DXA) mean arm muscle mass (MM) estimate—implicitly treated as the ‘true’ MM value—minus their corresponding bioelectrical impedance analysis (BIA)-derived MM estimate.

|  | Mean prediction error (kg) | Max. prediction error (+ve or -ve) (kg) | Root Mean Square Error (RMSE) (kg) |
| --- | --- | --- | --- |
| Tanita estimates | -0.031 | 1.569 | 0.236 |
| Powell estimates (unadjusted $\Omega$ values) | 0.118 | 1.598 | 0.259 |
| Powell estimates (adjusted $\Omega$ values) | 0.18 | 1.435 | 0.291 |
| Novel estimates (unadjusted $\Omega$ values) | -0.007 | 1.35 | 0.244 |
| Novel estimates (adjusted $\Omega$ values) | 0.003 | 1.2 | 0.211 |

**Table SD5.IV.** Results of Diebold-Mariano tests of predictive accuracy. Test results are reported as ‘*P*-value (FPR score)’.

| | Tanita estimates | Powell estimates<br>(unadjusted $\Omega$<br>values) | Powell estimates<br>(adjusted $\Omega$ values) | Novel estimates<br>(unadjusted $\Omega$<br>values) | Novel estimates<br>(adjusted $\Omega$ values) |
| --- | --- | --- | --- | --- | --- |
| Tanita estimates | - | - | - | - | - |
| Powell estimates<br>(unadjusted $\Omega$ values) | < 0.001<br>(< 0.001) | - | - | - | - |
| Powell estimates<br>(adjusted $\Omega$ values) | < 0.001<br>(< 0.001) | < 0.001<br>(< 0.001) | - | - | - |
| Novel estimates<br>(unadjusted $\Omega$ values) | < 0.001<br>(< 0.001) | < 0.001<br>(< 0.001) | < 0.001<br>(< 0.001) | - | - |
| Novel estimates<br>(adjusted $\Omega$ values) | < 0.001<br>(< 0.001) | < 0.001<br>(< 0.001) | < 0.001<br>(< 0.001) | 0.73<br>(0.50) | - |

### Supplementary Data 6 Muscle quality

#### Properties of the muscle quality measures MQ and MQ’

**Figure SD6.I.**
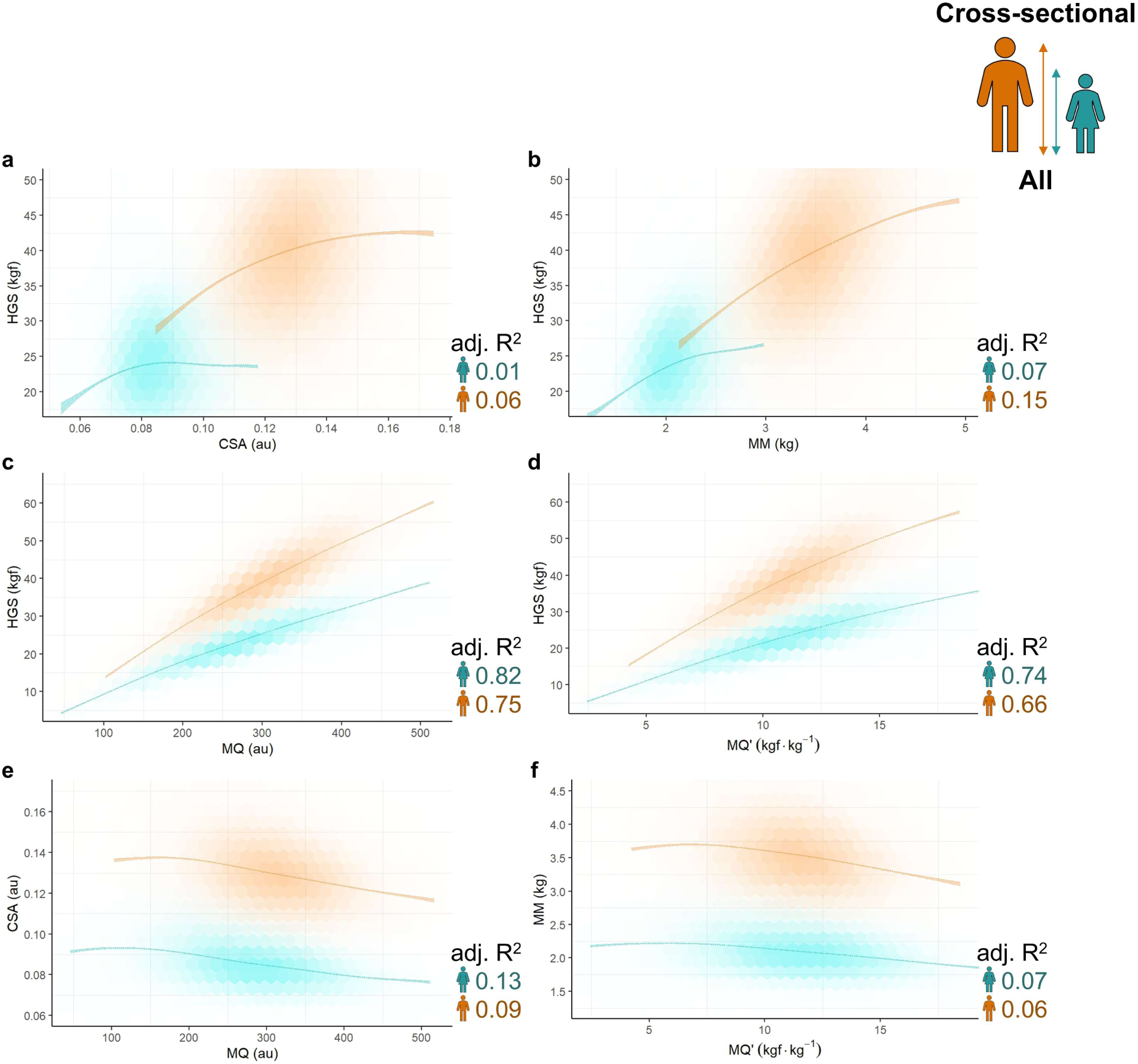
Properties of the muscle quality measures MQ and MQ’. Data are from baseline assessments of Cross-sectional Participants (females, teal; males, orange). Left column: apsects of MQ, calculated as *(mean hand grip strength) /(mean arm muscle cross-sectional area)*. Right column: aspects of MQ’, calculated as *(mean hand grip strength)/(mean arm muscle mass*). The observed data are plotted using hexagonal bin plotting, with greater opacity indicating higher density of participant measures. Coloured bands represent the 95% confidence intervals for the sex-specific generalised additive mixed model (GAMM)-fitted relationships *y ∼ s(x)*, where *s(x)* denotes a spline fitted to the independent variable (see Methods and Supplementary Data 2 for more details on GAMM-fitting procedures). **a–b** Fitted relationships between mean hand grip strength (HGS) and two measures of muscle size—mean arm muscle cross-sectional area (CSA—Methods) (a) and mean arm muscle mass (MM) (b)—demonstrate that HGS is only weakly correlated with muscle size. As age is not accounted for in the GAMMs, the especially weak correlation between HGS and muscle size in females accords with our finding that female MM changes little with age, whereas HGS decreases markedly. **c–f** MQ and MQ’ measures are more reflective of HGS (c–d) than of muscle size (e– f). An inverse correlation between muscle quality and size is apparent in both sexes, whether size is defined as (CSA) (e) or MM (f), confirming that the correlation is not an artefact of our ulna length calcuations (Supplementary Discussion). Because arm muscles tend to decrease in both quality and size with age (Results), controlling for age would be expected to strengthen— rather than diminish—the inverse correlation.

#### Ulna length and muscle quality

##### 1. Calculating ulna length using predicted maximum standing height minimises artefactual sexual dimorphism in age-related decline in muscle quality

The example below illustrates how estimates of MQ are affected by the use of observed standing height *versus* predicted maximum standing height (PMSH) in MQ calculations.

In this example, MQ is estimated at both 40 and 69 yr—i.e. at both ends of the Cross-sectional Participants age range—using the mean female Cross-sectional Participant values for HGS (23.65 kgf) and MM (2.1 kg). These values are then used to estimate MQ using i) observed standing height and ii) PMSH. Even in the absence of true MQ decline (i.e. no change of HGS, UL, and MM), general age-related decline in standing height will affect MQ—as estimated in this study—due to the lower *estimated* UL of older, shorter participants (Methods). In this example, a 69-year-old female of mean observed standing height for her age group (160.43 cm) is predicted to have a MQ value 6.7% lower (−18.98 au) than that of a 40-year-old female of mean observed standing height for her age group (164.5 cm) (283.23 vs 264.25 au), with this difference wholly due to the shorter estimated ulna length (UL) of the older female (25.15 vs 23.46 cm— 1.69 cm). If the equivalent analysis is performed using PMSH rather than observed standing height, the difference in predicted UL is attenuated (25.15 vs 24.16 cm—0.99 cm), and the difference in MQ reduces to −3.91% (−11.08 au). While a birth cohort effect (i.e. a trend towards younger individuals attaining greater maximum standing heights) probably contributes to the lower average observed standing height of older UKB participants, this example nevertheless demonstrates how an artefactual age-related decline in MQ may arise simply from participants experiencing a normal age-related reduction in standing height. Calculating UL using PMSH rather than observed standing height in this study was thus warranted to minimise the impact of this phenomenon.

**Mean values for female Cross-sectional Participants**

HGS (kgf) = 23.65 MM (kg) = 2.1

Observed standing height (cm) at 40 yr = 164.5

Observed standing height (cm) at 69 yr = 160.43

PMSH (cm) at 40 yr = 164.53

PMSH (cm) at 69 yr = 162.37

UL (cm) at 40 yr, calculated using observed standing height = 25.15

UL (cm) at 69 yr, calculated using observed standing height = 23.46

UL (cm) at 40 yr, calculated using PMSH = 25.15

UL (cm) at 69 yr, calculated using PMSH = 24.16

**MQ calculations**

CSA = MM / UL

MQ = HGS / CSA

**MQ (observed standing height)**

<u>At 40 yr</u>

CSA = 2.1 / 25.15 = 0.0835

MQ = 23.65 / 0.0835 = 283.23 au

<u>At 69 yr</u>

CSA = 2.1 / 23.46 = 0.0895

MQ = 23.65 / 0.0895 = 264.25 au

<u>Difference</u>

−18.98 au (−6.7%)

**MQ (PMSH)**

<u>At 40 yr</u>

CSA = 2.1 / 25.15 = 0.0835

MQ = 23.65 / 0.0835 = 283.23 au

<u>At 69 yr</u>

CSA = 2.1 / 24.16 = 0.0869

MQ = 23.65 / 0.0869 = 272.15 au

<u>Difference</u>

−11.08 au (−3.91%)

#### 2. Muscle quality decline in greater in females irrespective of how ulna length is estimated

**Figure SD6.II.**
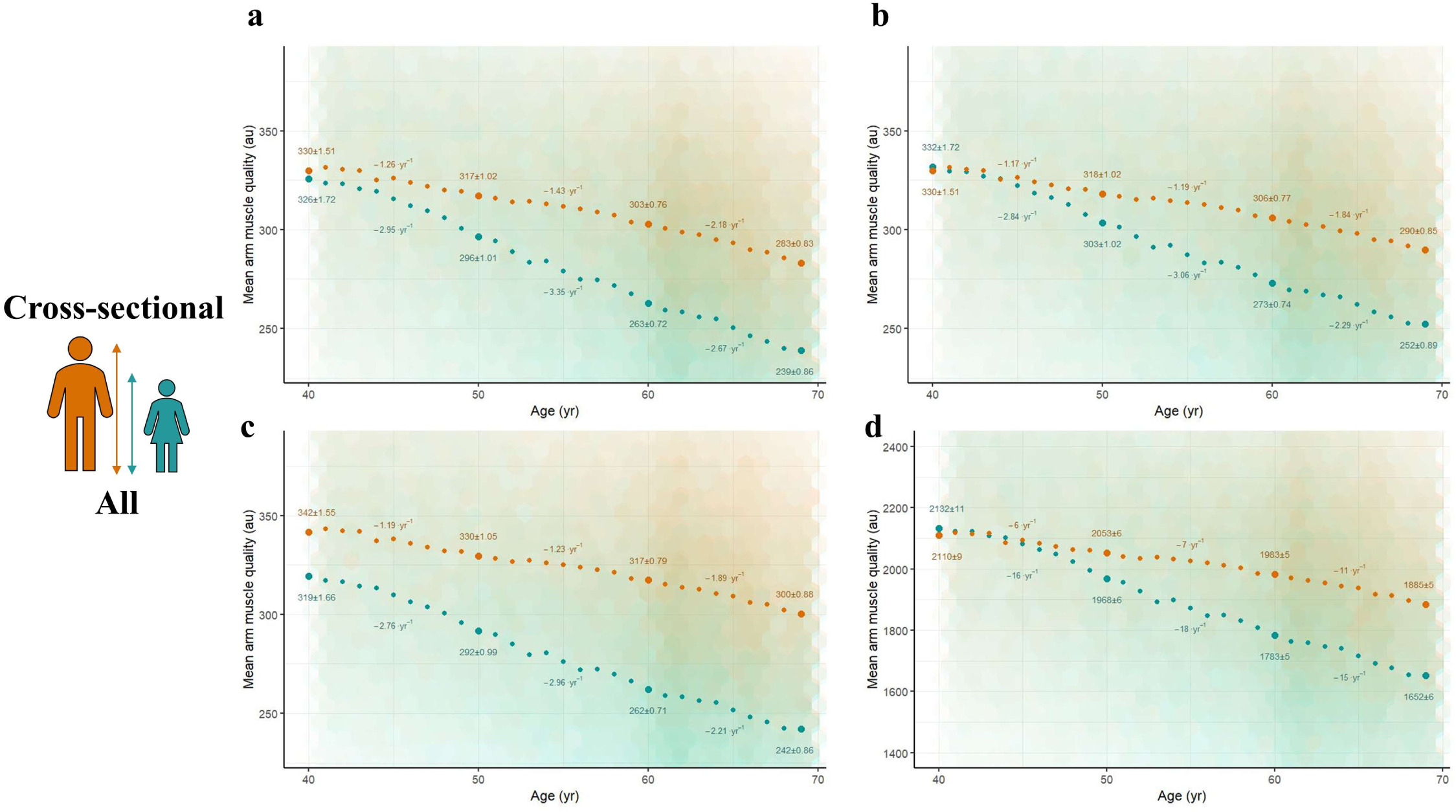
Muscle quality decline is greater in females irrespective of how ulna length is estimated. Sex-specific relationships between muscle quality and age are shown for the Cross-sectional Participants (females, teal; males, orange). Large points and values (mean ± SEM) are observed values at 40, 50, 60, and 69 yr, and inferred annual change over the intervening period. The observed data are plotted using hexagonal bin plotting, with greater opacity indicating higher density of participant measures. *y*-axes are truncated to visualise better the general trends in the data. **a** Muscle quality is calculated using observed standing height (cm) rather than predicted maximum standing height (PMSH—Methods; also see ‘1) Calculating ulna length using observed standing height introduces an artefactual age-related decline in muscle quality’). **b** Female ulna length (UL) is estimated using a re-arranged version of the female equation given in Table 5 (Equation F) of Madden et al. (2020) (referenced in main manuscript)—i.e. *UL = (PMSH − 84.67)/3.14*—as opposed to the Elia (2003) (referenced in main manuscript) equation used in the primary analyses (Methods). Male muscle quality estimates are unchanged. **c** The Madden et al. equations for Asian, black, and white individuals have ethnicity-specific slopes (Asian = 3.26; black/white = 3.14), and sex-and-ethnicity-specific intercepts (Asian females = 77.62; Asian males = 83.58; black females = 79.55; black males = 85.8; white females = 84.67; white males = 90.92). UL is calculated using the appropriate ethnicity-specific slope, and an intermediate intercept of *(female intercept + male intercept)/2)*—i.e. UL *(Asian) = (PMSH−((77.62 + 83.58)/2))/3.26*; *UL (black) = (PMSH−((79.55 + 85.8)/2))/3.14*; and *UL (white) = (PMSH−((84.67 + 90.92)/2))/3.14*—to eliminate any possible artefactual sexual dimorphism arising from the use of sex-specific equations. **d** UL is not estimated, and muscle quality is instead calculated as *HGS/(MM/PMSH)*, assuming simple isometric scaling of UL with PMSH. The primary finding of substantially greater age-related muscle quality decline in females persists across all scenarios, confirming that it is not simply an artefact of our UL estimates.

**Formal model comparison of muscle quality measures in SD6.I (equivalent to ‘MQ Model 1’ in Supplementary Data 2)**

*Muscle quality* ∼ sex + s(age) + <u>s(age, by = sex)</u> + ethnicity + season of attendance + s(UKB Assessment Centre, bs = "re") + s(dynamometer instance, bs = "re")*

*+ s(dynamometer instance, device day of use, bs = "re") + s(BIA impedance device, bs = “re”) + s(BIA impedance device, device day of use, bs = "re")*

**\*panel a AIC_GLOBAL_** 4,138,970; **AIC_SEX-SPECIFIC_** 4,135,609; **ΔAIC** 3,361; **prob_AW_** >99.9%

**panel b AIC_GLOBAL_** 4,147,876; **AIC_SEX-SPECIFIC_** 4,144,731; **ΔAIC** 3,145; **prob_AW_** >99.9%

**panel c AIC_GLOBAL_** 4,142,129; **AIC_SEX-SPECIFIC_** 4,139,456; **ΔAIC** 2,673; **prob_AW_** >99.9%

**panel d AIC_GLOBAL_** 5,494,340; **AIC_SEX-SPECIFIC_** 5,490,899; **ΔAIC** 3,441; **prob_AW_** >99.9%

#### Muscle quality measures and associated anthropometric variables have adequately normal distributions

**Figure SD6.III.**
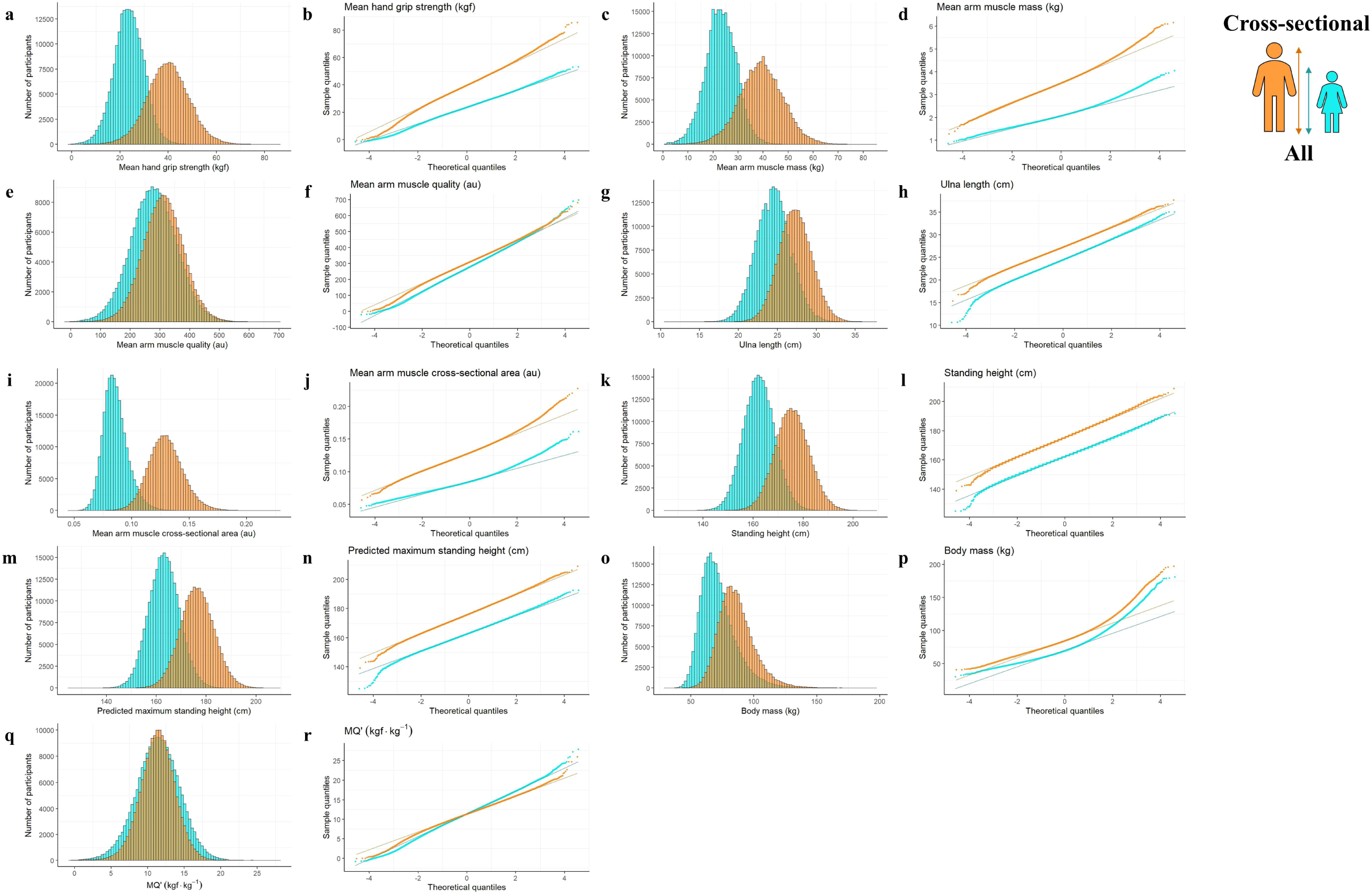
Muscle quality measures and associated anthropometric variables have adequately normal distributions. Values are for Cross-sectional Participants at baseline assessment (females, teal; males, orange). Histograms and qq-plots show mean hand grip strength (HGS) (**a**–**b**), mean arm muscle mass (MM) (**c–d**), mean arm muscle quality (MQ) (**e–f**), ulna length (**g– h**), mean arm cross-sectional area (**i–j**), standing height (**k–l**), predicted maximum standing height (**m–n**), body mass (**o–p**), and MQ’ (see Supplementary Fig. 8) (**q–r**).

**Table SD6.I.** Muscle quality measures and associated anthropometric variables have adequately normal distributions. Values are for Cross-sectional Participants at baseline assessment. West et al. (1995) consider skewness values <|2| and kurtosis values <|7| to indicate adequately normal distributions (referenced in main manuscript). Furthermore, non-normality is unlikely to be problematic in large public health data sets (e.g. UK Biobank) when sample sizes exceed ∼500, even when data are extremely non-normal (Lumley et al. 2002, *Annu. Rev. Public Health* 23:151–169.; DOI: 10.1146/annurev.publheath.23.100901.140546).

|  | Females |  | Males |  |
| --- | --- | --- | --- | --- |
|  | Skewness | Kurtosis | Skewness | Kurtosis |
| Mean hand grip strength (HGS) (kgf) | -0.05 | 3.25 | -0.01 | 3.41 |
| Mean arm muscle mass (MM) (kg) | 0.55 | 3.83 | 0.30 | 3.45 |
| Mean arm muscle quality (au) | 0.02 | 3.15 | -0.03 | 3.31 |
| Ulna length (cm) | 0.04 | 3.16 | 0.06 | 3.13 |
| Mean arm cross-sectional area (cm) | 0.68 | 4.02 | 0.30 | 3.53 |
| Standing height (cm) | 0.03 | 3.15 | 0.05 | 3.12 |
| Predicted maximum standing height (cm) | 0.02 | 3.16 | 0.04 | 3.13 |
| Body mass (kg) | 1.11 | 5.20 | 0.89 | 4.98 |
| MQ' (i.e. HGS/MM) (kgf·kg<sup>-1</sup>) | -0.09 | 3.20 | -0.14 | 3.40 |

### Supplementary Data 7 Controlling for body size

#### Participant matching

**Figure SD7.I.**
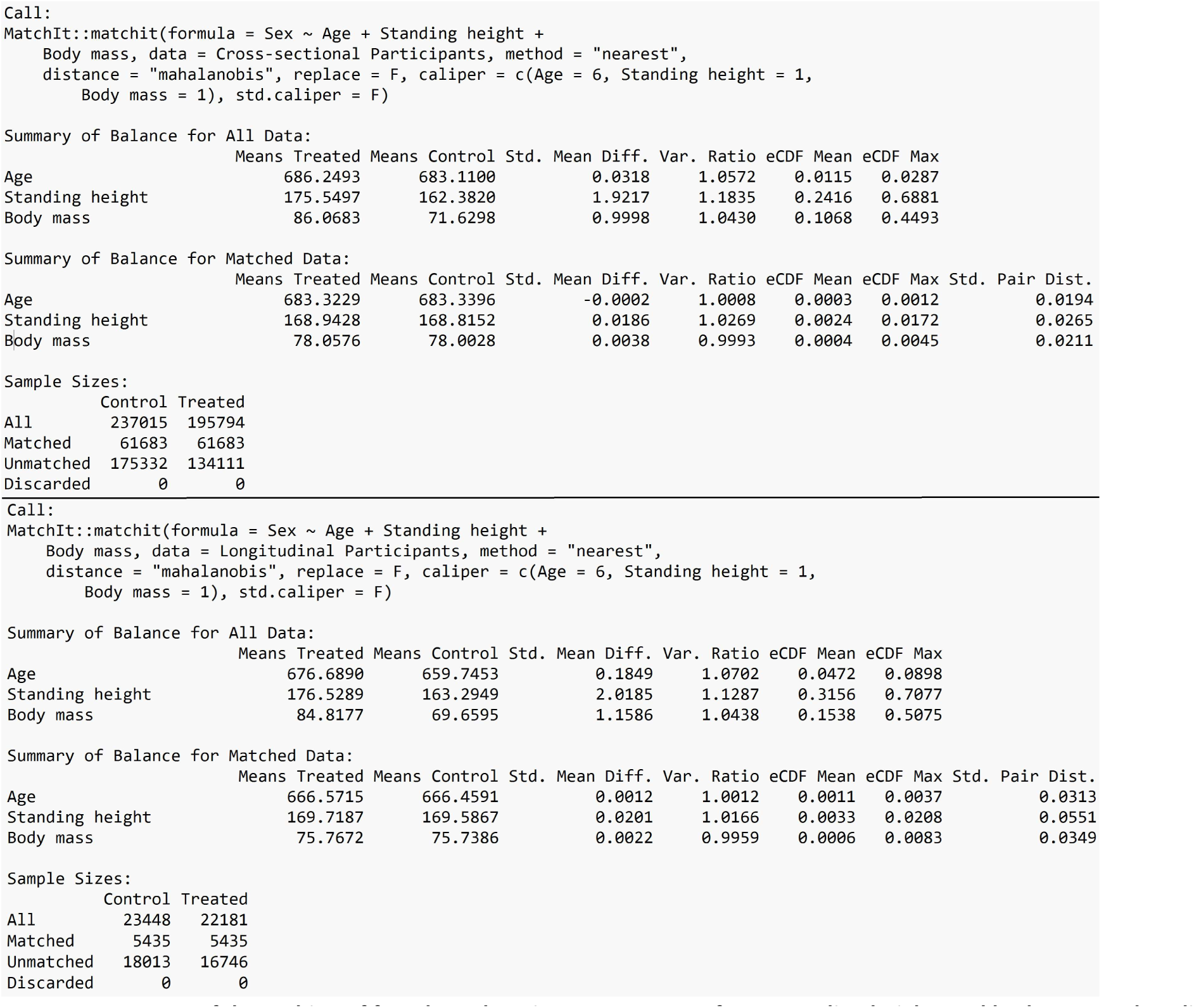
Successful matching of female:male pairs on measures of age, standing height, and body mass at baseline assessment. Summaries of the matching procedures used to generate the Matched Cross-sectional Participants (left) and the Matched Longitudinal Participants (right) participant groups (Methods), based on age (months), standing height (cm), and body mass (kg) at baseline assessment. Standard Mean Differences between −0.0002 and 0.0201 and Variance Ratios between 0.9959 and 1.0269 indicate excellent balance (i.e. successful matching), as per the recommendations of Greifer (2023) (referenced in main manuscript).

#### Controlling for body size within GAMMs

To identify the optimal body-size covariates to include in GAMMs, we first compared HGS model 1_GLOBAL_ (Supplementary Data 2) (here ‘HGS Model A’ to differentiate GAMMs discussed in this Supplementary Data from those presented in the main manuscript)—which does not incorporate body size—with two models incorporating standing height and body mass. In the first of these two models (HGS Model B) covariates were added as standing height^2^ and body mass^2/3^, which are the appropriate scaling factors for HGS under an assumption of isometric scaling (Jaric, 2002—referenced in main manuscript). In the second body-size model (HGS Model C), standing height and body mass were instead fitted as global splines:

**HGS Model A**

*HGS ∼ sex + s(age) + ethnicity + season of attendance + s(UKB Assessment Centre, bs = "re") + s(dynamometer instance, bs = "re") + s(dynamometer instance, device day of use, bs = "re")*

**HGS Model B**

*HGS ∼ sex + s(age) + ethnicity + season of attendance + s(UKB Assessment Centre, bs = "re") + s(dynamometer instance, bs = "re") + s(dynamometer instance, device day of use, bs = "re") + standing height*^2^ *+ body mass*^2*/*3^

**HGS Model C**

*HGS ∼ sex + s(age) + ethnicity + season of attendance + s(UKB Assessment Centre, bs = "re") + s(dynamometer instance, bs = "re") + s(dynamometer instance, device day of use, bs = "re") + s(standing height) + s(body mass)*

Multimodel comparison was then performed as per Methods, except that all three approximating models were compared simultaneously:

**Table SD7.I.** AIC scores and prob_AW_s for a three-way model comparison of HGS Models A–C.

|  | HGS Model A | HGS Model B | HGS Model C |
| --- | --- | --- | --- |
| AIC score | 2,441,316 | 2,407,242 | 2,406,431 |
| (prob<sub>AW</sub>) | (<0.1%) | (<0.1%) | (>99.9%) |

Model comparison unequivocally supported HGS Model C as the best approximating model in the candidate set (prob_AW_ >99.9%), thereby confirming that HGS does scale with body size, and that this scaling is allometric rather than isometric. We therefore retained HGS Model C for inclusion in subsequent model comparisons.

In order to test for sex-specific effects on HGS specifically attributable to ageing, and not mediated by possible sex-specific age-related differences in body size, we performed a four-way model comparison including HGS Model C and GAMMs incorporating sex-specific age splines (HGS Model D), sex-specific body size splines (HGS Model E), and sex-specific splines for both age and body size (HGS Model F):

**HGS Model D**

*HGS ∼ sex + s(age) + s(age, by = sex) + ethnicity + season of attendance + s(UKB Assessment Centre, bs = "re") + s(dynamometer instance, bs = "re") + s(dynamometer instance, device day of use, bs = "re") + s(standing height) + s(body mass)*

**HGS Model E**

*HGS ∼ sex + s(age) + ethnicity + season of attendance + s(UKB Assessment Centre, bs = "re") + s(dynamometer instance, bs = "re") + s(dynamometer instance, device day of use, bs = "re") + s(standing height) + s(body mass) + s(standing height, by = sex) + s(body mass, by = sex)*

**HGS Model F**

*HGS ∼ sex + s(age) + s(age, by = sex) + ethnicity + season of attendance + s(UKB Assessment Centre, bs = "re") + s(dynamometer instance, bs = "re") + s(dynamometer instance, device day of use, bs = "re") + s(standing height) + s(body mass) + s(standing height, by = sex) + s(body mass, by = sex)*

**Table SD7.II.** AIC scores and prob_AW_s for a four-way model comparison of HGS Models C–F.

|  | HGS Model C<br>(no sex-specific terms) | HGS Model D<br>(sex-specific ageing terms) | HGS Model E<br>(sex-specific body-size terms) | HGS Model F<br>(sex-specific ageing and body-size terms) |
| --- | --- | --- | --- | --- |
| AIC score<br>(prob<sub>AW</sub>) | 2,406,431<br>(<0.1%) | 2,405,983<br>(<0.1%) | 2,403,705<br>(<0.1%) | 2,403,315<br>(>99.9%) |

HGS Model F was unequivocally supported as being the best approximating model in the candidate set (prob_AW_ >99.9%). This analysis confirmed that i) inclusion of sex-specific body-size splines substantially improved model fit over global body-size splines alone (e.g. HGS Model D vs HGS Model F; ΔAIC = 2,668; prob_AW_ >99.9%), and ii) a sex difference in the HGS-age relationship persists after accounting for global and sex-specific relationships between HGS and body size (HGS Model E vs HGS Model F; ΔAIC = 390; prob_AW_ >99.9%).

Analogous two-step model comparison procedures were then performed for MM and MQ, albeit with different isometric scaling factors used for MM (standing height^3^ and body mass^1^) and MQ (standing height^0^ and body mass^0^). Although MQ was not predicted to scale with body size at all under simple isometric scaling, we nevertheless had reason to believe that it might do so in practice, for example due to the the potential relationship between standing height and forearm muscle pennation angle (see Supplementary Discussion). Results of the MM and MQ comparisons are presented below:

**MM Model A**

*MM ∼ sex + s(age) + ethnicity + season of attendance + s(UKB Assessment Centre, bs = "re") + s(BIA impedance device, bs = “re”) + s(BIA impedance device, device day of use, bs = "re")*

**MM Model B**

*MM ∼ sex + s(age) + ethnicity + season of attendance + s(UKB Assessment Centre, bs = "re") + s(BIA impedance device, bs = “re”) + s(BIA impedance device, device day of use, bs = "re") + standing height*^3^ *+ body mass*^1^

**MM Model C**

*MM ∼ sex + s(age) + ethnicity + season of attendance + s(UKB Assessment Centre, bs = "re") + s(BIA impedance device, bs = “re”) + s(BIA impedance device, device day of use, bs = "re") + s(standing height) + s(body mass)*

**MM Model D**

*MM ∼ sex + s(age) + s(age, by = sex) + ethnicity + season of attendance + s(UKB Assessment Centre, bs = "re") + s(BIA impedance device, bs = “re”) + s(BIA impedance device, device day of use, bs = "re") + s(standing height) + s(body mass)*

**MM Model E**

*MM ∼ sex + s(age) + ethnicity + season of attendance + s(UKB Assessment Centre, bs = "re") + s(BIA impedance device, bs = “re”) + s(BIA impedance device, device day of use, bs = "re") + s(standing height) + s(body mass) + s(standing height, by = sex) + s(body mass, by = sex)*

**MM Model F**

*MM ∼ sex + s(age) + s(age, by = sex) + ethnicity + season of attendance + s(UKB Assessment Centre, bs = "re") + s(BIA impedance device, bs = “re”) + s(BIA impedance device, device day of use, bs = "re") + s(standing height) + s(body mass) + s(standing height, by = sex) + s(body mass, by = sex)*

**Table SD7.III.**
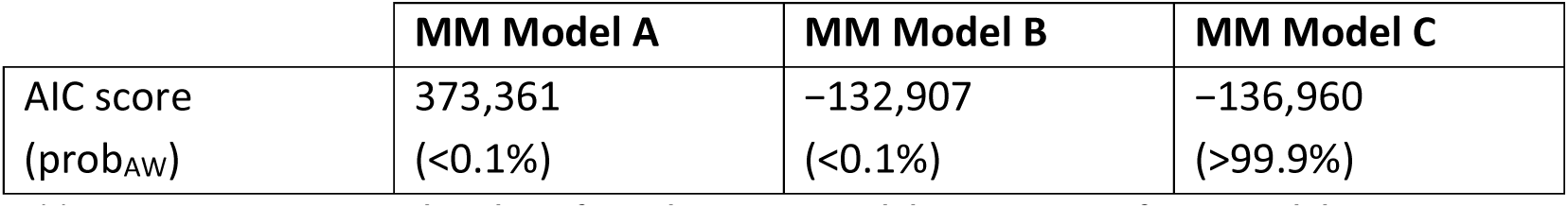
AIC scores and prob_AW_s for a three-way model comparison of MM Models A–C.

|  | **MM Model A** | **MM Model B** | **MM Model C** |
| --- | --- | --- | --- |
| AIC score | 373,361 | -132,907 | -136,960 |
| (prob<sub>AW</sub>) | (<0.1%) | (<0.1%) | (>99.9%) |

**Table SD7.IV.** AIC scores and prob_AW_s for a four-way model comparison of MM Models C–F.

Table SD7.III. AIC scores and prob<sub>AW</sub>s for a three-way model comparison of MM Models A–C.
|  | **MM Model C**<br>(no sex-specific terms) | **MM Model D**<br>(sex-specific ageing terms) | **MM Model E**<br>(sex-specific body-size terms) | **MM Model F**<br>(sex-specific ageing and body-size terms) |
| --- | --- | --- | --- | --- |
| AIC score<br>(prob<sub>AW</sub>) | -136,960<br>(<0.1%) | -148,393<br>(<0.1%) | -162,851<br>(<0.1%) | -174,605<br>(>99.9%) |

**MQ Model A**

*MQ ∼ sex + s(age) + ethnicity + season of attendance + s(UKB Assessment Centre, bs = "re") + s(dynamometer instance, bs = "re") + s(dynamometer instance, device day of use, bs = "re") + s(BIA impedance device, bs = “re”) + s(BIA impedance device, device day of use, bs = "re")*

**MQ Model B**

*MQ ∼ sex + s(age) + ethnicity + season of attendance + s(UKB Assessment Centre, bs = "re") + s(dynamometer instance, bs = "re") + s(dynamometer instance, device day of use, bs = "re") + s(BIA impedance device, bs = “re”) + s(BIA impedance device, device day of use, bs = "re") + standing height*^0^ *+ body mass*^0^

**MQ Model C**

*MQ ∼ sex + s(age) + ethnicity + season of attendance + s(UKB Assessment Centre, bs = "re") + s(dynamometer instance, bs = "re") + s(dynamometer instance, device day of use, bs = "re") + s(BIA impedance device, bs = “re”) + s(BIA impedance device, device day of use, bs = "re") + s(standing height) + s(body mass)*

**MQ Model D**

*MQ ∼ sex + s(age) + s(age, by = sex) + ethnicity + season of attendance + s(UKB Assessment Centre, bs = "re") + s(dynamometer instance, bs = "re") + s(dynamometer instance, device day of use, bs = "re") + s(BIA impedance device, bs = “re”) + s(BIA impedance device, device day of use, bs = "re") + s(standing height) + s(body mass)*

**MQ Model E**

*MQ ∼ sex + s(age) + ethnicity + season of attendance + s(UKB Assessment Centre, bs = "re") + s(dynamometer instance, bs = "re") + s(dynamometer instance, device day of use, bs = "re") + s(BIA impedance device, bs = “re”) + s(BIA impedance device, device day of use, bs = "re") + s(standing height) + s(body mass) + s(standing height, by = sex) + s(body mass, by = sex)*

**Table SD7.V.** AIC scores and prob_AW_s for a three-way model comparison of MQ Models A–C. Note that MQ is not expected to scale with body size, meaning that MQ Models A and B are identical in practice. However, MQ Model B is included here to aid interpretation of results by keeping presentation consistent across HGS, MM, and MQ comparisons.

|  | **MQ Model A** | **MQ Model B** | **MQ Model C** |
| --- | --- | --- | --- |
| AIC score<br>(prob<sub>AW</sub>) | 4,145,095<br>(<0.1%) | 4,145,095<br>(<0.1%) | 4,036,991<br>(>99.9%) |

**Table SD7.VI.** AIC scores and prob_AW_s for a four-way model comparison of MQ Models C–F.

|  | **MQ Model C**<br>(no sex-specific terms) | **MQ Model D**<br>(sex-specific ageing terms) | **MQ Model E**<br>(sex-specific body-size terms) | **MQ Model F**<br>(sex-specific ageing and body-size terms) |
| --- | --- | --- | --- | --- |
| AIC score<br>(prob<sub>AW</sub>) | 4,036,991<br>(<0.1%) | 4,033,771<br>(<0.1%) | 4,036,496<br>(<0.1%) | 4,033,275<br>(>99.9%) |

**MQ Model F**

*MQ ∼ sex + s(age) + s(age, by = sex) + ethnicity + season of attendance + s(UKB Assessment Centre, bs = "re") + s(dynamometer instance, bs = "re") + s(dynamometer instance, device day of use, bs = "re") + s(BIA impedance device, bs = “re”) + s(BIA impedance device, device day of use, bs = "re") + s(standing height) + s(body mass) + s(standing height, by = sex) + s(body mass, by = sex)*

MM and MQ analysis accorded with HGS analysis in identifying Model F as the best-supported approximating model in the candidate set (each prob_AW_ >99.9%). We interpreted this as evidence that controlling for body size within GAMMs was best achieved by including both global and sex-specific body-size splines. Sex-specific terms were substituted for group-specific terms (i.e. Premenopausal females, Postmenopausal females, and Males) in models fitted on the Menopause Participants (Methods; Supplementary Data 2).

## Footnotes

[a] ‘Asian’, ‘black’, ‘Chinese’, ‘mixed’, ‘other’, ‘unknown’, or ‘white’, as determined from the ‘Ethnic background’ data field (Data-Field 21000).

## Notes

### Competing Interest Statement

The authors have declared no competing interest.

### Funding Statement

This study was funded by Medical Research Council Scientist Programme Grant MR/N021231/1.

### Author Declarations

The North West Multi-centre Research Ethics Committee gave ethical approval for this work.

### Summary of Updates

Revised manuscript. The most substantial of these changes are briefly described below: (i) We now present a second, additional muscle quality measure (MQ prime), defined as [hand grip strength / arm muscle mass]. The findings of our MQ prime analysis are consistent with those of our original muscle quality measure (with MQ defined as [hand grip strength / arm muscle cross-sectional area]). Each measure represents a feasible biomechanical model of muscle quality under different assumptions of arm muscle fibre pennation; that is, the angle of insertion of muscle fibres into their tendons. The confirmation of marked sexual dimorphism under both models increases confidence in our finding of greater age-related muscle quality decline in females. (ii) We have incorporated sex hormone (testosterone and oestradiol) data into the manuscript, and demonstrate that observed sex differences in skeletal muscle ageing do not reflect age-sex differences in levels of circulating sex hormones. (iii) We have re-fitted our generalised additive mixed models with global and sex-specific body-size (standing height and body mass) splines, and demonstrate formally that doing so improves model fit when accounting for systematic sex differences in body size between the sexes. This addresses concerns that, given marked female/male differences in body size and composition, assuming any particular relationship between body size and muscle properties might artefactually create dimorphism in muscle properties. (iv) We have included multiple linear regression modelling of observed data as an additional form of regression modelling, as this method produces estimates of sex-specific annual rates of decline in muscle strength, mass, and quality that are more robust than those that we originally presented. (v) We have made various changes to improve readability, including moving some of the more technical parts of the discussion into a Supplementary Discussion to limit overall word count.

